# Sex-stratified GWAS of Body Fat Percentage after Adjusting for Testosterone and SHBG in the UK Biobank

**DOI:** 10.1101/2023.03.09.23287061

**Authors:** Delnaz Roshandel, Andrew D. Paterson, Satya Dash

## Abstract

**Introduction:** Adiposity, particularly centripetal adiposity/reduced gluteofemoral adiposity, increases dyslipidemia, type 2 diabetes (T2D) and coronary artery disease (CAD). Genome-wide association studies (GWAS) to date have identified 12 loci associated with body fat percentage (BFP). Biological sex influences both overall adiposity and fat distribution. Further, testosterone and sex hormone binding globulin (SHBG) influence adiposity and metabolic function, with differential effects of testosterone in men and women.

**Methods:** We performed sex-stratified GWAS of BFP in white British individuals from the UK biobank adjusting for SHBG and testosterone. We further investigated association of the identified loci with high density cholesterol (HDL), triglyceride (TG), T2D, CAD, and MRI-derived abdominal subcutaneous adipose tissue (ASAT), visceral adipose tissue (VAT) and gluteofemoral adipose tissue (GFAT) using publicly available data from large GWAS. We also performed 2-sample Mendelian Randomization (MR) using identified BFP variants as instruments to investigate causal effect of BFP on HDL, TG, T2D and CAD in males and females separately.

**Results:** We identified 193 and 174 autosomal loci explaining 3.35% and 2.60% of the variation in BFP in males and females, respectively. In addition, we identified 2 Chr X loci in men. Only 38 of these loci associated with BFP in both males and females. Seven loci in men including the 2 loci on Chr X and ten loci in females have not been associated with any adiposity or cardiometabolic traits previously. The majority of BFP loci did not associate with cardiometabolic traits. Of the BFP loci associated with cardiometabolic traits several had paradoxically beneficial cardiometabolic effects with favourable fat distribution. Consistent with that sex-stratified MR analyses using identified BFP variants as instruments did not find convincing supportive evidence that increased BFP has deleterious cardiometabolic effects in either sex with highly significant heterogeneity.

**Conclusions:** Adjusting for testosterone and SHBG with sex-stratified analyses substantially increased the number of BFP associated loci. Despite this adjustment, there was limited genetic overlap in BFP in males and females. Further identified loci in general did not have adverse cardiometabolic effects which may reflect the protective effect of favourable fat distribution and the modulation of cardiometabolic risk by testosterone and SHBG.

## INTRODUCTION

Obesity is a chronic multisystem disease which affects more than 600 million adults and 100 million children ^1^. Cardiometabolic diseases/traits such as insulin resistance, dyslipidemia, type 2 diabetes (T2D) and coronary artery disease (CAD) are leading causes of morbidity and mortality for people living with obesity ^2^.

Obesity/adiposity are highly heritable: to date more than 1000 genetic loci have been associated with adiposity and related traits ^3,4^. Although obesity generally increases the risk of cardiometabolic disease, this can be further modulated by fat distribution ^5-8^. Some adiposity associated loci are paradoxically associated with improved cardiometabolic profile. This is likely in part because these loci associate with ‘favourable’ fat distribution with increased subcutaneous femoro-gluteal adiposity and/or reduced centripetal/visceral adiposity ^5,6,8-10^.

Body mass index (BMI) is commonly used to diagnose obesity but is an imperfect measure of overall adiposity ^11^. A previous GWAS of body fat percentage (BFP) in the UK Biobank identified 12 loci ^12^. Sex and sex hormones differentially impact adiposity ^13^. On average females have higher BFP on average with ‘favourable’ fat distribution. Genetic data indicate that increased testosterone likely has beneficial effects on adiposity and metabolic traits in men, but may be deleterious in women, consistent with some observational data ^13-18^. Sex hormone binding globulin (SHBG) modulates bioavailable sex hormone concentration and may have independent effects on adiposity and cardiometabolic traits ^13^. Given these important sex differences, we undertook sex-stratified genome-wide association studies (GWAS) of BFP adjusted for total testosterone and SHBG in the UK Biobank. We hypothesized this would increase statistical power to detect sex-specific BFP associated loci. We further investigated the association of identified loci with fat distribution, high density lipoprotein cholesterol (HDL), triglyceride (TG), T2D and CAD. We identified a total of 329 autosomal loci, 193 in males and 174 in females. Only 38 loci were common between the two sexes. This is a significant improvement from the previous published GWAS of BFP where they reported only 12 loci ^12^ of which 9 were replicated in our analysis. Of these 329 loci, 15 (5 in males and 10 in females) have not been associated with any adiposity related traits previously. We also identified multiple loci in males and a single locus in females on chromosome X associated with BFP of which two loci in males have not been associated with any adiposity related traits previously.

## METHODS

Ethics approval for this study was obtained in the Hospital for Sick Children (HSC #1000073707).

### Study Population

All white British subjects (Unique Data Identifier (UDI) 22006-0.0 = 1) from the UK Biobank who had no missing data for BFP (UDI 23099-0.0), age when attended assessment centre (UDI 21003-0.0), serum albumin (UDI 30600-0.0), SHBG (UDI 30830-0.0) or serum testosterone (UDI 30850-0.0) were included in the analysis. Subjects with sex chromosome aneuploidy (UDI 22019-0.0 = 1) and those excluded from kinship inference process or had ten or more third-degree relatives (UDI 22021-0.0 = 10 or -1) were excluded from the analysis. Sex was defined using UDI 22001-0.0 (female = 0, male = 1). Participants who were on medical treatments that may interfere with sex hormones were also excluded (N = 1,683 males & 1,628 females). The full list of these medications is provided in Table S1. Females with testosterone levels >10 nmol/L were also excluded (N = 28,471).

Multivariable linear regression was used for testing the association of covariates (i.e. age, age^2^, serum albumin, centered albumin^2^, serum SHBG, centered SHBG^2^, serum testosterone, centered testosterone^2^) with BFP in males and females separately using R v3.5. To test BFP mean and variance difference in males and females, t.test and var.test were used respectively in R v4.2.1 ^19^.

### GWAS

GWAS (Chr1-23) were performed using REGENIE (v3.1.1) ^20^ on research analysis platform (RAP). In step 1, only genotyped single nucleotide polymorphisms (SNPs) with minor allele count (MAC) >80 (minor allele frequency ∼ 0.001) and Hardy-Weinberg Equilibrium (HWE) p >1E-15 were included in the analysis. SNPs with inter-chromosome linkage disequilibrium (LD) ^20^ were excluded. In step 2, all SNPs comprising genotyped and centrally imputed to the Haplotype Reference Consortium (HRC) or the UK10K + 1000 Genomes phase 3 panel if the SNP was not available in HRC with MAC >80 and high imputation quality (INFO > 0.5) were included.

BFP (UDI 23099-0.0) measured by Tanita BC418MA body composition analyser was used as outcome. Previous analyses indicates that BFP assessed by bioimpedance is strongly associated with dual energy X-ray absorptiometry (DEXA) based measures of adiposity (Pearson correlation coefficient = 0.92) ^21^. Age (UDI 21003-0.0) and its quadratic term (centered age^2^), serum albumin (UDI 30600-0.0) and its quadratic term (centered albumin^2^), serum SHBG (UDI 30830-0.0) and its quadratic term (centered SHBG^2^), serum testosterone (UDI 30850-0.0) and its quadratic term (centered testosterone^2^), and first ten genetic principal components (PCs, UDI 22009-0.1-10) were included in the model as covariates. The GWAS was performed in males and females separately.

To identify independent GWAS signals, genome-wide significant (GWS, p < 5E-8) SNPs were clumped with r^2^ set at 0.1 and radius set at 500 (less than 250 kb away from an index variant) in PLINK v2 using a random 5000 sample of participants included in the GWAS as reference. These loci were excluded from further investigation if they were associated with SHBG or testosterone in their corresponding sex (p >5E-8) ^13^ (Figure 1, Table S2).

**Figure 1:**
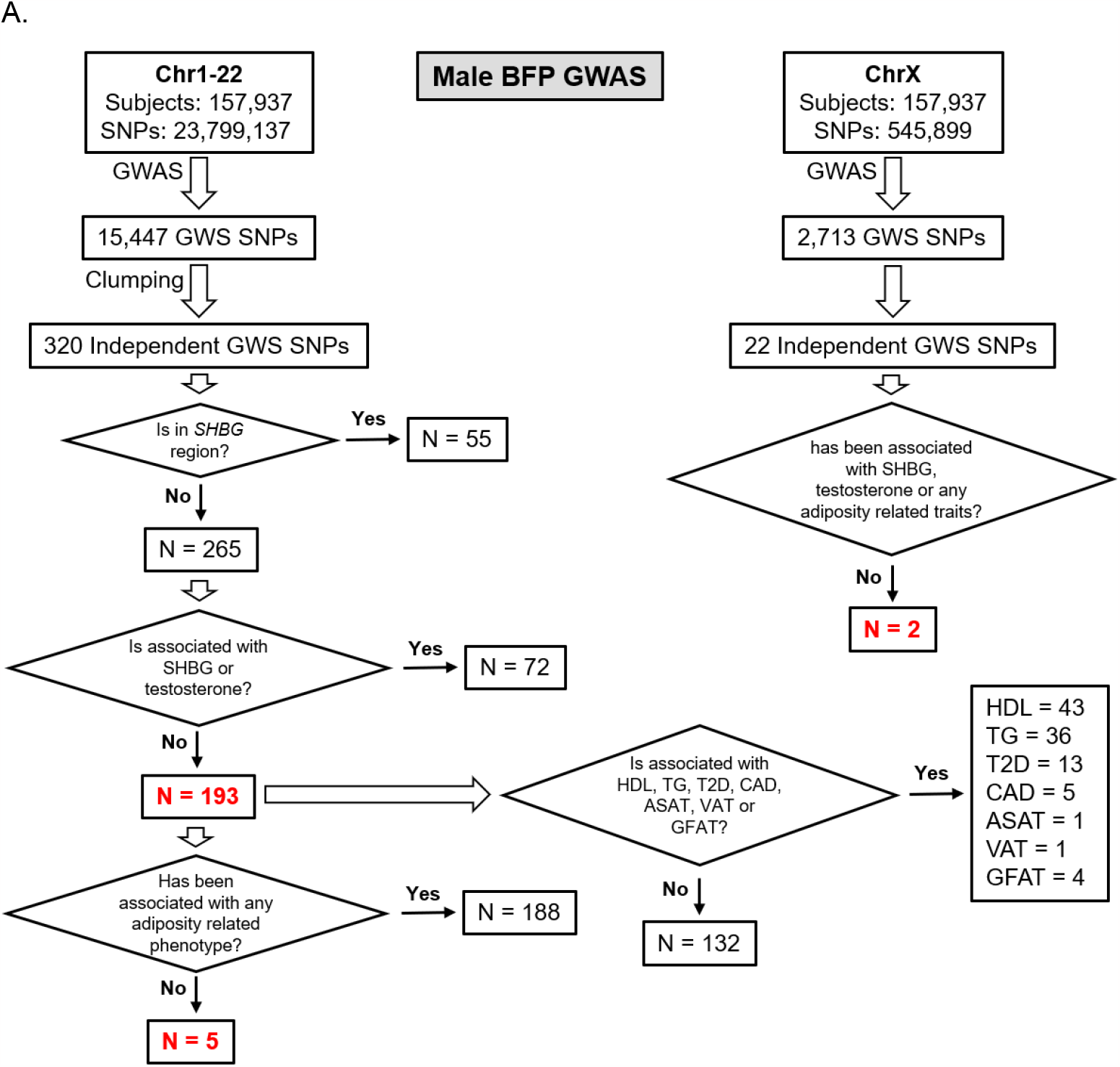
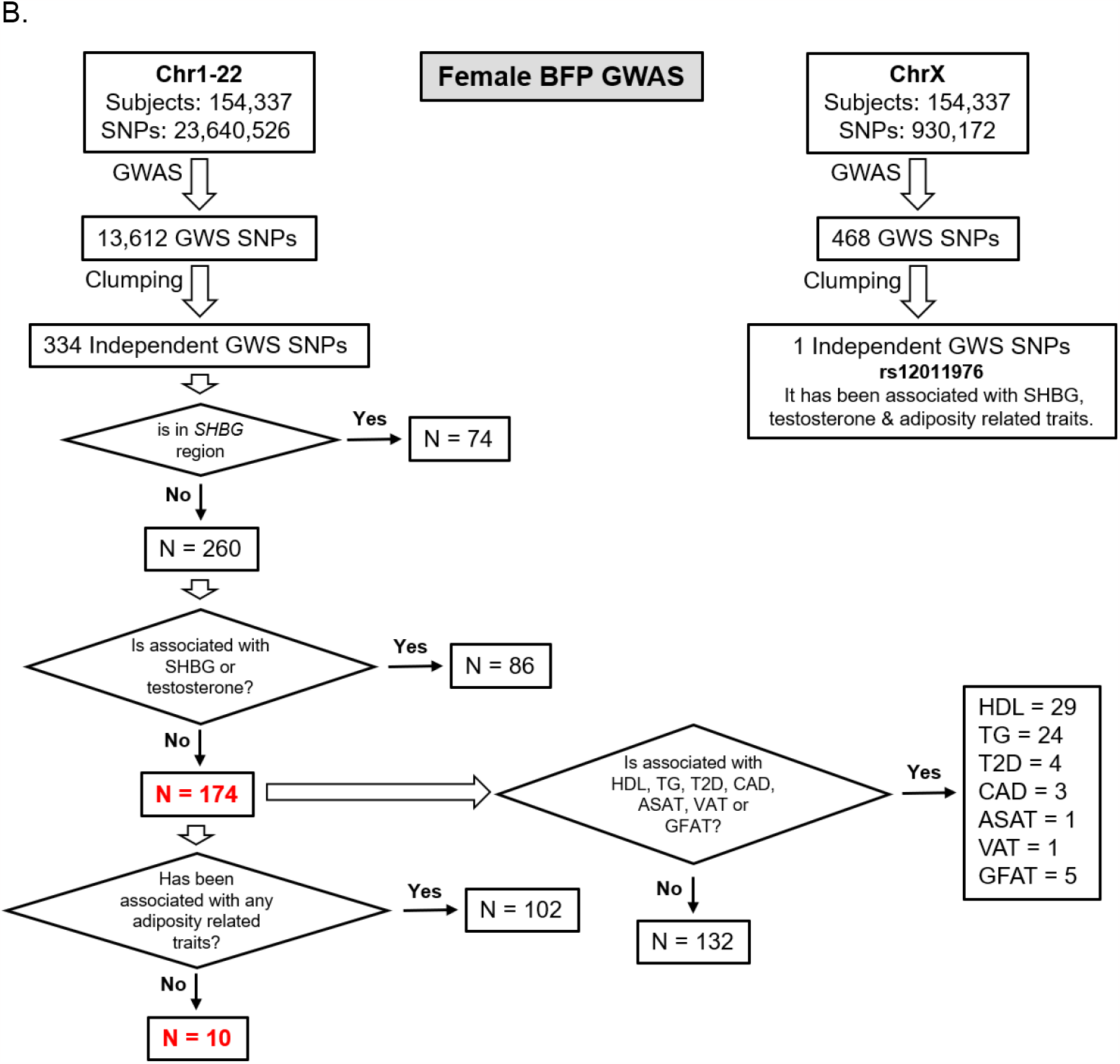
Study design and major findings

To calculate the variance in BFP explained by the identified SNPs in each sex, we performed clumping with a much stricter r^2^ (< 0.001) and used linear regression in R v4.2.1 ^19^ with the same covariates included in the GWAS to test the association of individual SNPs with BFP. The variance explained by each SNP was calculated by subtracting the base model (only covariates in the model) R^2^ from the full model (SNP + all covariates) R^2^. Subsequently, the total variance explained was calculated as sum of the variance explained by individual SNPs.

Association of identified independent loci were investigated with T2D ^22^, CAD ^23^, sex-stratified HDL ^24^ and TG ^24^. We also investigated association of identified independent loci with MRI-derived abdominal subcutaneous adipose tissue (ASAT), visceral adipose tissue (VAT) and gluteofemoral adipose tissue (GFAT) adjusted for BMI and height in their corresponding sex ^10^. For SNPs having paradoxical effect on BFP and lipid levels, we also investigated their association with waist-hip ratio (WHR) adjusted for body mas index (BMI) ^3^ (Figure 1, Table S2).

To identify novel loci, we investigated if the SNPs were associated with BFP ^12^, BMI ^3^, WHR ^3^, or any other adiposity related phenotypes (e.g. appendicular lean mass ^25^) previously, by examining the NHGRI-EBI GWAS catalogue (https://www.ebi.ac.uk/gwas, accessed Nov 2022 ^26^) and Neale’s round 2 GWAS results (http://www.nealelab.is/uk-biobank; UKBB GWAS Imputed v3 - File Manifest Release 20180731).

We also performed a separate GWAS (Chr1-22) including both sexes to investigate the SNP x Sex interaction using REGENIE. Same SNP inclusion criteria were used and same covariates plus sex were included in the model. The results of the test for interaction effect (i.e. ADD-INT_SNPxSex=0) were reported.

### HLA Imputation

Association of 362 four-digit HLA haplotypes (UDI 22182-0.0) were tested with BFP in males and females separately using linear regression in R v4.2.1 ^19^ with age, age^2^, albumin, albumin^2^, SHBG, SHBG^2^, testosterone, testosterone^2^ and first ten genetic PCs as covariates in the model.

### Mendelian Randomization (MR)

Two-sample MR was used to investigate the causal effect of BFP (exposure) on T2D, CAD, HDL and TG (outcomes) in males and females separately using MR-Base platform ^27^. For exposure, we used summary statistics from our current analyses and performed the clumping with r^2^ <0.001 and a 5000 random sample of UK biobank participants as LD reference. For outcomes, we used publicly available summary statistics from the published GWAS as explained above (Table S2) ^22-24^. Palindromic SNPs with intermediate allele frequencies were excluded. We used five methods for MR analysis including MR Egger, inverse variance weighted (IVW), weighted median, simple mode and weighted mode. However, we primarily focused on MR Egger results. MR Egger relaxes “no horizontal pleiotropy” assumption (the effects of the SNPs on the outcome not mediated by the exposure) allowing the net-horizontal pleiotropic effect across all SNPs to be unbalanced or directional. It returns an unbiased causal effect even if the “no horizontal pleiotropy” assumption is violated for all SNPs. Heterogeneity was tested with MR Egger and IVW methods. Horizontal pleiotropy was tested using MR Egger. The Wald ratio method was used for single SNP MR and the IVW method was used for leave-one-out analysis ^27^.

## RESULTS

### Males

#### Autosomal GWAS

157,937 males were included in the analysis (Table S3). One male subject was excluded from analysis as his whole-body fat plus fat-free mass was greater than his weight. BFP was normally distributed with mean (SD) of 25.3 (5.8) % (Figure S1A). In the multivariable analysis, age was associated with higher BFP whereas albumin, SHBG and testosterone were all associated with lower BFP. Age, albumin, SHBG, testosterone, and their quadratic terms together explained 13% of variation in BFP. Testosterone and its quadratic term explained 8% of variation in BFP (Table S4 & S5).

23,799,137 SNPs on Chr1-22 were included in the GWAS (GC lambda = 1.20) (Figure 2A). 15,447 SNPs were associated with BFP at GWS level (Males.xlsx File, Sheet A). There were 320 independent GWS SNPs including 55 SNPs in *SHBG* (Chr7:7.4Mb; Males.xlsx File, Sheet B). Less than half (119 out of 320) of these independent GWS SNPs were associated with BFP in females, all with the same direction of effect as males (Males.xlsx File, Sheet C). Five SNPs in *SHBG* region were missing from Ruth et al analysis of SHBG/Testosterone; but the rest (N = 50) were all associated with SHBG and/or testosterone. Of 265 SNPs in non-*SHBG* loci, 193 (only 38 common with females) were not associated with SHBG or testosterone including an indel (Chr4:99262829, TG>T) missing in Ruth et al analysis (Males.xlsx File, Sheet D). After, further clumping of these 193 SNPs with r^2^ < 0.001, 161 SNP were left, and they explained 3.35% of the variation in BFP in males (Males.xlsx File, Sheet J).

**Figure 2:**
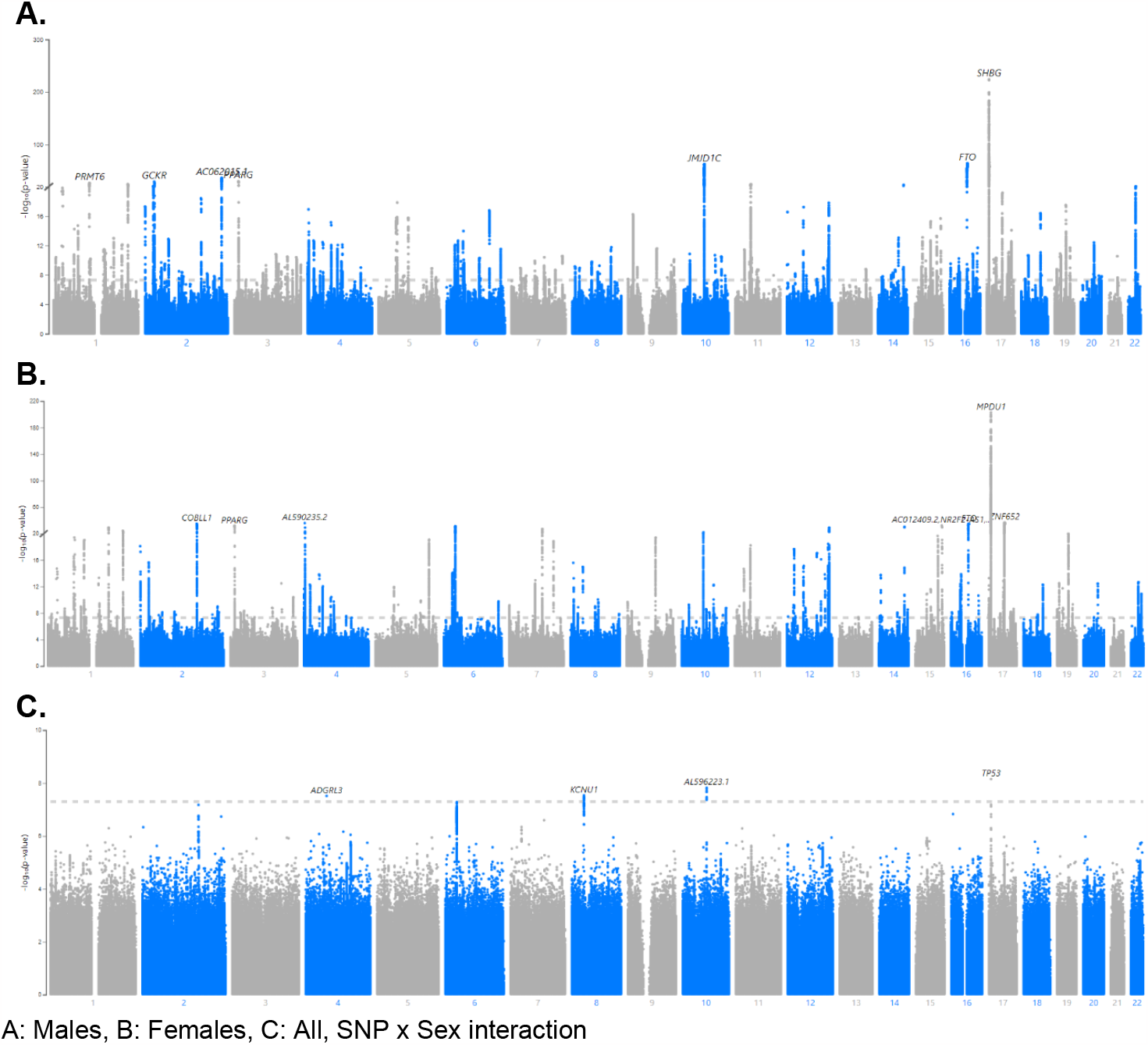
Manhattan plots for BFP GWAS in males, females and SNP*sex interaction

#### Novel autosomal BFP loci

Of these 193 SNPs, 188 were not associated with BFP in prior published GWAS ^12^ and 94 were not identified by Neale’s round 2 GWAS results of BFP in males (Males.xlsx File, Sheet I). Five of these 193 autosomal SNPs, have not been associated with any adiposity related phenotypes (Table 1), fat depots (Table S6), HDL, TG, T2D or CAD (Males.xlsx File, Sheet E) previously.

**Table 1:**
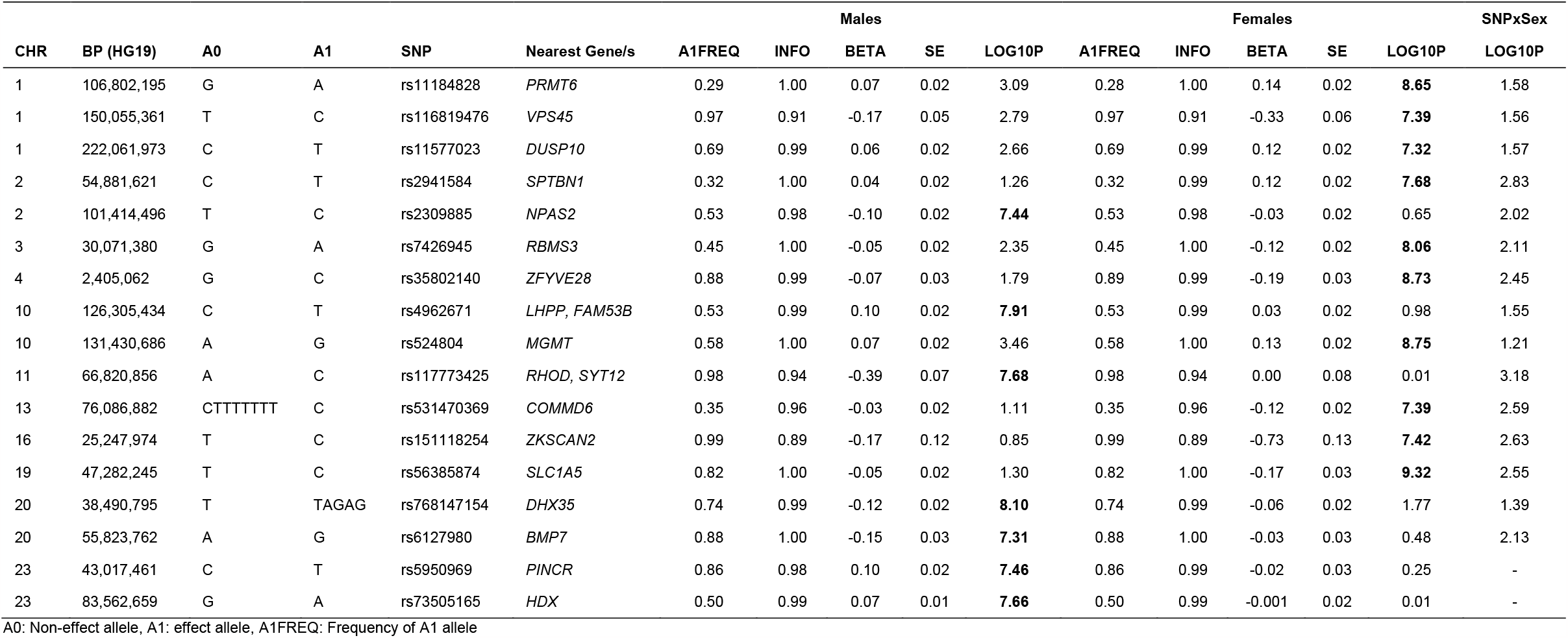
Newly identified loci for BFP in males or females not previously associated with adiposity related phenotypes

#### Association of autosomal BFP GWAS Loci with cardiometabolic phenotypes

Of 193 autosomal SNPs associated with BFP in males, the majority (N = 132, 68%) were not associated with cardiometabolic phenotypes. Sixty-one were associated with lipid levels, T2D, CAD or fat depots with some associated with multiple traits: HDL: 43, TG: 36, T2D: 13, CAD: 5, GFAT: 4, VAT: 1, and ASAT: 1 (Figure 3, Males.xlsx File, Sheet E). Twenty-three SNPs were associated with both HDL and TG. Of these, 22 had consistent effect on HDL and TG (i.e. the allele associated with higher HDL was associated with lower TG and vice versa). However, rs190712692 (Chr19:45,425,178 (GRCh37); A>G; near *APOC1*) associated with lower BFP (β (SE) = -0.24 (0.04), -log^10^(p) = 7.93) was associated with lower HDL (β (SE) = -0.070 (0.004), p = 3.4E-72) and lower TG (β (SE) = -0.154 (0.004), p = 4.70E-338). This SNP was also associated with increased risk of CAD (β (SE) 0.15 (0.01), p = 2.72E-27) (Table 2).

**Figure 3:**
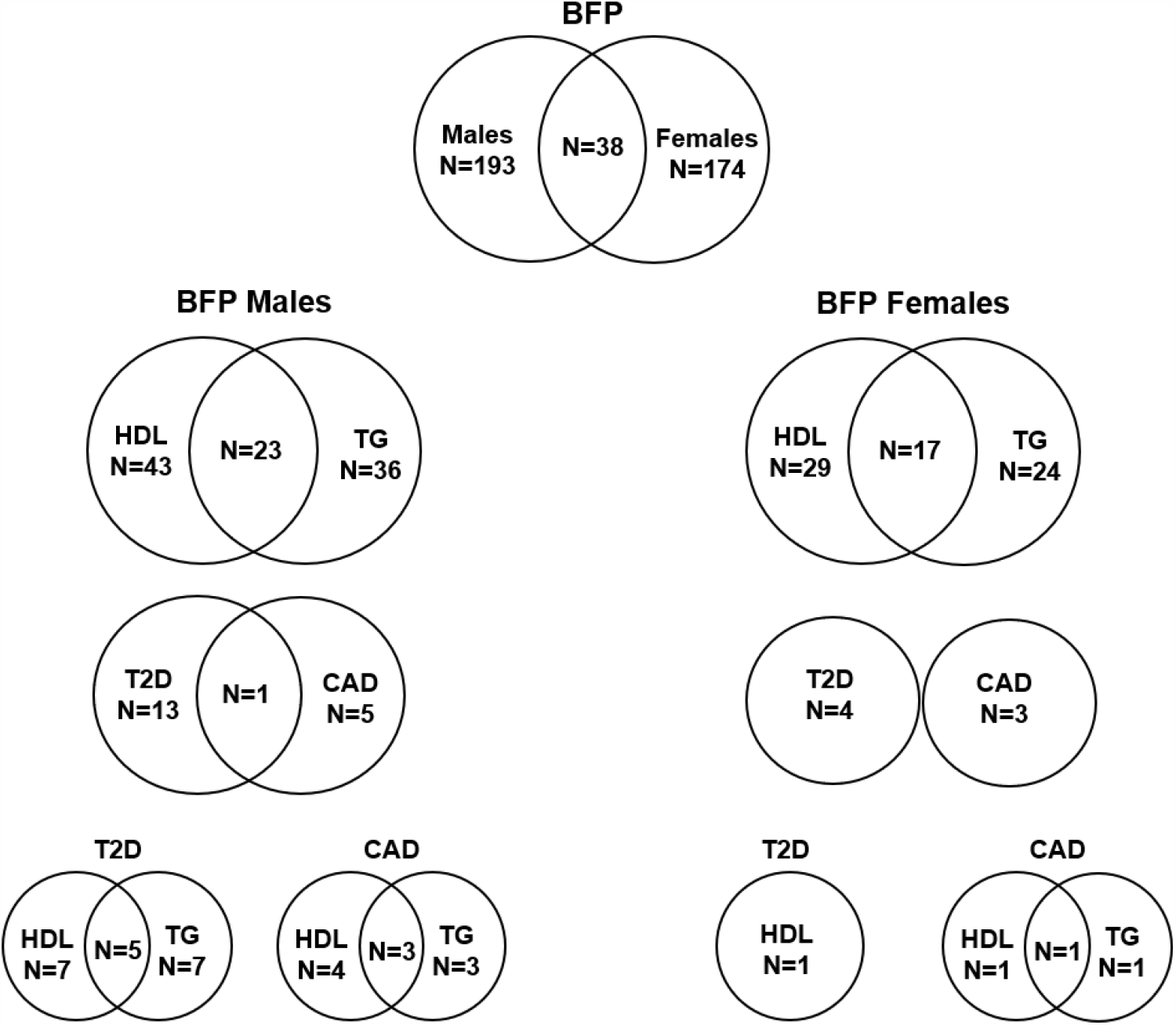
Number of independent SNPs associated with BFP in males and females, and of them the number of SNPs associated with HDL, TG, T2D and CAD The numbers in each circle shows the total number of SNPs associated with the corresponding trait and the numbers in the common area shows the number of SNPs associated with both phenotypes.

**Table 2:** BFP GWAS loci in males having paradoxical effect on HDL and TG

| Sex-Stratified |  |  |  |  |  |  |  |  |  |  |  |  |  |  |  |  |  |  |  |  |  |  |  | Sex-Combined |
| --- | --- | --- | --- | --- | --- | --- | --- | --- | --- | --- | --- | --- | --- | --- | --- | --- | --- | --- | --- | --- | --- | --- | --- | --- |
|  |  |  |  |  |  |  | BFP |  | WHR Adj BMI |  | HDL |  | TG |  | ASAT |  | VAT |  | GFAT |  | T2D |  | CAD |  |
| SNP | CHR | BP (HG19) | Gene | A0 | A1 | A1FREQ | β | LOG10P | β | LOG10P | β | LOG10P | β | LOG10P | B | LOG10P | β | LOG10P | β | LOG10P | β | LOG10P | β | LOG10P |
| rs78058190 | 2 | 219,699,999 | PRKAG3 | A | G | 0.95 | 0.26 | 7.51 | 0.008 | 0.51 | 0.09 | 73.09 | -0.07 | 57.05 | 0.13 | 5.39 | -0.07 | 1.96 | 0.15 | 7.32 | -0.08 | 6.35 | -0.07 | 6.85 |
| rs112695539 | 2 | 227,004,358 | NYAP2 | T | G | 0.96 | -0.27 | 8.48 | 0.000 | 0.002 | -0.03 | 13.82 | 0.03 | 12.43 | -0.09 | 3.43 | -0.05 | 1.24 | -0.09 | 3.64 | 0.07 | 4.96 | -0.004 | 0.11 |
| rs150535373 | 3 | 12,410,328 | PPARG | A | G | 0.99 | -0.53 | 11.21 | -0.023 | 1.10 | -0.05 | 10.48 | 0.05 | 12.47 | -0.17 | 4.18 | -0.01 | 0.00 | -0.13 | 2.72 | 0.13 | 6.22 | 0.04 | 1.24 |
| rs62271373 | 3 | 150,066,540 | TSC22D2 | A | T | 0.94 | 0.24 | 9.03 | -0.002 | 0.09 | 0.04 | 22.05 | -0.03 | 15.38 | 0.05 | 1.54 | 0.01 | 0.27 | 0.1 | 4.96 | -0.09 | 9.00 | -0.06 | 5.96 |
| rs71602277 | 4 | 157,714,979 | PDGFC | TA | T | 0.69 | -0.12 | 8.99 | NA | NA | -0.02 | 17.60 | 0.02 | 20.21 | -0.01 | 0.47 | 0.01 | 0.57 | -0.06 | 7.72 | NA | NA | 0.02 | 1.35 |
| rs998584 | 6 | 43,757,896 | VEGFA | A | C | 0.52 | 0.12 | 9.84 | -0.027 | 24.88 | 0.03 | 92.36 | -0.04 | 111.77 | 0.03 | 1.85 | -0.06 | 9.01 | 0.07 | 11.92 | -0.04 | 7.64 | -0.04 | 11.54 |
| rs9641894 | 7 | 130,465,054 | KLF14 | G | T | 0.58 | 0.11 | 8.66 | 0.008 | 2.73 | 0.02 | 27.41 | -0.01 | 13.15 | 0.004 | 0.17 | 0.02 | 1.19 | 0.003 | 0.06 | -0.05 | 13.41 | -0.02 | 3.42 |
| rs4741016 | 9 | 1,036,132 | DMRT2 | C | T | 0.77 | 0.12 | 7.43 | 0.005 | 0.80 | 0.01 | 11.96 | -0.01 | 8.51 | 0.02 | 0.66 | -0.003 | 0.14 | 0.01 | 0.27 | -0.02 | 2.89 | -0.004 | 0.27 |
| rs7133378* | 12 | 124,409,502 | DNAH10 | A | G | 0.68 | -0.17 | 17.79 | 0.016 | 8.42 | -0.03 | 44.92 | 0.02 | 20.64 | -0.03 | 1.70 | 0.02 | 0.92 | -0.05 | 5.33 | 0.03 | 5.47 | 0.03 | 8.40 |
| rs1716407* | 12 | 124,515,218 | ZNF664 | A | G | 0.41 | 0.14 | 12.98 | -0.015 | 7.55 | 0.02 | 23.10 | -0.02 | 17.96 | 0.02 | 1.08 | -0.02 | 0.89 | 0.05 | 7.00 | -0.03 | 5.10 | -0.02 | 6.00 |
| rs12454712 | 18 | 60,845,884 | BCL2 | C | T | 0.62 | -0.11 | 7.95 | -0.001 | 0.14 | -0.01 | 15.80 | 0.01 | 14.42 | 0.01 | 0.68 | -0.01 | 0.62 | -0.02 | 1.08 | 0.05 | 11.62 | 0.02 | 3.30 |
| rs56361048† | 19 | 33,885,318 | PEPD | C | T | 0.86 | -0.16 | 9.01 | -0.009 | 1.52 | -0.02 | 19.52 | 0.02 | 17.66 | 0.02 | 0.74 | -0.05 | 3.21 | -0.02 | 0.70 | 0.05 | 6.89 | 0.02 | 1.70 |
| rs7258937† | 19 | 33,938,800 | PEPD | T | C | 0.49 | -0.16 | 17.48 | -0.002 | 0.24 | -0.02 | 24.39 | 0.01 | 12.68 | 0.01 | 0.39 | -0.01 | 0.38 | -0.04 | 3.85 | 0.03 | 5.32 | 0.02 | 3.62 |
| rs190712692 | 19 | 45,425,178 | APOC1 | A | G | 0.95 | -0.24 | 7.93 | NA | NA | -0.07 | 71.47 | -0.15 | >112 | -0.05 | 1.60 | 0.02 | 0.19 | -0.003 | 0.03 | NA | NA | 0.15 | 26.57 |
| rs4821764 | 22 | 38,599,364 | MAFF | A | G | 0.42 | 0.17 | 20.20 | 0.007 | 1.71 | 0.02 | 33.89 | -0.02 | 44.08 | 0.02 | 1.18 | 0.004 | 0.15 | 0.05 | 5.32 | -0.03 | 3.96 | 0.001 | 0.07 |

A1: Effect Allele, A0: Non-effect allele, A1FREQ: Frequency of A1 allele, LOG10P: $-\log_{10}$ (p-value)
\* These SNPs remained GWS when both were included in the model (Table S10).
<sup>†</sup> Only rs7258937 remained GWS when both SNPs were included in the model (Table S10).

#### Autosomal BFP GWAS loci with paradoxical effects on lipids, T2D and CAD

Lipids: Twenty SNPs had paradoxical associations with BFP and HDL (i.e. the direction of effect on BFP and HDL was the same); and 21 had paradoxical associations with BFP and TG (i.e. the direction of effect on BFP was the opposite of effect direction on TG) with 14 having paradoxical associations with both HDL and TG. Of these 14 SNPs, three were associated with WHR adjusted for BMI all with opposite direction of effect on BFP and WHR including rs998584 (Chr6:43,757,896; A>C; *VEGFA*), rs7133378 (Chr12: 124,409,502; A>G; *DNAH10*) and rs1716407 (Chr12:124,515,218; A>G; *ZNF664*). rs998584 was associated with multiple cardiometabolic phenotypes: the BFP increasing allele associated with increased GFAT, reduced VAT and reduced risk of T2D and CAD underscoring the role of fat distribution in cardiometabolic disease in males. rs78058190 (Chr2:219,699,999; A>G; *PRKAG3*) and rs71602277 (Chr4:157,714,979; TA>T; *PDGFC*) were also associated with fat depots with the BFP increasing allele associating with increased GFAT (Table 2, Figure 4A).

**Figure 4:**
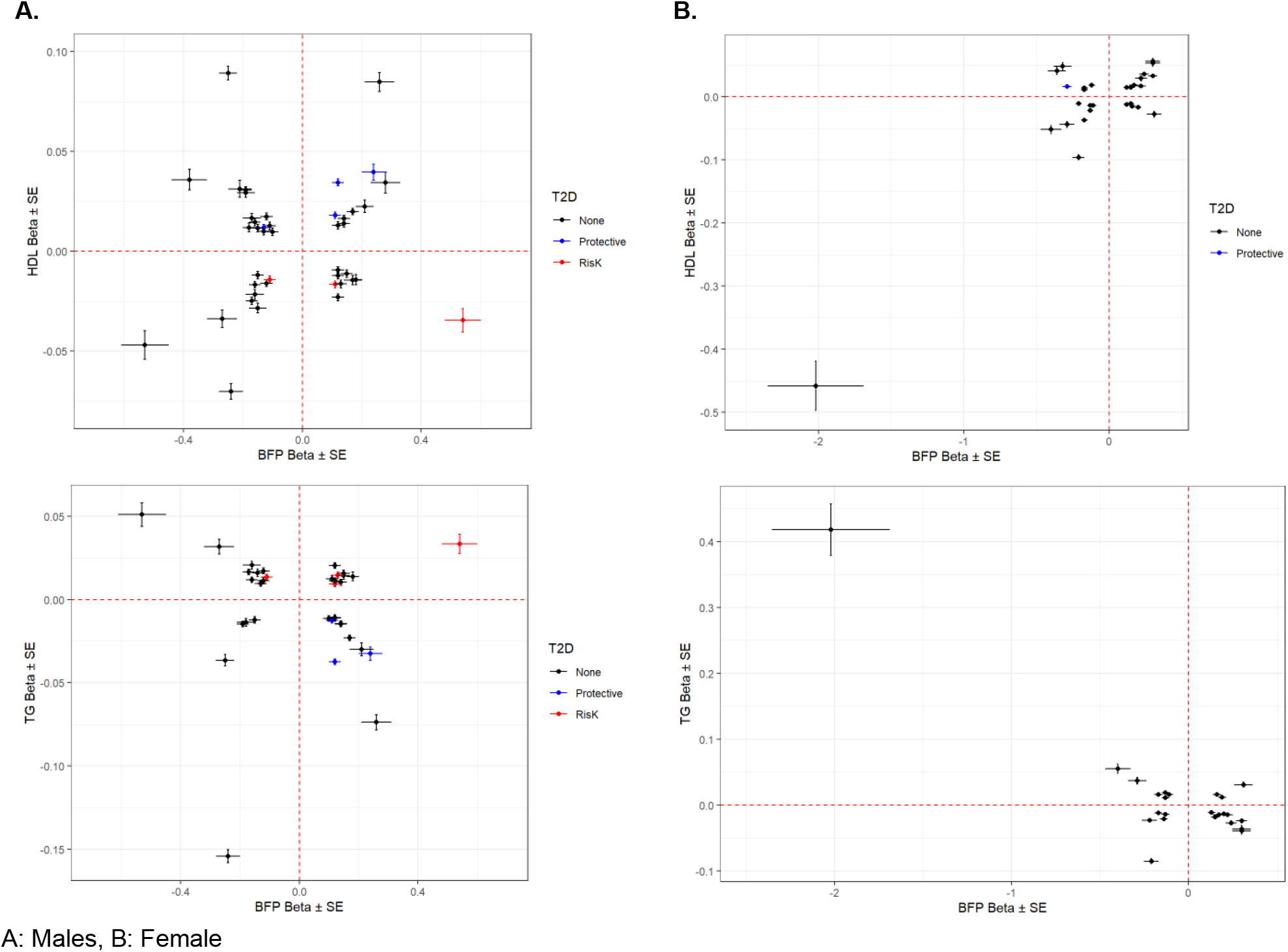
Association of SNPs with HDL and TG vs. BFP The SNPs associated with BFP in each sex that were also associated with HDL (top) and TG (bottom) in their corresponding sex ^24^ were included in the plots. SNPs with risk and protective effect on T2D (both sexes combined) were shown in red and blue, respectively ^22^.

Lipids and T2D: The BFP increasing alleles of rs62271373 (Chr3:150,066,540; A>T; *TSC22D2*), rs9641894 (Chr7:130,465,054; G>T; *KLF14*), and rs12454712 (Chr18:60,845,884; C>T; *BCL2*) were associated with reduced T2D risk, higher HDL and lower TG (Table 2, Figure 4A).

Lipids and CAD: The BFP decreasing allele of rs7133378 (Chr12:124,409,502; A>G; *DNAH10*) was associated with lower HDL and higher TG, and increased risk of CAD (Table 2, Figure 4A).

#### Chr X

545,899 SNPs on Chr X were tested for association with BFP. There were 2,713 GWS SNPs including 22 independent signals (Figure S2A, Males.xlsx File, Sheet F & G). Of these, two SNPs (rs5950969 and rs73505165) have not been associated with SHBG, testosterone or any adiposity related phenotypes previously (Table 1).

#### HLA Imputation

None of the HLA haplotypes reached GWS threshold with top associated haplotype being *HLA-DRB4*0103* (β (SE) = 0.09 (0.02), p = 5.61E-5) (Males.xlsx File, Sheet H).

#### MR

The MR Egger method did not show any significant causal effect for BFP on T2D (p = 0.58), CAD (p = 0.95), HDL (p = 0.63) or TG (p = 0.58). Some but not all of the other methods showed some significant associations but overall we did not find any reliable evidence that increased BFP increases risk of T2D, CAD or TG levels, or decreases HDL; and there was highly significant evidence for heterogeneity (p < 2E-110). There was no evidence for horizontal pleiotropy (p >0.05) (Males.xlsx File, Sheet K).

Single SNP MR results demonstrated three groups of SNPs suggesting positive, negative and no significant causal effect of BFP on the four outcomes consistent with high levels of heterogeneity.

Leaving out rs35198068 within *TCF7L2* a known locus for T2D which has opposite effects on BFP and T2D led to significance of MR analysis. None of the SNPs made significant difference in MR analysis results of CAD, HDL or TG in the leave-one-out analysis (Males_MR_Plots.pdf).

### Females

#### Autosomal GWAS

154,337 females were included in the analysis (Table S3). BFP was normally distributed with mean (SD) of 36.7 (6.9) % (Figure S1B). Both mean and variance of BFP were significantly higher in females compared to males (p < 2.2E-16). In the multivariable analysis, age and testosterone were associated with higher BFP in females whereas albumin and SHBG were associated with lower BFP. Age, albumin, SHBG, testosterone, and their quadratic terms together explained 23% of variation in BFP. SHBG and its quadratic term explained 18% of variation BFP (Table S4 & S5).

23,640,526 SNPs on Chr1-22 were included in the GWAS (GC lambda = 1.19) (Figure 2B). 13,612 SNPs were associated with BFP at GWS level (Females.xlsx File, Sheet A). There were 334 independent GWS SNPs including 74 SNPs in *SHBG* (Females.xlsx File, Sheet B). One-hundred and nineteen SNPs were common with males (Females.xlsx File, Sheet C). Six SNPs in *SHBG* region were missing in Ruth et al analysis; but the rest were all associated with SHBG and/or testosterone except for an indel (Chr17:7200613; CAA>C) which was not associated with SHBG (p = 2.00E-5) or testosterone (p = 0.47). Of 260 SNPs in non-*SHBG* loci, 174 (only 38 common with males) were not associated with SHBG or testosterone (Females.xlsx File, Sheet D). After, further clumping of these 174 SNPs with r^2^ < 0.001, 140 SNP were left, and they explained 2.60% of the variation in BFP in females (Females.xlsx File, Sheet J).

#### Novel autosomal BFP loci

Of these 174 SNPs, 169 were not associated with BFP in prior published GWAS ^12^ and 104 were not identified by Neale’s round 2 GWAS results of BFP in females (Females.xlsx File, Sheet I). Ten of these SNPs have not been associated with any adiposity related phenotypes (Table 1), fat depots (Table S6), HDL, TG, T2D or CAD previously (Females.xlsx File, Sheet E).

#### Association of autosomal BFP GWAS loci with cardiometabolic Phenotypes

Of 174 autosomal SNPs associated with BFP in females, the majority (132, 75%) did not associate with cardiometabolic phenotypes. Forty-two SNPs were associated with lipid levels, T2D, CAD or fat depots with some associated with multiple traits: HDL: 29, TG: 24, T2D: 4, CAD: 3, GFAT: 5, VAT: 1, and ASAT: 1 (Figure 3, Females.xlsx File, Sheet E).

Seventeen SNPs were associated with both HDL and TG. Of these, 16 had consistent effect on HDL and TG (i.e. the allele associated with higher HDL was associated with lower TG and vice versa). However, the C allele of rs7412 (Chr19: 45,412,079; T>C, a missense variant within *APOE* which defines the Ɛ2 haplotype) which was associated with lower BFP (β (SE) = -0.21 (0.04), -log_10_(p) = 7.45) was associated with lower HDL (β (SE) = -0.096 (0.004), p = 9.3E-163) and lower TG (β (SE) = -0.085 (0.004), p = 9.65E-128). rs7412 is in high LD with rs190712692 (D’ = 1, R^2^ = 0.86, based on CEU population in the 1000 Genomes Project v3) which had similar effect on HDL and TG from males GWAS. Similar to rs190712692, rs7412 was associated with increased risk of CAD (β (SE) = 0.15 (0.01), p = 1.04E-54) (Table 3).

**Table 3:**
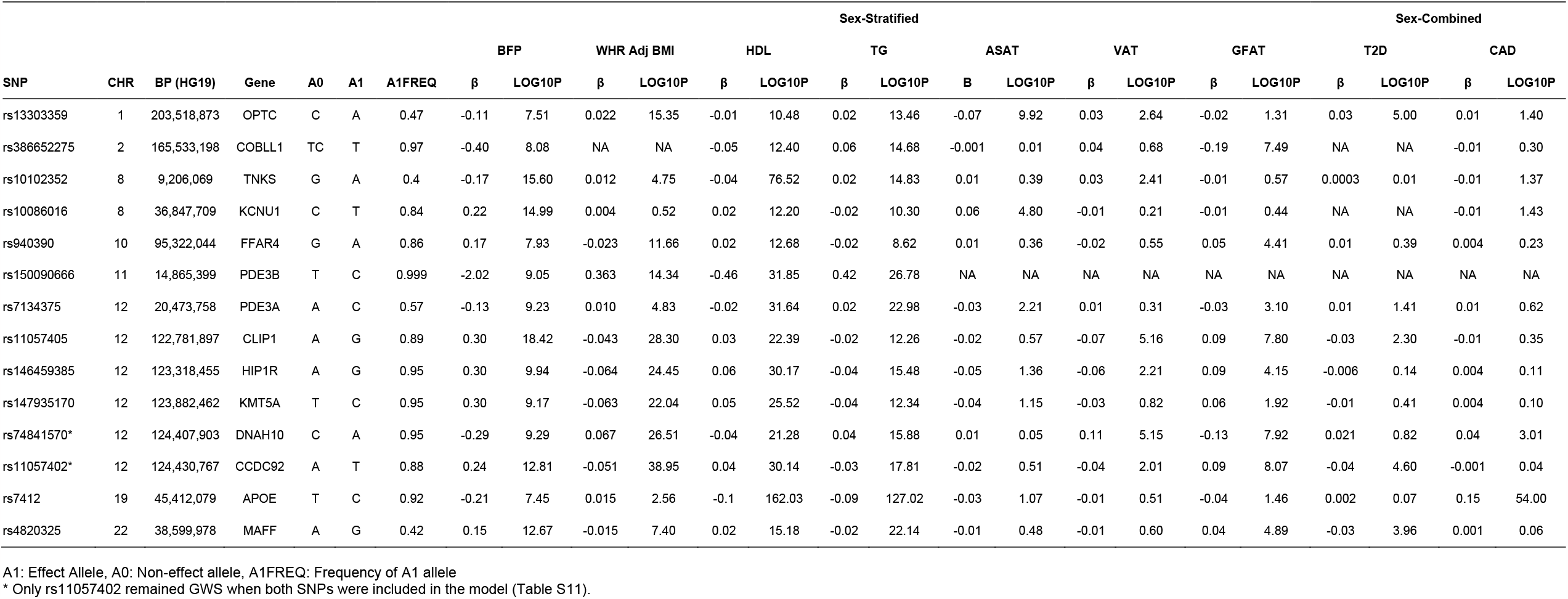
BFP GWAS loci in females having paradoxical effect on HDL and TG

#### Autosomal BFP GWAS loci with paradoxical effects on cardiometabolic phenotypes

Eighteen SNPs had paradoxical associations with BFP and HDL (i.e. the direction of effect on BFP and HDL was the same); and 16 had paradoxical associations with BFP and TG (i.e. the direction of effect on BFP was the opposite of effect direction on TG) with 13 having paradoxical associations with both HDL and TG. This included a rare rs150090666 (Chr11:14,865,399, Freq = 0.001) stop-gain (**C**GA>**T**GA, Arg861>*) variant within *PDE3B* (NP_001350499.1) with a large effect on BFP (β (SE) = 2.02 (0.33), -log_10_(p) = 9.05) as well as HDL (β (SE) = 0. 458 (0.039), p = 1.40E-32) and TG (β (SE) = -0.418 (0.039), p = 1.66E-27) (Figure 4B). Rare coding variants in *PDE3B* have been associated with BMI adjusted WHR in sex-combined analyses previously ^10,28^. Of the 13 SNPs with paradoxical effect on HDL and TG, nine were associated with WHR adjusted for BMI all with opposite direction of effect on BFP and WHR. Four SNPs including rs386652275 (Chr2:165,533,198; TC>T; *COBLL1*), rs11057405 (Chr12:122,781,897; A>G; *CLIP1*), rs74841570 (Chr12:124,407,903; C>A; *DNAH10*), and rs11057402 (Chr12:124,430,767; A>T; *CCDC92*) were associated with GFAT. For all 4 SNPs, the BFP increasing allele associated with increased GFAT and improved lipids (higher HDL and lower TG).

The BFP lowering allele of rs13303359 (Chr1: 203,518,873, C>A; *OPTC*) was associated with lower ASAT, lower HDL and higher TG (Table 3, Figure 4B).

#### Chr X

930,172 SNPs on Chr X were tested for association with BFP. There were 468 GWS SNPs all in one locus (rs12011976: Chr23:109,836,588) (Figure S2B, Females.xlsx File, Sheet F & G). This locus has been associated with SHBG, testosterone and adiposity related phenotypes previously. It was also associated with BFP in males.

#### HLA Imputation

None of the HLA haplotypes reached GWS threshold with top associated haplotype being *HLA-B*4403* (β (SE) = -0.17 (0.05), p = 2.39E-4) (Females.xlsx File, Sheet H).

#### MR

The MR Egger method showed significant positive causal effect of BFP on HDL (β (SE) = 0.047 (0.022), p = 0.028) with significant evidence for horizontal pleiotropy (p = 0.017) and heterogeneity (p < 2E-110). However, this result was not supported by the other four MR methods. Both weighted median (β (SE) = -0.013 (0.003), p = 1.54E-5) and simple mode (β (SE) = -0.038 (0.010), p = 3.75E-4) showed that BFP has negative causal effect on HDL. IVW and weighted mode did not show any significant causal effects but the direction of effect was consistent with weighted median and simple mode methods. The MR Egger method did not show any significant causal effect for BFP on T2D (p = 0.65), CAD (p = 0.86) or TG (p = 0.066). Some, but not all, of the other methods showed significant associations but collectively we did not find any reliable evidence that increased BFP increases risk of T2D, CAD or TG levels; and there was highly significant evidence for heterogeneity (p < 2E-75). There was no evidence for horizontal pleiotropy (p >0.05) (Females.xlsx File, Sheet K).

Similar to males, single SNP MR results demonstrated three groups of SNPs suggesting positive, negative and no significant causal effect of BFP on the four outcomes consistent with high levels of heterogeneity.

None of the SNPs made significant difference in MR analysis results of T2D, CAD, HDL or TG in the leave-one-out analysis (Females_MR_Plots.pdf).

### SNP x Sex Interaction GWAS

312,274 subjects and 28,442,141 SNPs were included in the GWAS (GC lambda = 1.03). Only 25 SNPs in 4 loci reached the GWS threshold: rs754823863, rs16885587, rs5030937 and rs55745760 (Figure 1, Table S7). rs754823863 and rs55745760 were not GWS associated with BFP in males or females whereas rs16885587 and rs5030937 were associated with BFP in only females (Table 4).

**Table 4:** SNPs interacting with sex affecting BFP

| CHR | BP (HG19) | SNP | Gene | A0 | A1 | All (SNP x Sex Interaction) |  |  |  |  | Males |  |  |  |  | Females |  |  |  |  |
| --- | --- | --- | --- | --- | --- | --- | --- | --- | --- | --- | --- | --- | --- | --- | --- | --- | --- | --- | --- | --- |
|  |  |  |  |  |  | A1FREQ | INFO | BETA | SE | LOG10P | A1FREQ | INFO | BETA | SE | LOG10P | A1FREQ | INFO | BETA | SE | LOG10P |
| 4 | 62,634,106 | rs754823863 | ADGRL3 | T | C | 0.999 | 0.73 | 3.60 | 0.65 | 7.52 | 0.999 | 0.73 | -1.56 | 0.44 | 3.41 | 0.999 | 0.73 | 1.96 | 0.48 | 4.33 |
| 8 | 36,794,458 | rs16885587 | KCNU1 | C | T | 0.872 | 0.98 | 0.23 | 0.04 | 7.54 | 0.872 | 0.98 | 0.01 | 0.03 | 0.09 | 0.872 | 0.98 | 0.22 | 0.03 | 12.07 |
| 10 | 70,975,897 | rs5030937 | HKDC1 | T | C | 0.308 | 0.99 | -0.17 | 0.03 | 7.82 | 0.308 | 0.99 | 0.05 | 0.02 | 1.89 | 0.308 | 0.99 | -0.10 | 0.02 | 5.45 |
| 17 | 7,568,925 | rs55745760 | TP53 | T | C | 0.597 | 0.96 | -0.17 | 0.03 | 8.15 | 0.597 | 0.96 | 0.01 | 0.02 | 0.11 | 0.596 | 0.96 | -0.16 | 0.02 | 12.97 |

## DISCUSSION

Sex and sex-hormones are significant determinants of adiposity. We therefore performed sex-stratified GWAS of BFP in the UK Biobank including SHBG and testosterone in the model. Our data suggests that 1. This approach increases the power to detect BFP associated loci. 2. Despite adjustment for testosterone and SHBG, the majority of BFP loci do not overlap between sexes 3. Identified loci, which do not associate with testosterone and SHBG, generally do not appear to have significant deleterious cardiometabolic effects.

### Genetic determinants of BFP

We identified 193 autosomal loci in males and 174 autosomal loci in females associated with BFP a significant increase from the 12 loci identified in a previously published BFP GWAS ^12^. The identified loci explained 3.35% and 2.60% of the variation in BFP in males and females, respectively; whereas 12 previously identified loci explained only 0.57% of the variation in BFP ^12^. Of these, 94 loci in males and 104 loci in females were not identified in unpublished analyses by the Neale lab. Seven loci in males (including 2 on Chr X) and 10 in females have not been associated with any adiposity related/cardiometabolic traits previously. Despite adjustment for SHBG and testosterone, a minority of loci (38 loci) associated with BFP in both sexes, underscoring the differential genetic regulators of BFP in each sex.

Eight out of twelve previously reported loci for BFP ^12^ were associated with BFP in both males and females in our analyses. Another locus, *IRS1* (insulin receptor substrate 1), was only associated with BFP in males. The other three loci were not replicated in males or females: *SPRY2, IGF2BP1* and *CRTC1*. There were multiple independent signals in *FTO* locus for males and in *COBLL1* locus for females (Table S8).

### Variation in BFP explained by other factors

Age and its quadratic term explained only ∼3% of BFP variation in both males and females. Adding albumin, SHBG and testosterone, and their quadratic terms to the model significantly improved the proportion of variation explained to 13% and 23% in males and females, respectively. Both mean and variance of BFP were significantly higher in females compared to males. SHBG was associated with lower BFP in both sexes whereas testosterone was associated with lower BFP in males but it was associated with higher BFP in females consistent with previous data ^18^. SHBG explained a large proportion of BFP variation specially in females; whereas testosterone significantly improved the predictive power of the model in males.

### Association of BFP loci with cardiometabolic disease

Obesity generally increases cardiometabolic risk. Intriguingly, the majority of identified BFP loci in males and females did not associate with lipid levels or cardiometabolic diseases such as T2D (13 out of 193 and 4 out of 174 in males and females, respectively) or CAD (5 and 3 in males and females, respectively). A number of BFP increasing alleles, were paradoxically associated with higher HDL and lower TG, and in some cases reduced risk of T2D and CAD. They were also often were associated with lower WHR and in some cases increased GFAT. MR analysis also did not provide any convincing evidence of causal effect of BFP on HDL, TG, T2D or CAD; and showed highly significant levels of heterogeneity. Consistent with previous data, these finding indicate that higher BFP, in the absence of deleterious fat distribution, likely does not increase cardiometabolic risk. It is also likely indicative of the contribution of testosterone/SHBG, which are not associated with these variants, to cardiometabolic phenotypes ^13,14^.

Previous genetic studies have not reported adverse metabolic effects of reduced ASAT ^10^. At the *OPTC* locus the BFP lowering allele associated with reduced ASAT, lower HDL and higher TG. Whether the metabolic effects are due to reduced ASAT is not established. At another locus on Chr19 between *CEBPG* and *CEBPA*, a variant associated with higher BFP and VAT in females but not males was also associated with higher HDL levels (in both sexes) potentially suggesting the HDL increase is independent of fat distribution. At *APOC1* and *APOE* locus, we observed variants which were associated with both lower HDL and lower TG. However, these variants were associated with higher risk of CAD. This locus has been associated with total and LDL cholesterol as well as statin use, which may influence the association with CAD ^29,30^.

### Gene expression analyses

We investigated whether novel identified SNPs affected gene expression in different tissues based on the Genotype-Tissue Expression (GTEx) project v8 ^31,32^ with a more specific focus in visceral adipose tissue. We report that the novel loci did not affect expression of the genes that SNPs are located in or the nearest genes to the SNP (Table S9). For example, rs2941584 (Chr2: 54,881,621) is an intronic SNP within *SPTBN1* and is an expression quantitative trait loci (eQTL) for another gene in its vicinity, *EML6* which might be a more relevant gene as variants within *EML6* have been associated with extreme obesity previously ^33^. Another example is rs4962671 (Chr10:126,305,434) an intergenic SNP between *FAM53B* and *LHPP* which is an eQTL in visceral adipose tissue for *METTL10* (Chr10:126,447,406-126,480,439) ∼142 kb away. rs56385874 (Chr19: 47,282,245) is also an intronic SNP within *SLC1A5* but it is eQTL in visceral adipose tissue for two other genes: *FKRP* and *PRKD2* 20 and 62 kb away from the SNP, respectively. This data highlights candidate genes for further functional studies. **Limitations:** Our study had some limitations. The participation rate in UK Biobank at baseline was 5.5% of invitees, with higher participation rate in females than males, and participants were less likely to be obese and healthier than the general population ^34^. Mean and variance of BFP were different in males and females. Therefore, for a SNPs with similar effect size in both sexes, larger sample size was required to detect the association in females than males. For example, the required sample size (power >80% & α = 0.05) to detect association of a SNP with MAF of 20% and effect estimate of 1 was 879 and 1,198 in males and females, respectively. The UK Biobank age ranged between 40-70 years and 72% of the females have had menopause at baseline (UDI 2724-0.0). We did not include estradiol as a covariate in the model for females because the majority of them had the values that were either above or below the detection limit ^35^. These could result in biased estimates. ASAT, VAT, GFAT sample sizes were relatively small, therefore low statistical power could produce false negative results.

### Conclusion

By undertaking sex-stratified analyses adjusted for SHBG and testosterone, we have identified several novel BFP associated loci with limited overlap between sexes. Further, the majority of BFP increasing alleles at identified loci did not associate with adverse cardiometabolic parameters. MR analyses using these BFP associated loci did not provide convincing evidence that that BFP has deleterious cardiometabolic effects which likely underscores the contribution of SHBG and testosterone to fat distribution and cardiometabolic traits.

## Supporting information

Results Regarding BFP GWAS in Males

Plots regarding MR Analysis in Females

Results Regarding BFP GWAS in Males

Plots regarding MR Analysis in Males

## Data Availability

The summary stats for BFP GWAS in males and females are available on MY.LOCUSZOOM.ORG:
https://my.locuszoom.org/gwas/915413/?token=afa2e57d233c48ff972be83bf4360881
https://my.locuszoom.org/gwas/204024/?token=0fb3b984ef1f42a796a3b62de51b5daa
They also will become available on GWAS catalogue upon publication.

## ACKNOWLEDGEMENT

This research has been conducted using the UK Biobank Resource under Application Number 48873, and is supported by CIHR (Grant #175041) and Heart & Stroke Foundation of Canada (Grant # G-21-0031489).

The Genotype-Tissue Expression (GTEx) Project was supported by the Common Fund of the Office of the Director of the National Institutes of Health, and by NCI, NHGRI, NHLBI, NIDA, NIMH, and NINDS. The data used for the analyses described in this manuscript were obtained from the GTEx Portal on 01/01/23.

## DATA AVAILABILITY

The summary stats for BFP GWAS in males and females are available on MY.LOCUSZOOM.ORG:

https://my.locuszoom.org/gwas/915413/?token=afa2e57d233c48ff972be83bf4360881

https://my.locuszoom.org/gwas/204024/?token=0fb3b984ef1f42a796a3b62de51b5daa

They also will become available on GWAS catalogue upon publication.

