## Supplementary material for "Sex-stratified GWAS of Body Fat Percentage after Adjusting for Testosterone and SHBG in the UK Biobank": Plots regarding MR Analysis in Females

## 1- T2D

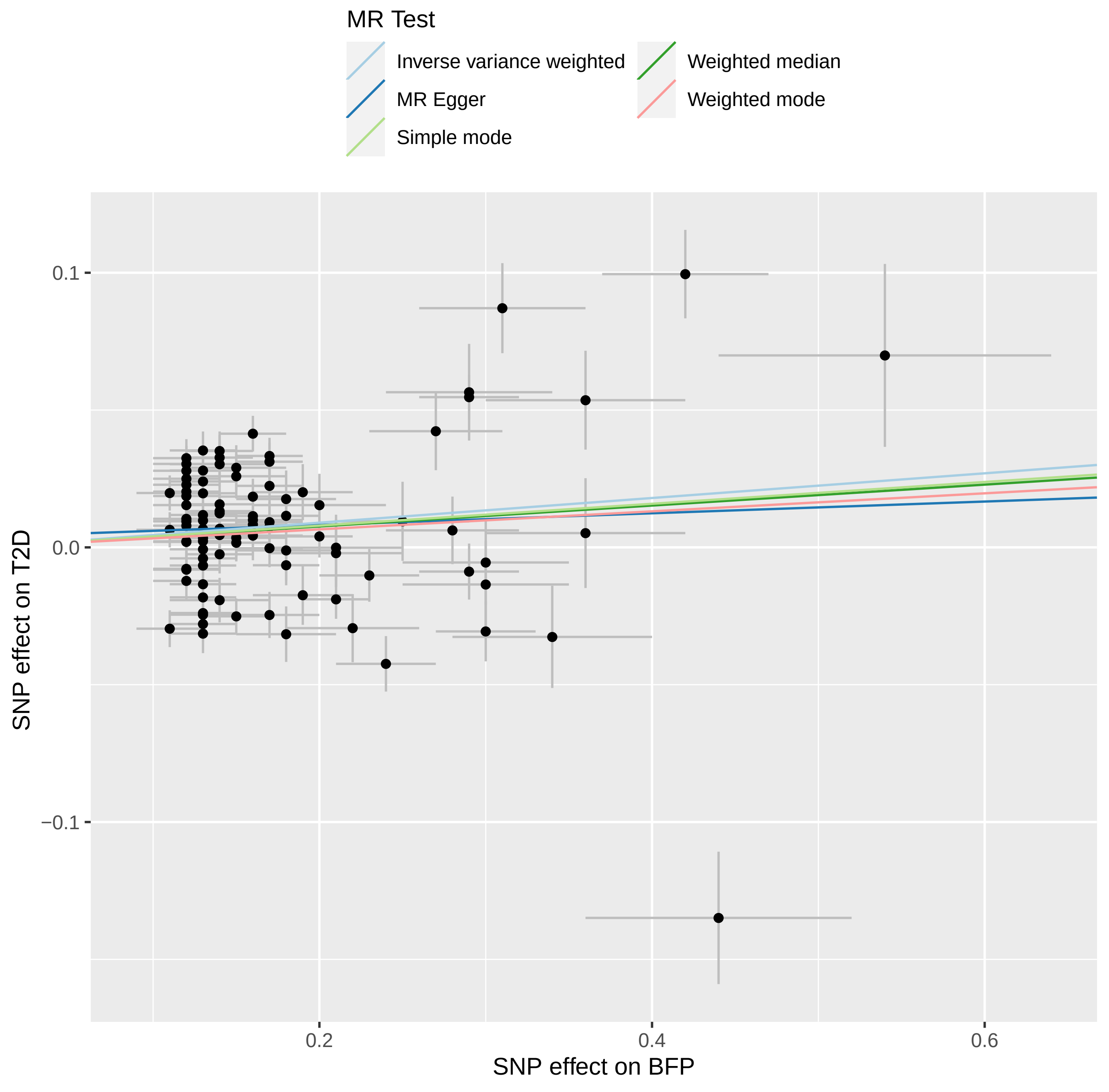

MR Test
Inverse variance weighted
MR Egger
Simple mode
Weighted median
Weighted mode
0.1
0.0
−0.1
0.2
0.4
0.6
SNP effect on T2D
SNP effect on BFP

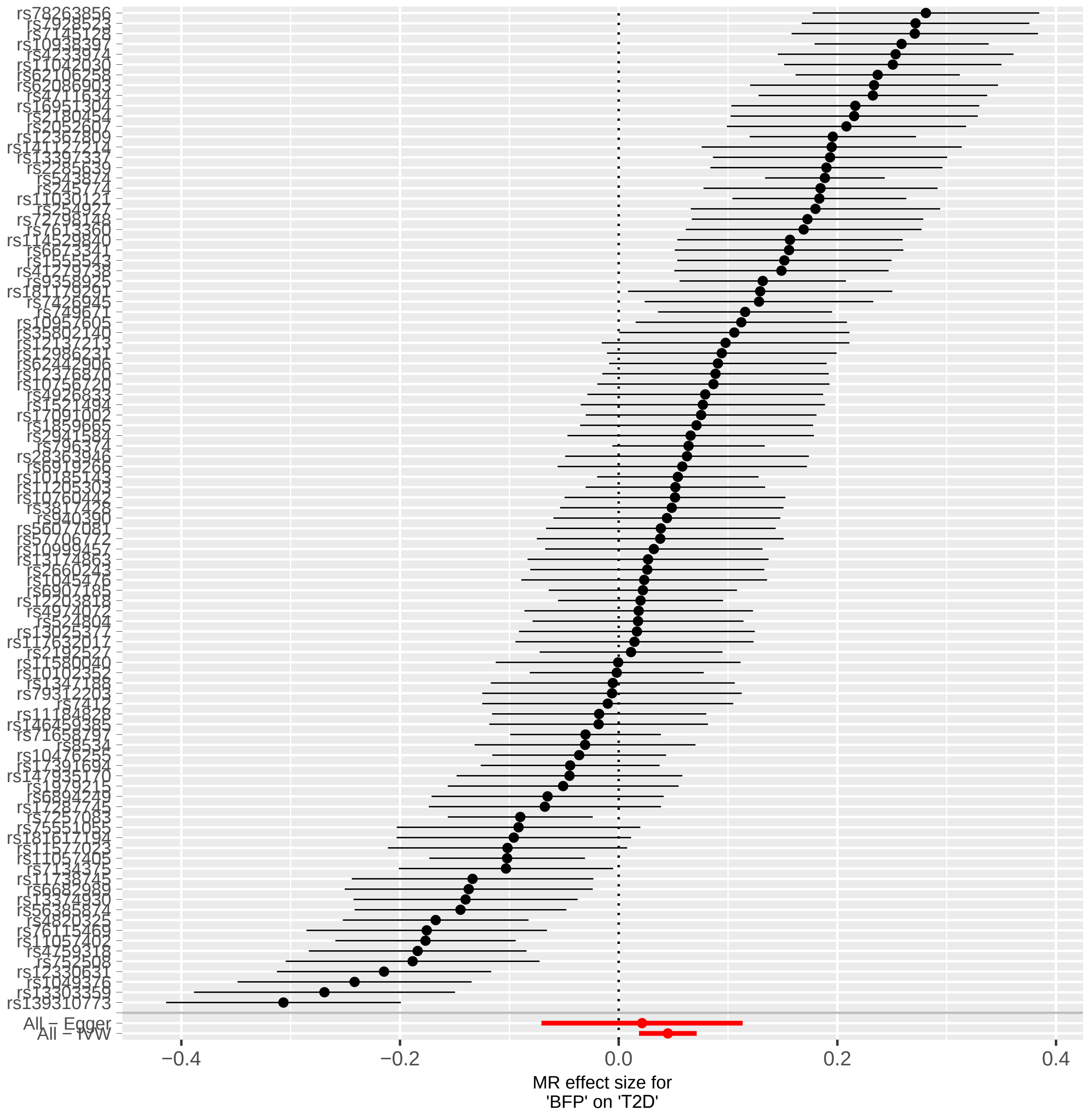

rs78263856
rs139310773
All – Egger
All – IVW
−0.4
−0.2
0.0
0.2
0.4
MR effect size for
'BFP' on 'T2D'

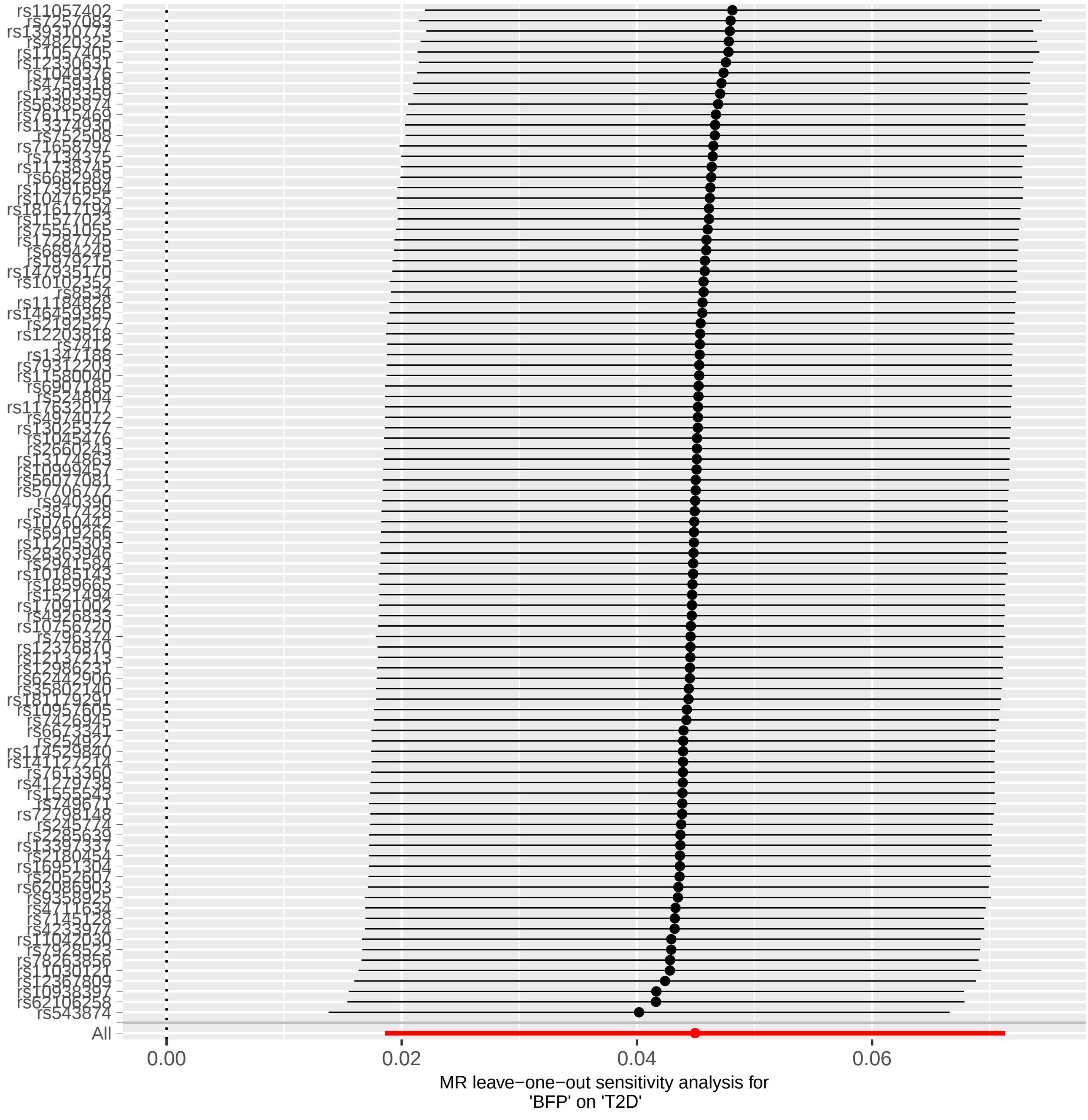
All
0.00
0.02
0.04
0.06
MR leave−one−out sensitivity analysis for
'BFP' on 'T2D'

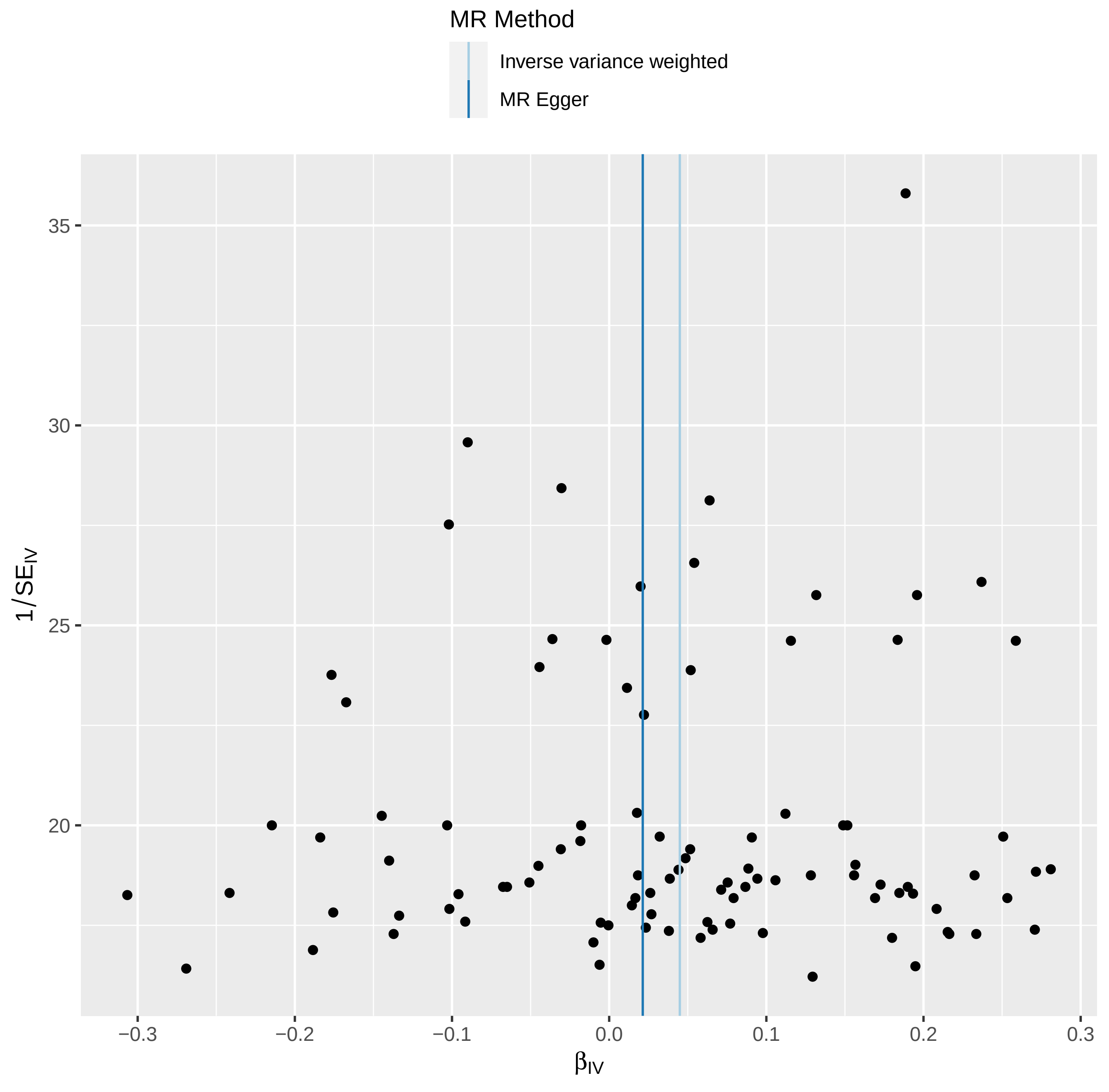

MR Method
Inverse variance weighted
MR Egger
35
30
25
20
1/SE_IV
−0.3
−0.2
−0.1
0.0
0.1
0.2
0.3
β_IV

### 2- CAD

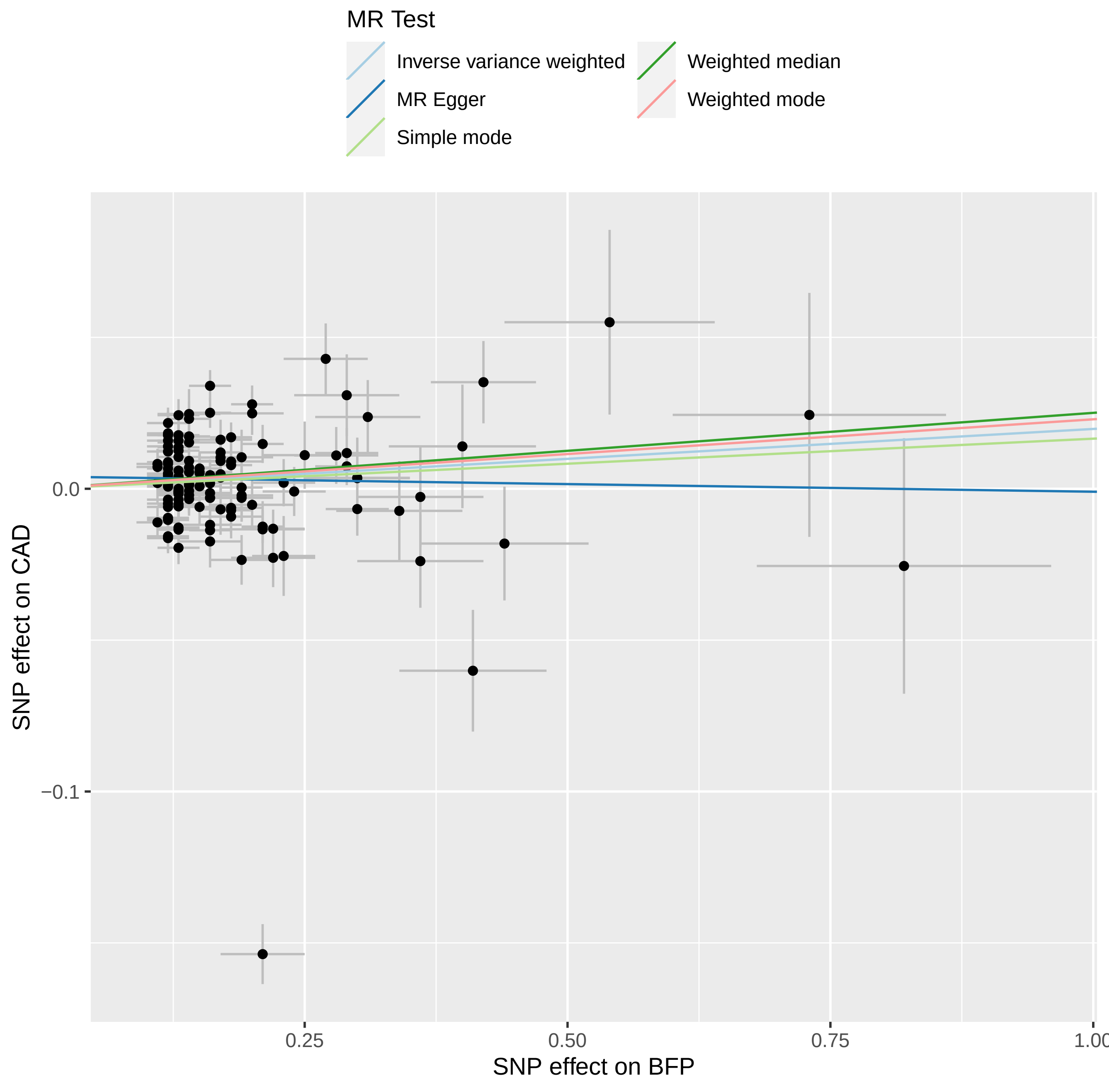
MR Test
Inverse variance weighted
MR Egger
Simple mode
Weighted median
Weighted mode
0.0
−0.1
SNP effect on CAD
0.25
0.50
0.75
1.00
SNP effect on BFP

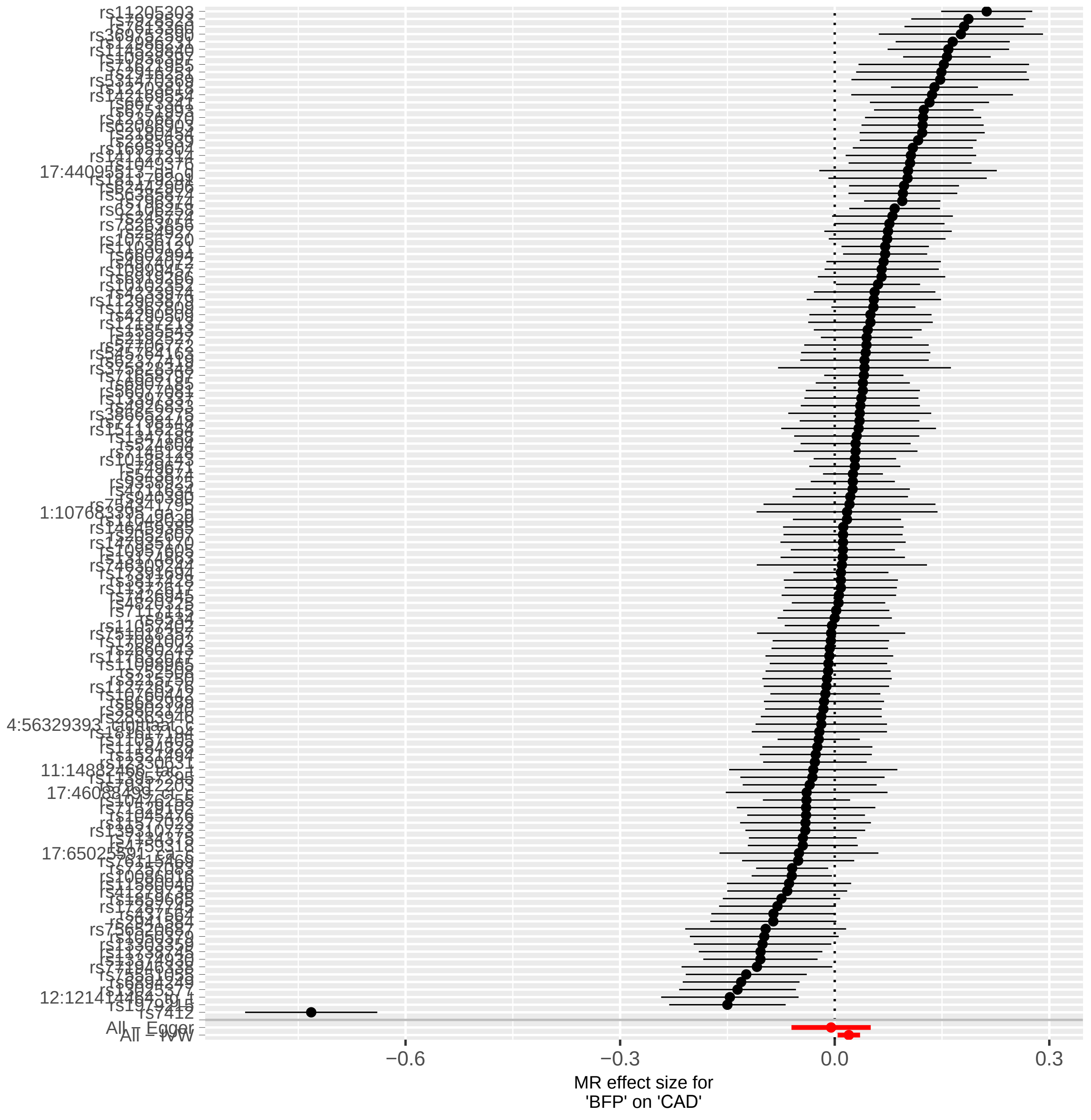
rs11205303
rs7412
All - Egger
All - IVW
−0.6
−0.3
0.0
0.3
MR effect size for
'BFP' on 'CAD'

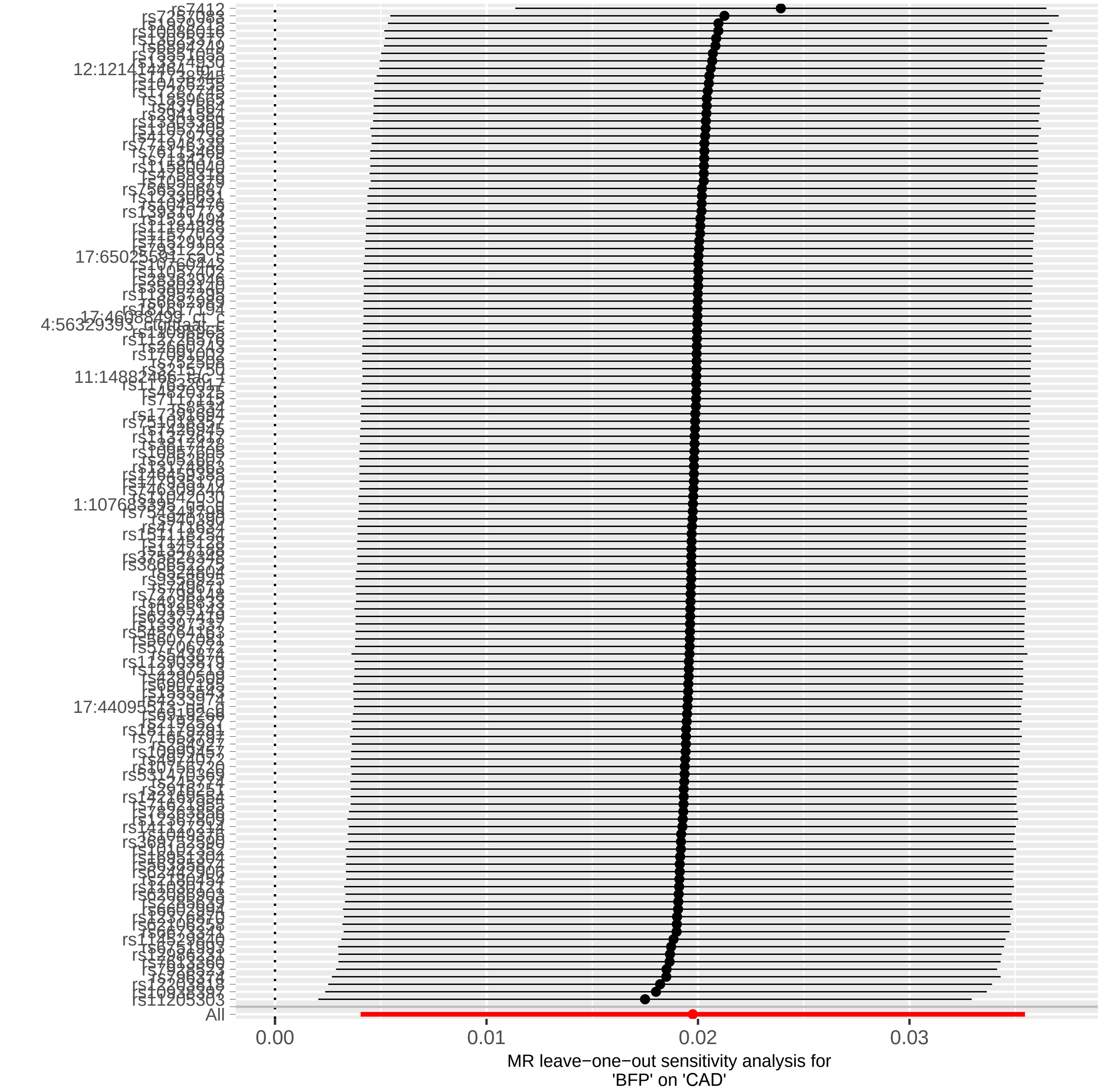

rs7412
rs11205303
All
0.00
0.01
0.02
0.03
MR leave−one−out sensitivity analysis for
'BFP' on 'CAD'

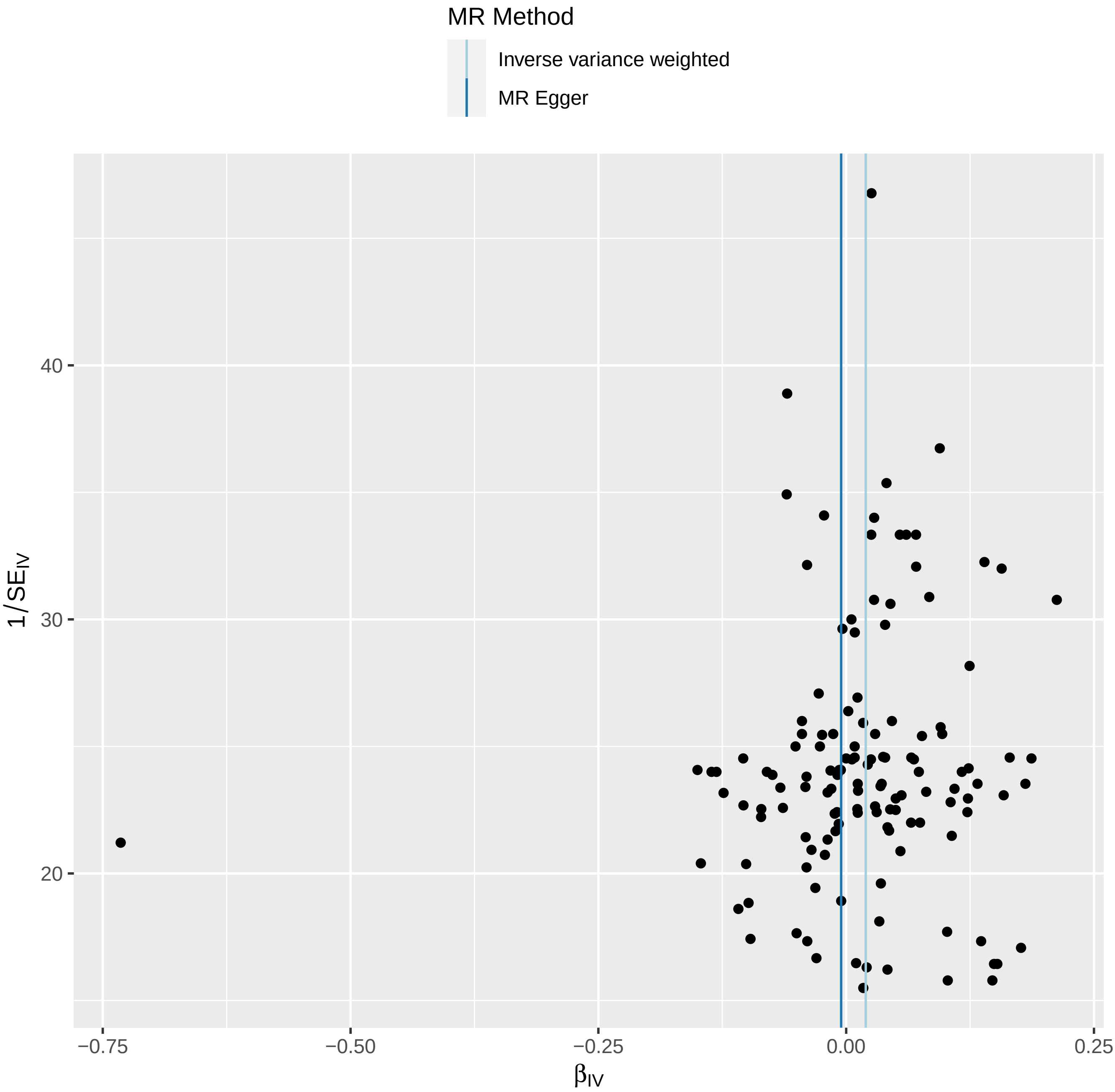

MR Method
Inverse variance weighted
MR Egger
40
30
20
$1/SE_{IV}$
−0.75
−0.50
−0.25
0.00
0.25
$\beta_{IV}$

### 3- HDL

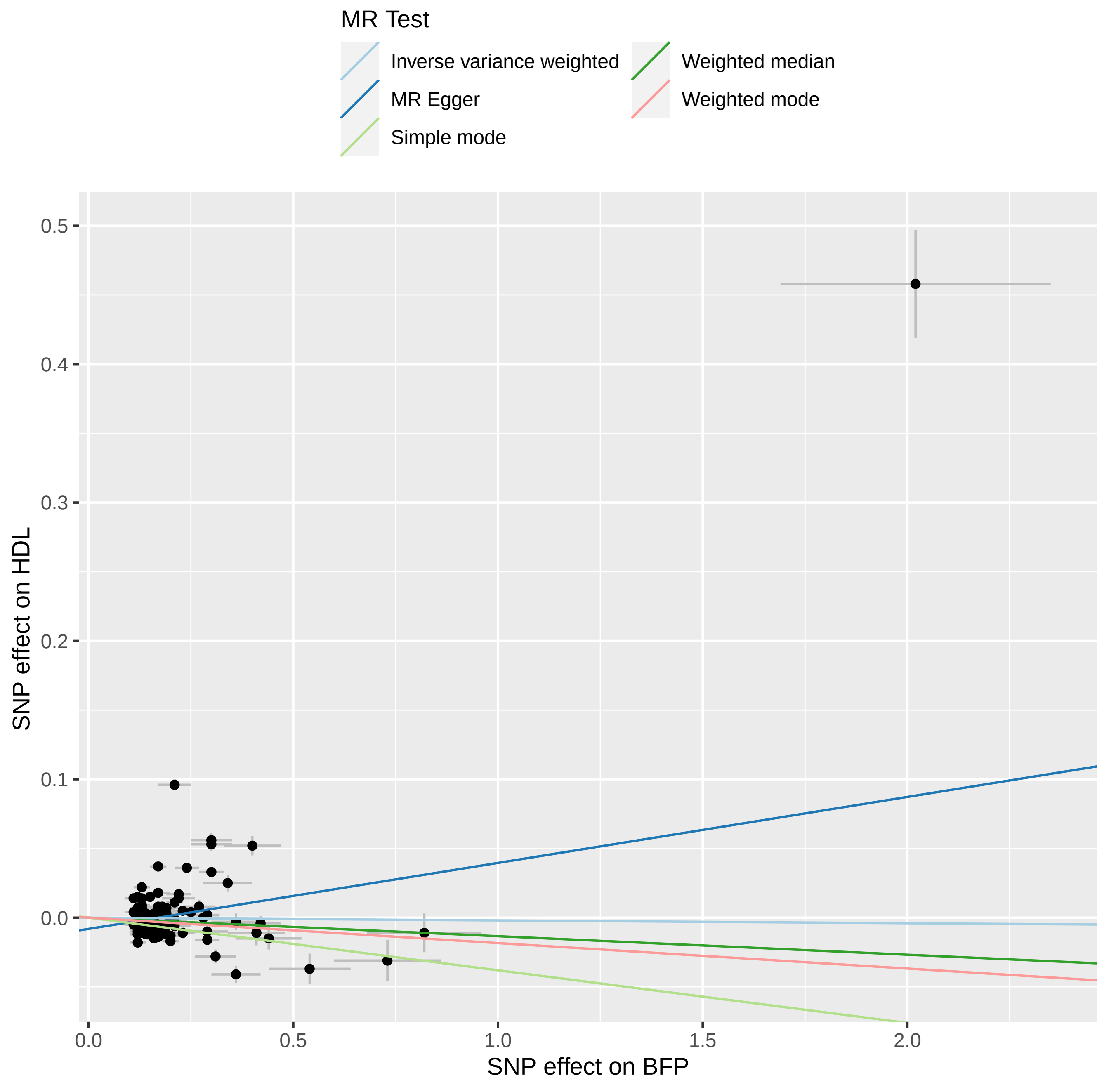
MR Test
Inverse variance weighted
MR Egger
Simple mode
Weighted median
Weighted mode
SNP effect on HDL
0.5
0.4
0.3
0.2
0.1
0.0
0.0
0.5
1.0
1.5
2.0
SNP effect on BFP

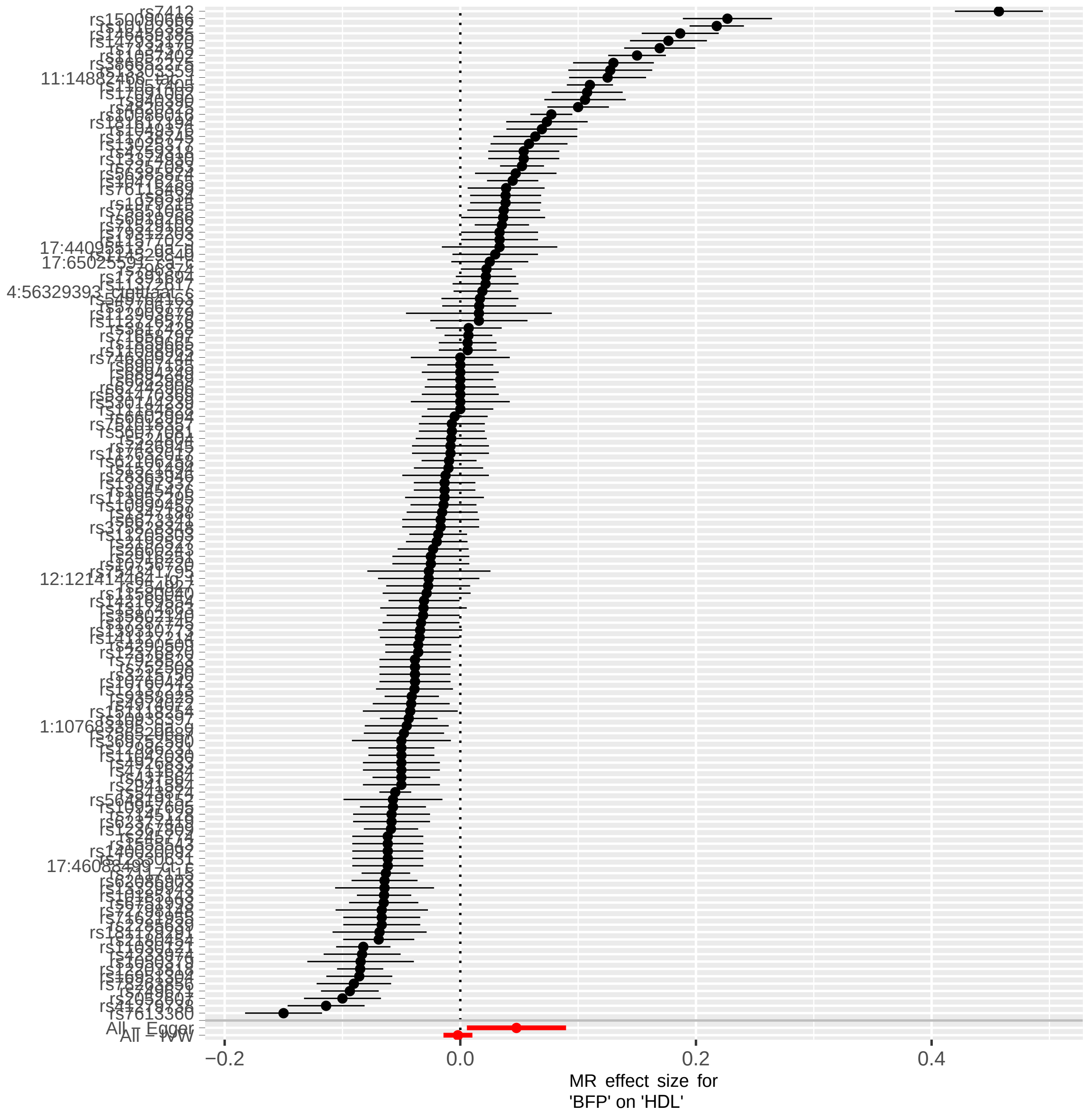
rs7412
rs7613360
All - Egger
All - IVW
−0.2
0.0
0.2
0.4
MR effect size for
'BFP' on 'HDL'

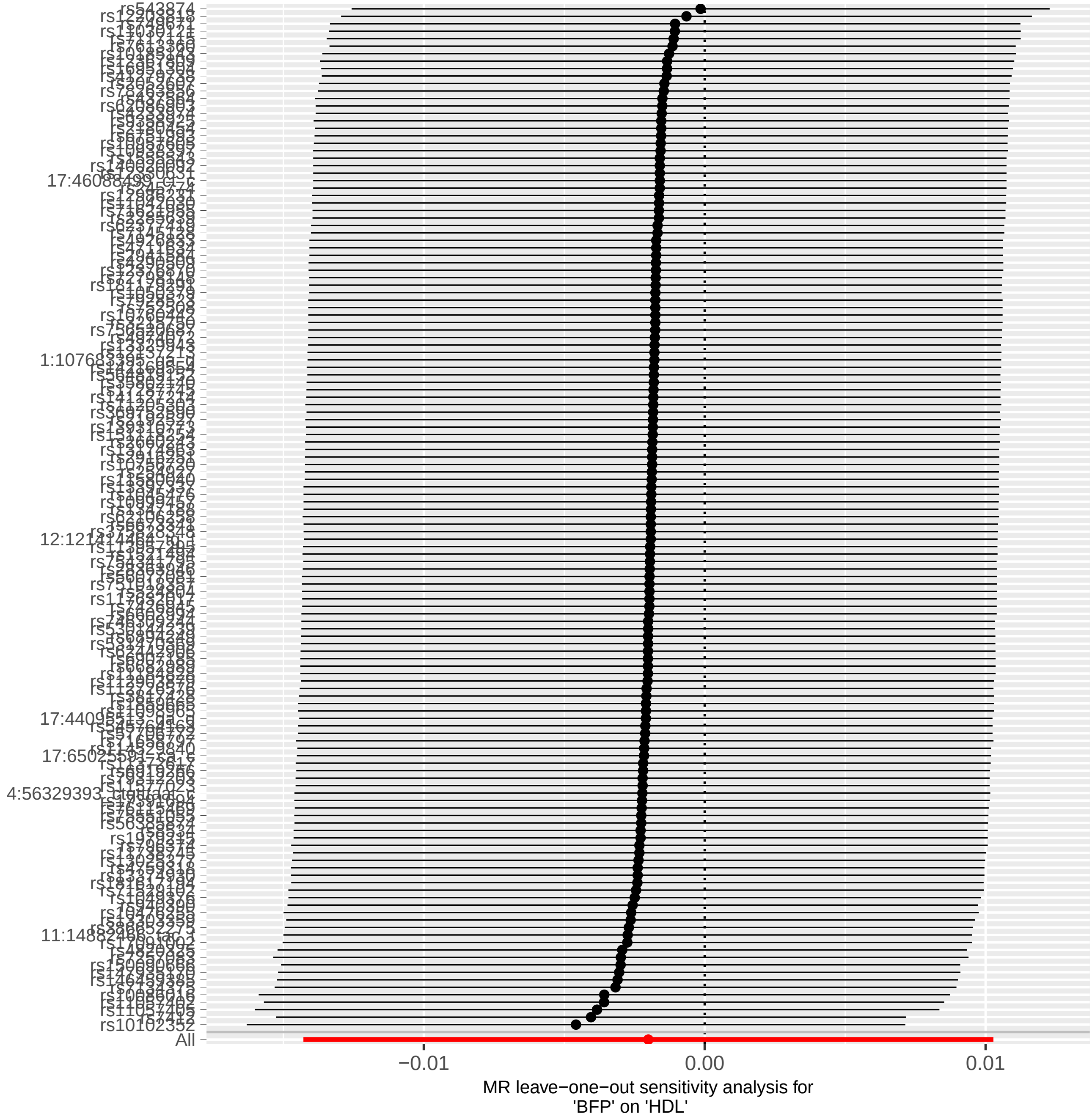
rs543874
rs10102352
All
−0.01
0.00
0.01
MR leave−one−out sensitivity analysis for
'BFP' on 'HDL'

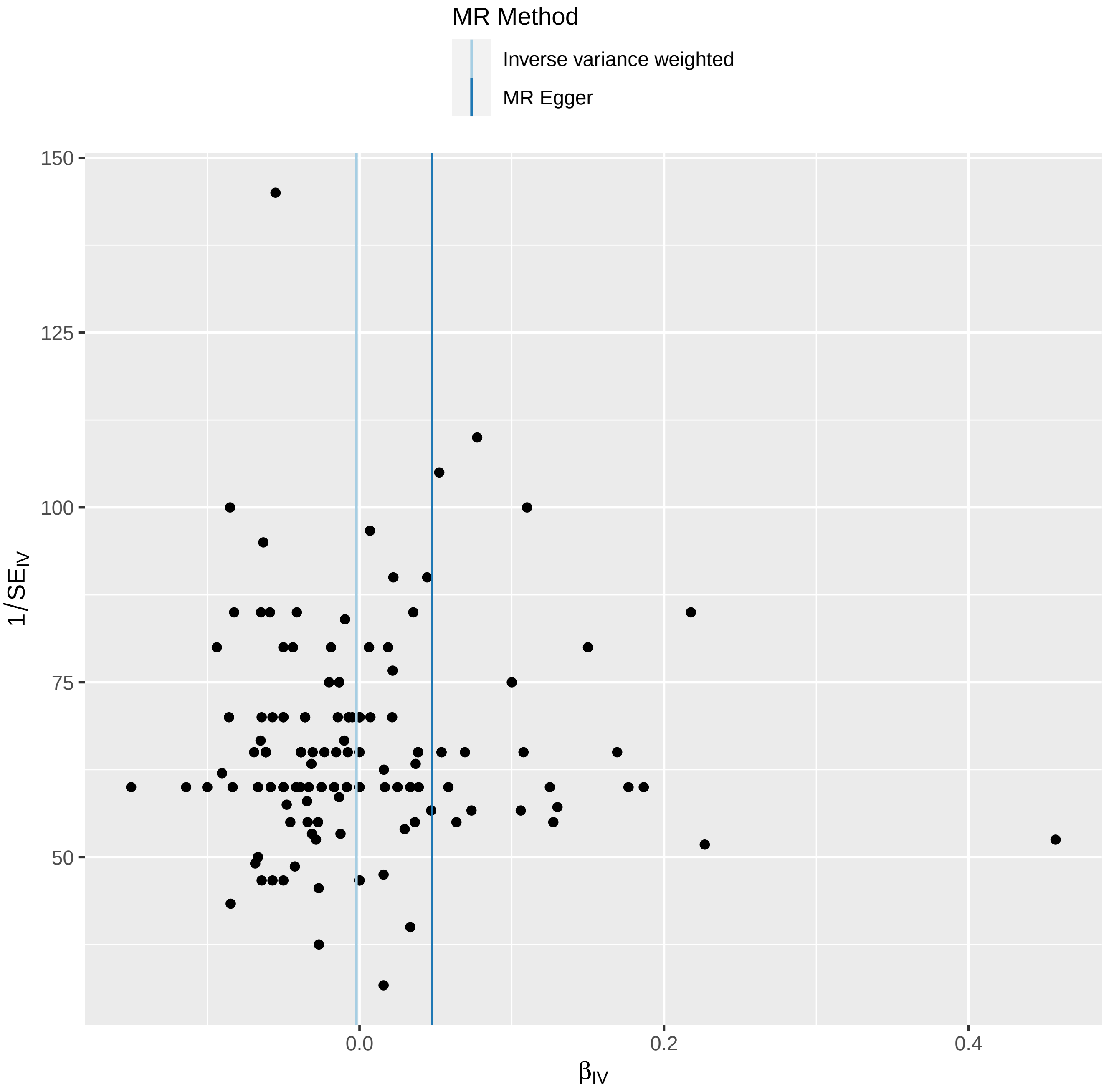
MR Method
Inverse variance weighted
MR Egger
150
125
100
75
50
0.0
0.2
0.4
$\beta_{IV}$
$1/SE_{IV}$

# 4- TG

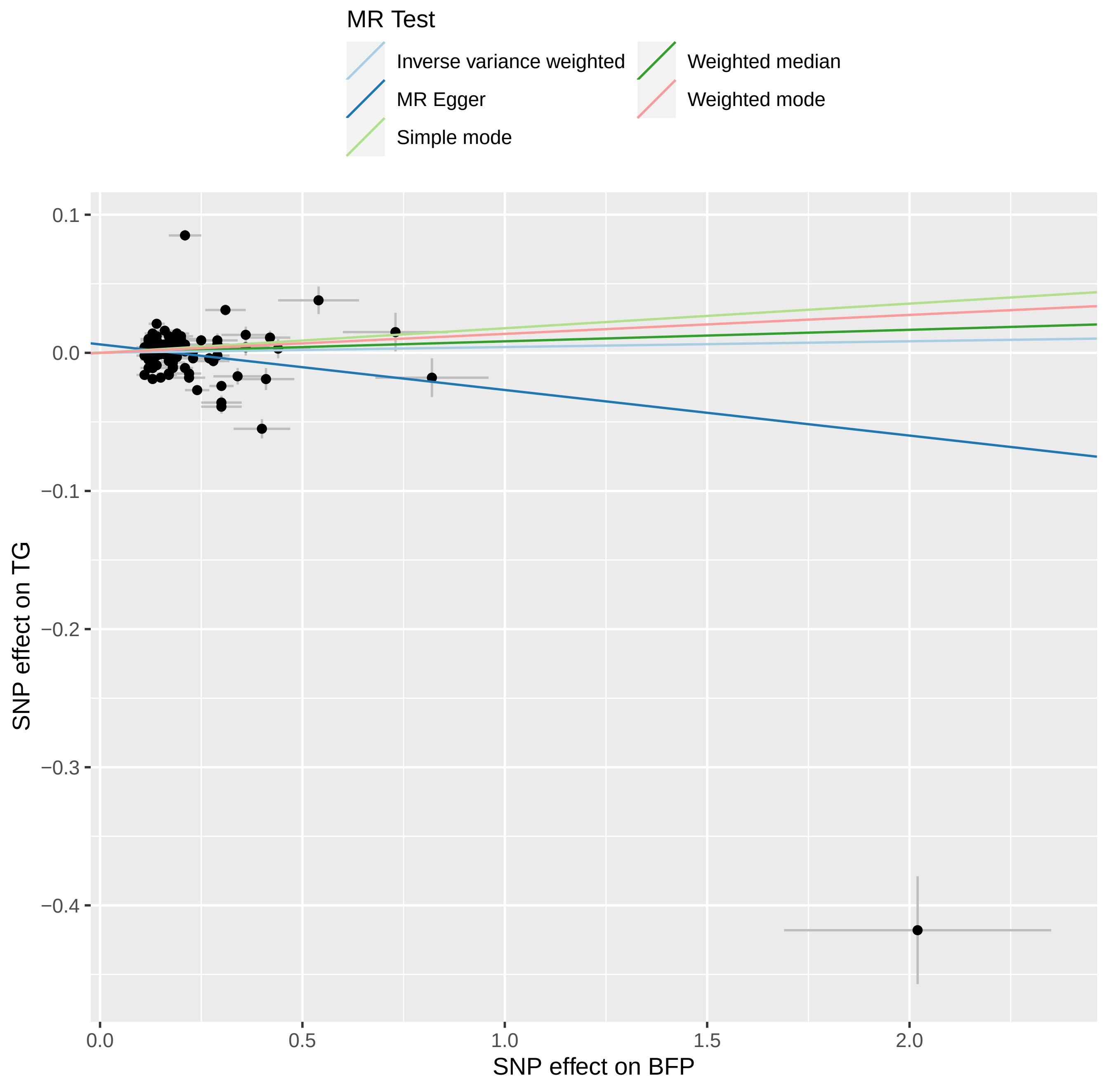
MR Test
Inverse variance weighted
MR Egger
Simple mode
Weighted median
Weighted mode
0.1
0.0
−0.1
−0.2
−0.3
−0.4
SNP effect on TG
0.0
0.5
1.0
1.5
2.0
SNP effect on BFP

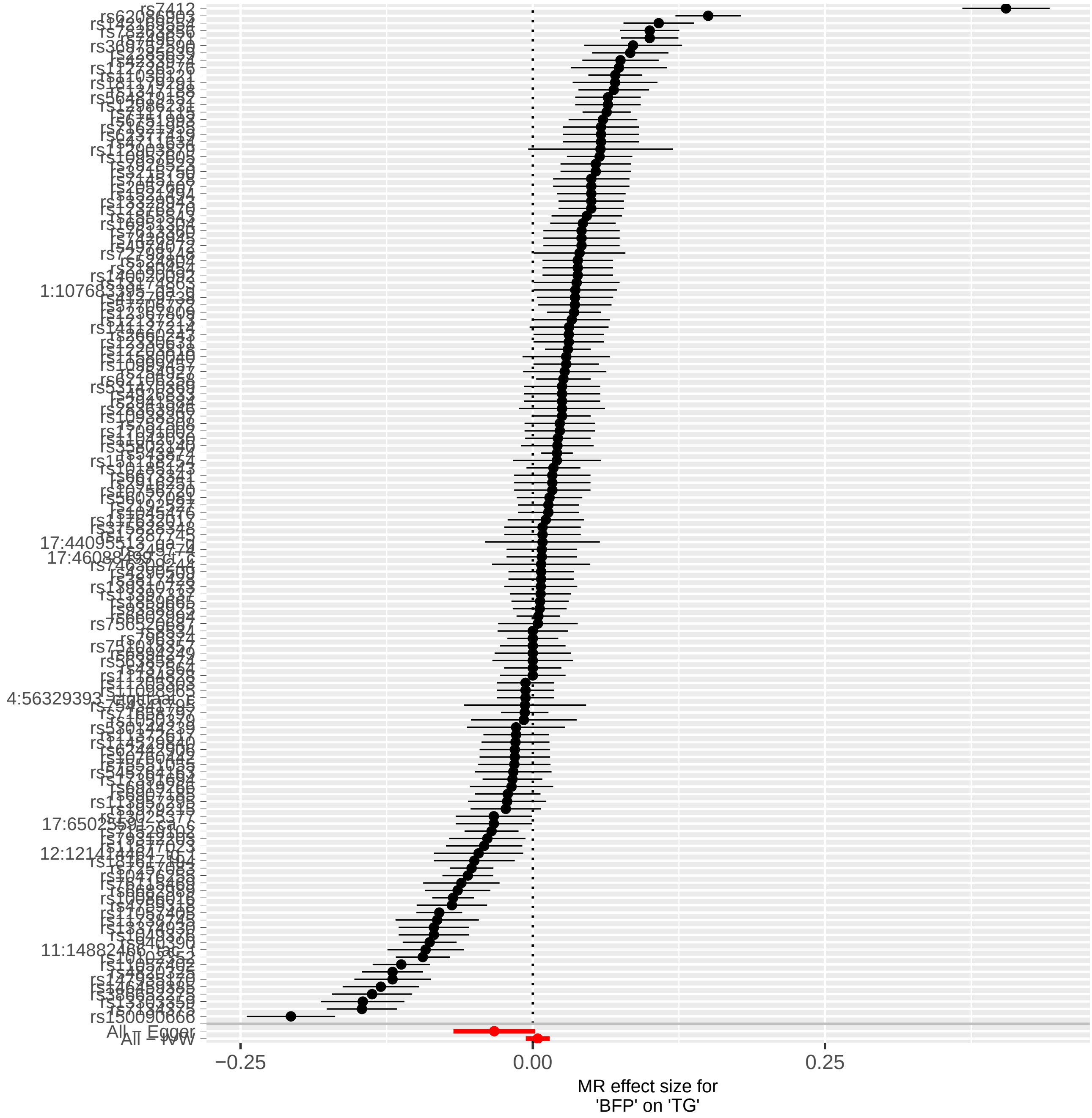
rs7412
rs150090666
All – Egger
All – IVW
−0.25
0.00
0.25
MR effect size for
'BFP' on 'TG'

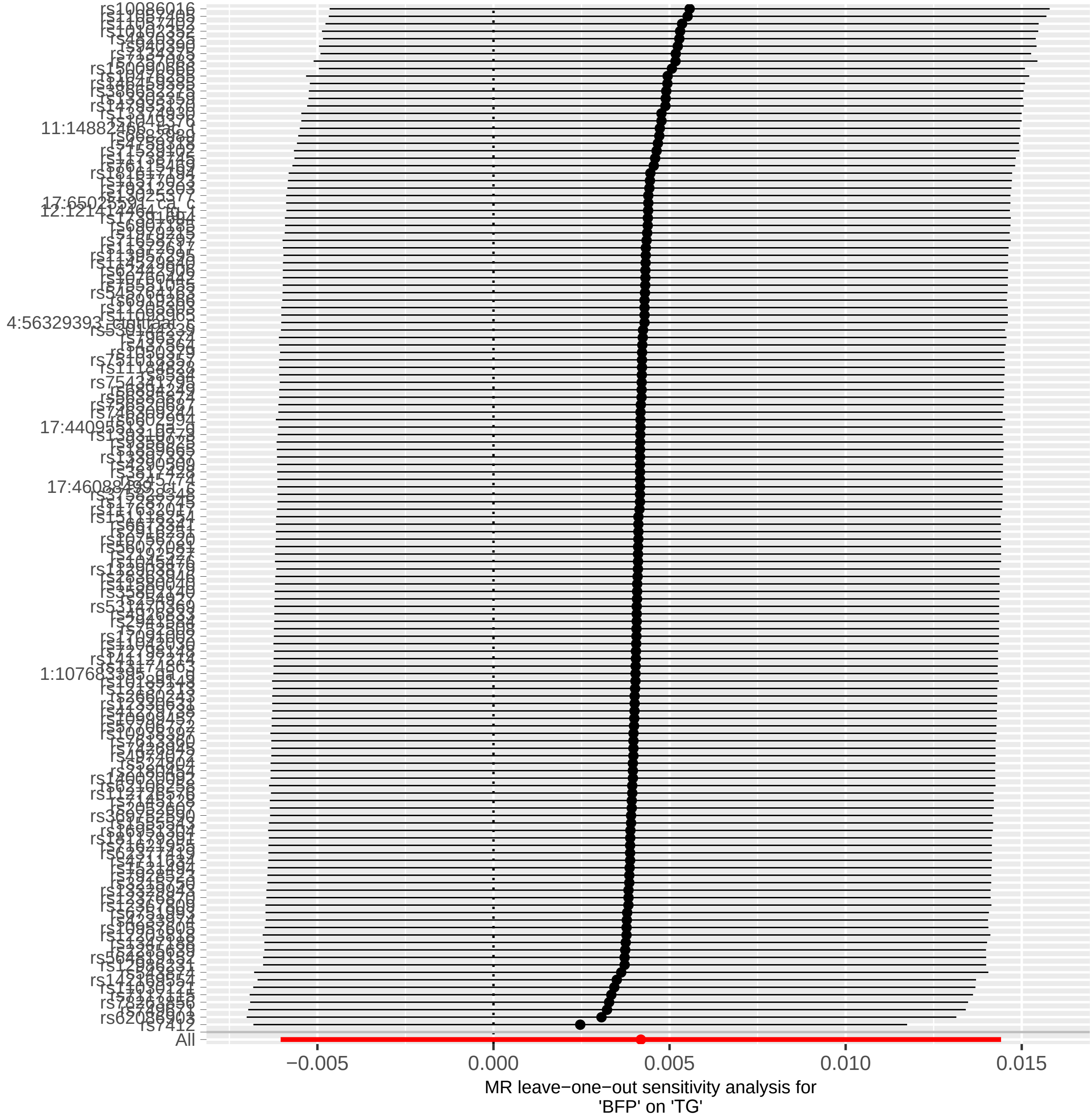
rs10086016
rs7412
All
−0.005
0.000
0.005
0.010
0.015
MR leave−one−out sensitivity analysis for
'BFP' on 'TG'

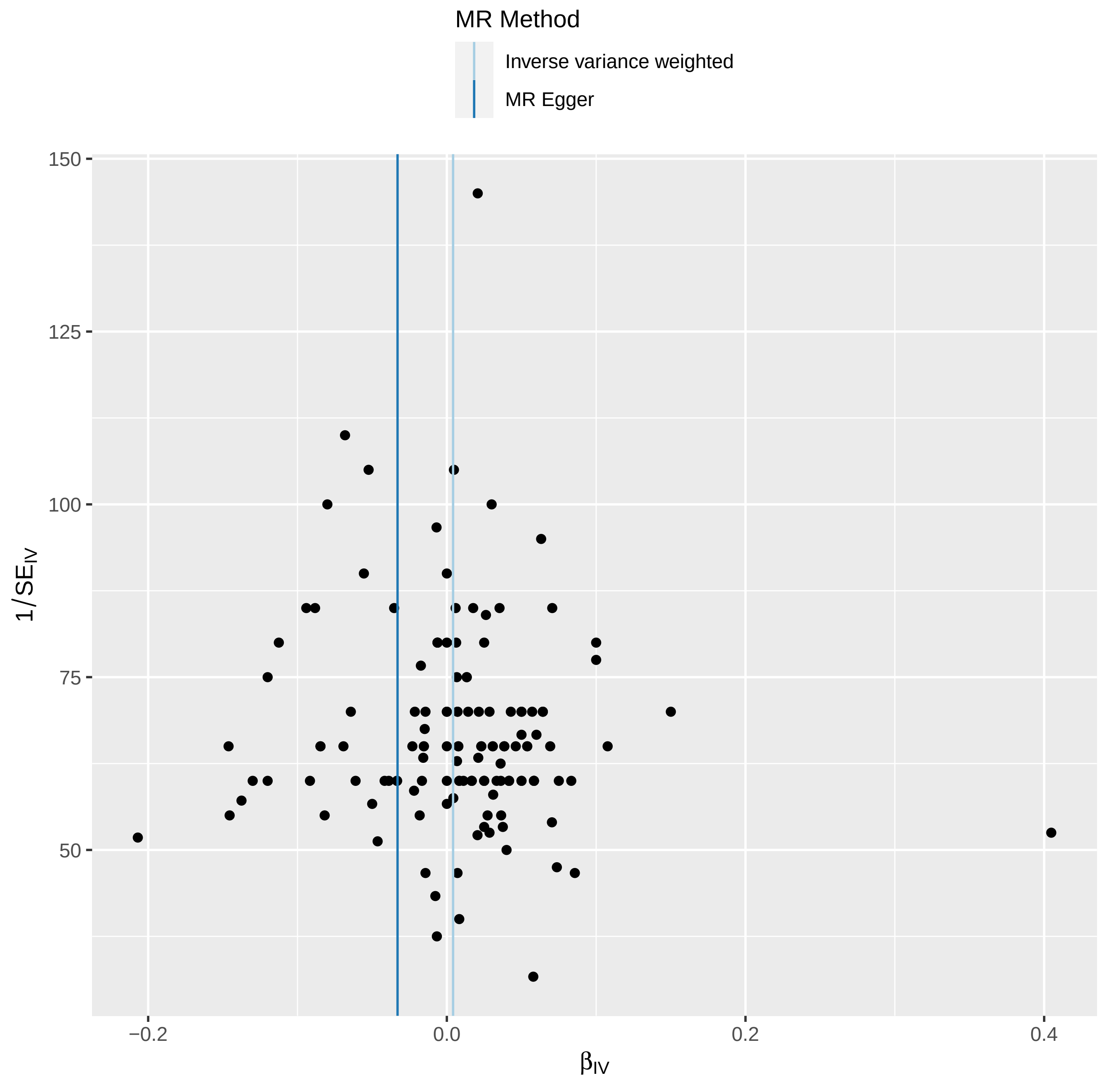

MR Method
Inverse variance weighted
MR Egger
150
125
100
75
50
1/SE$_{IV}$
−0.2
0.0
0.2
0.4
β$_{IV}$
