## Supplementary material for "Sex-stratified GWAS of Body Fat Percentage after Adjusting for Testosterone and SHBG in the UK Biobank": Plots regarding MR Analysis in Males

## 1- T2D

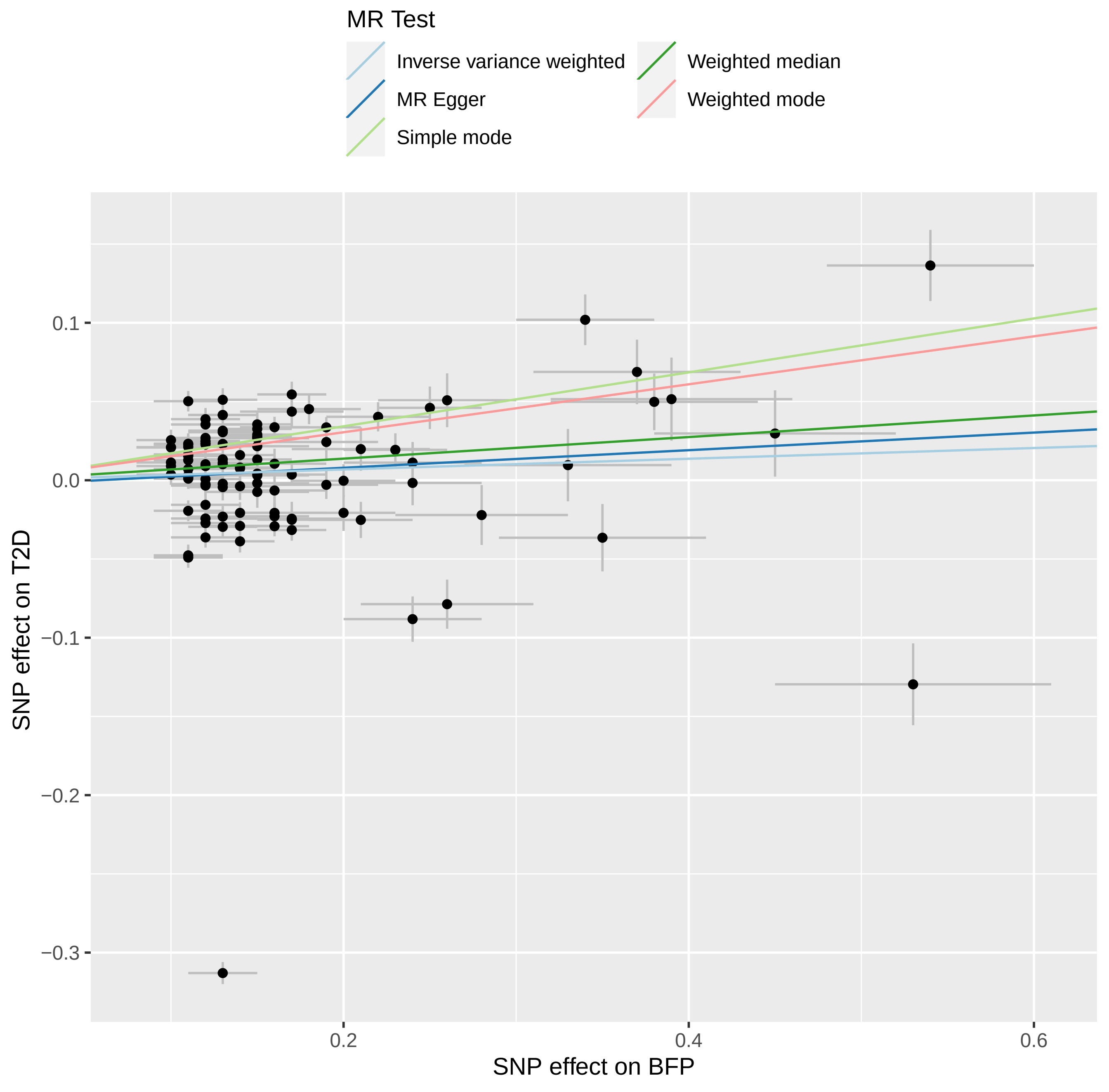
MR Test
Inverse variance weighted
MR Egger
Simple mode
Weighted median
Weighted mode
SNP effect on T2D
0.1
0.0
−0.1
−0.2
−0.3
0.2
0.4
0.6
SNP effect on BFP

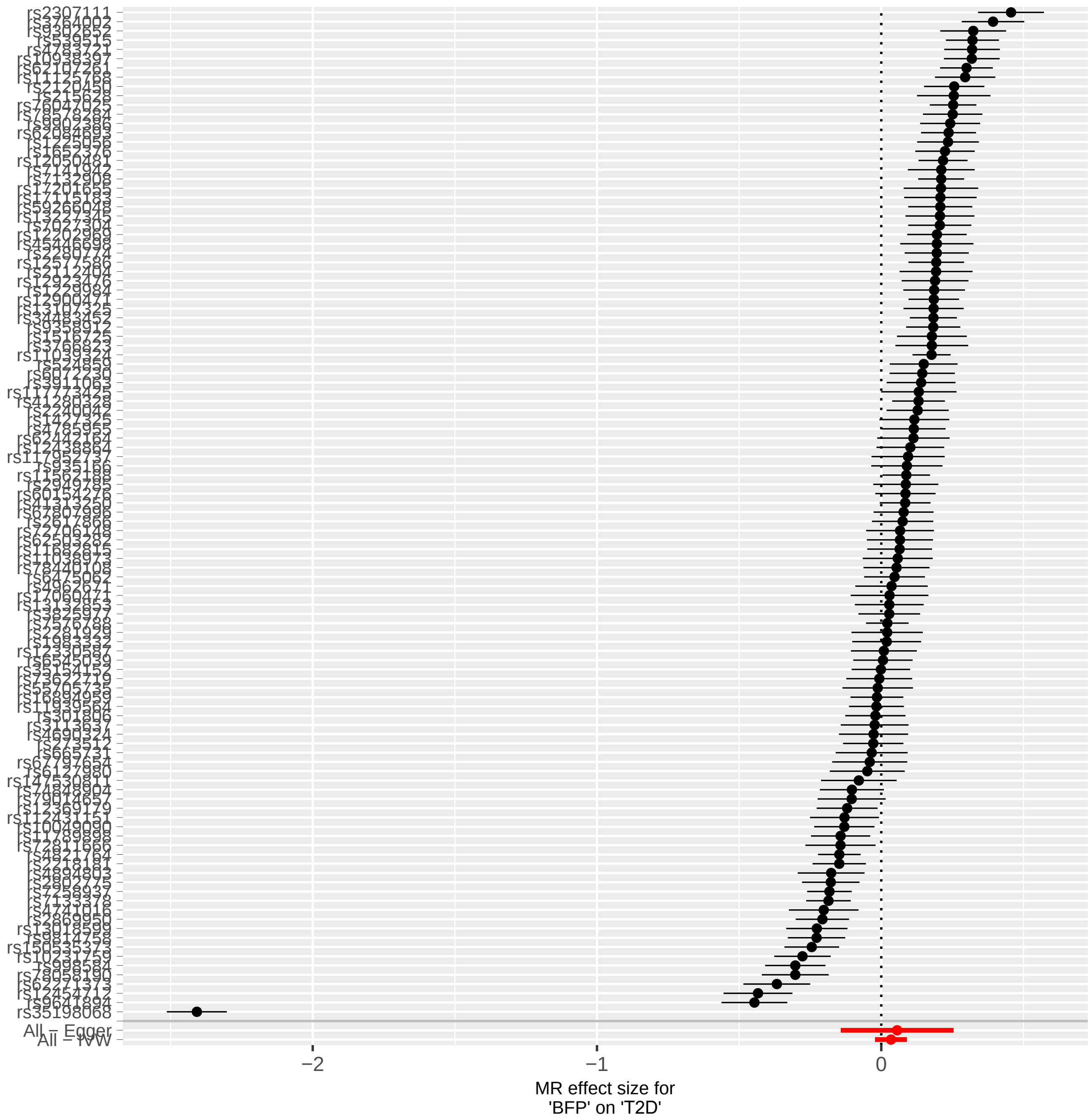
rs2307111
rs35198068
All – Egger
All – IVW
−2
−1
0
MR effect size for
'BFP' on 'T2D'

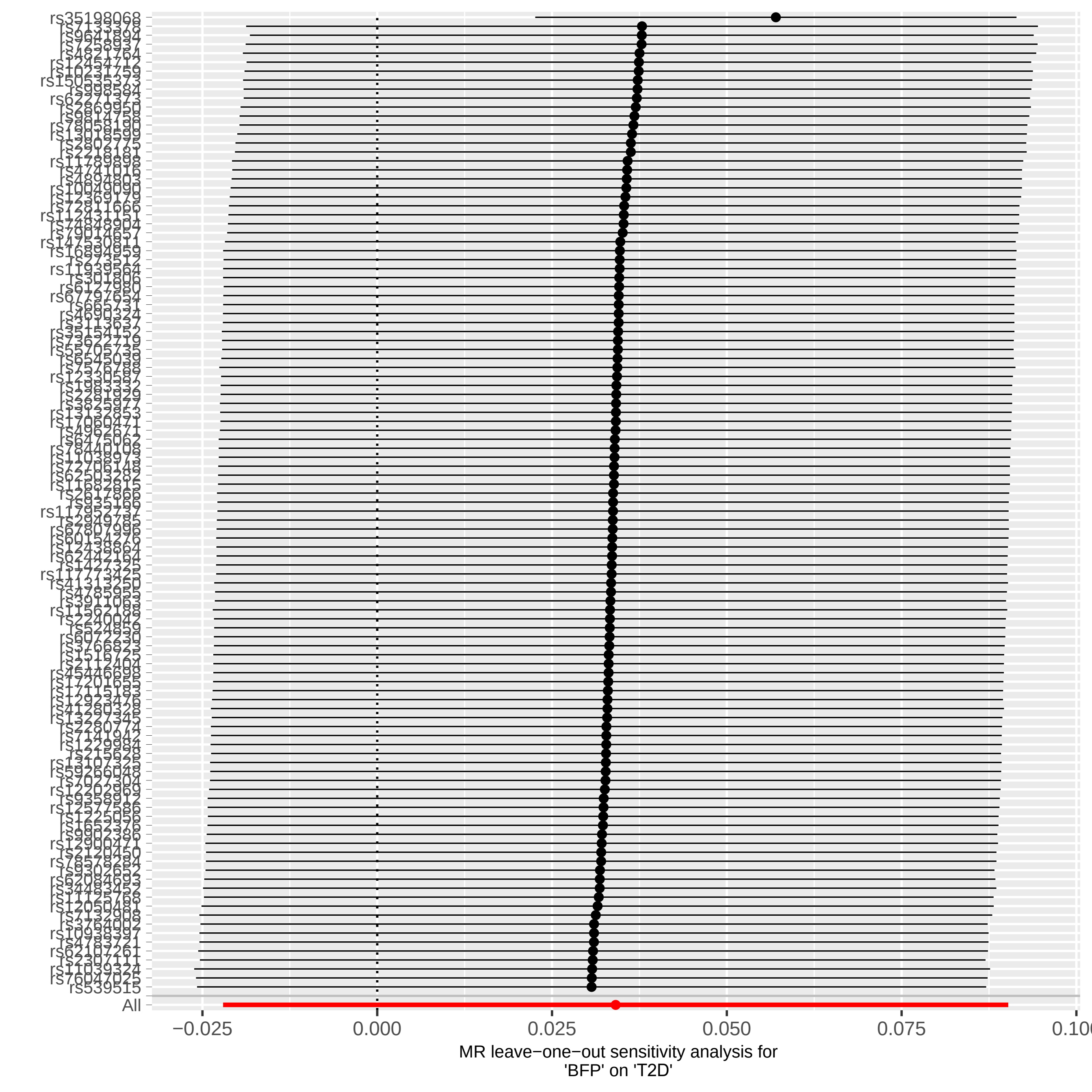
rs35198068
rs539515
All
−0.025
0.000
0.025
0.050
0.075
MR leave−one−out sensitivity analysis for
'BFP' on 'T2D'

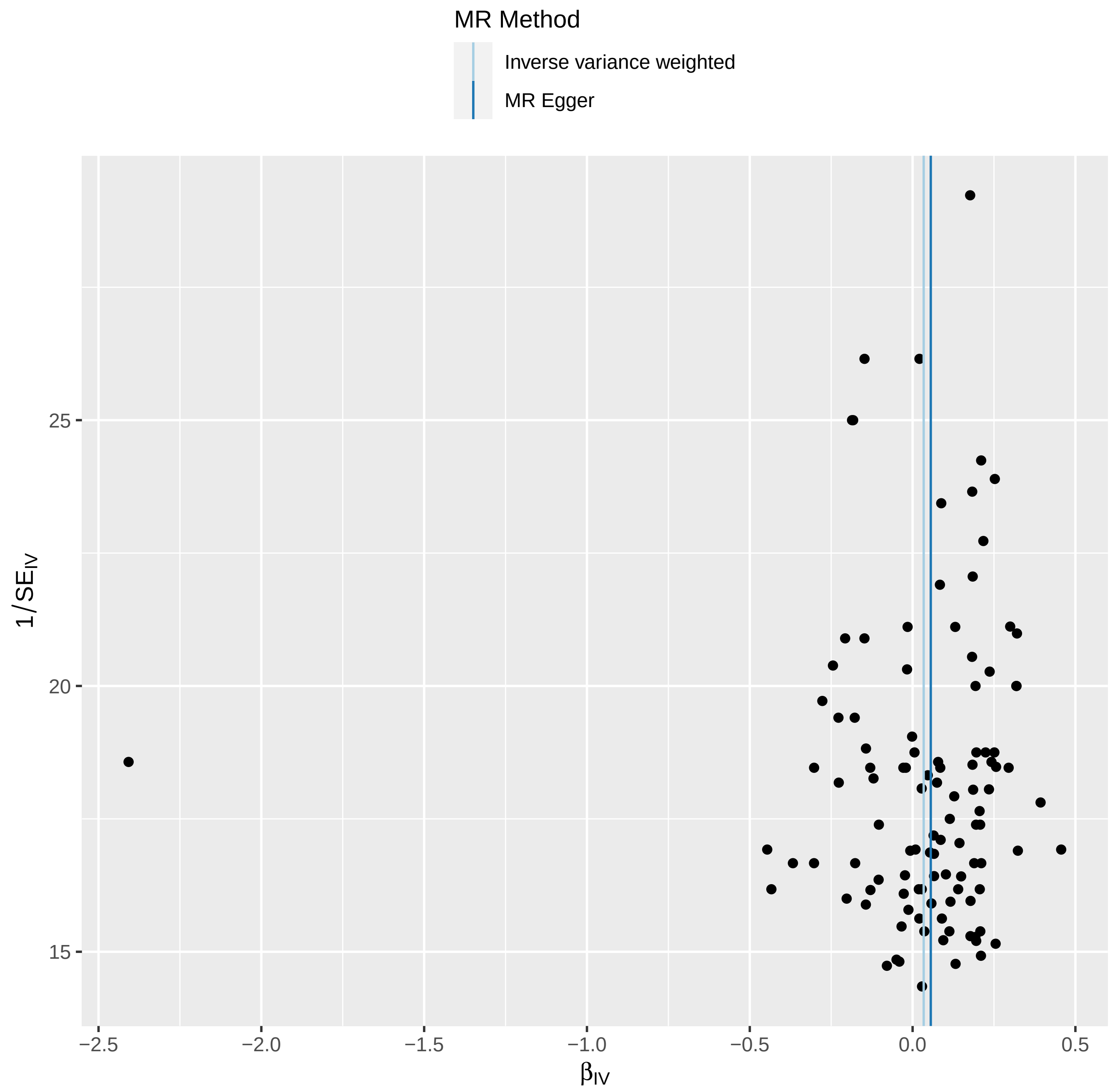
MR Method
Inverse variance weighted
MR Egger
1/SE_IV
25
20
15
−2.5
−2.0
−1.5
−1.0
−0.5
0.0
0.5
β_IV

### 2- CAD

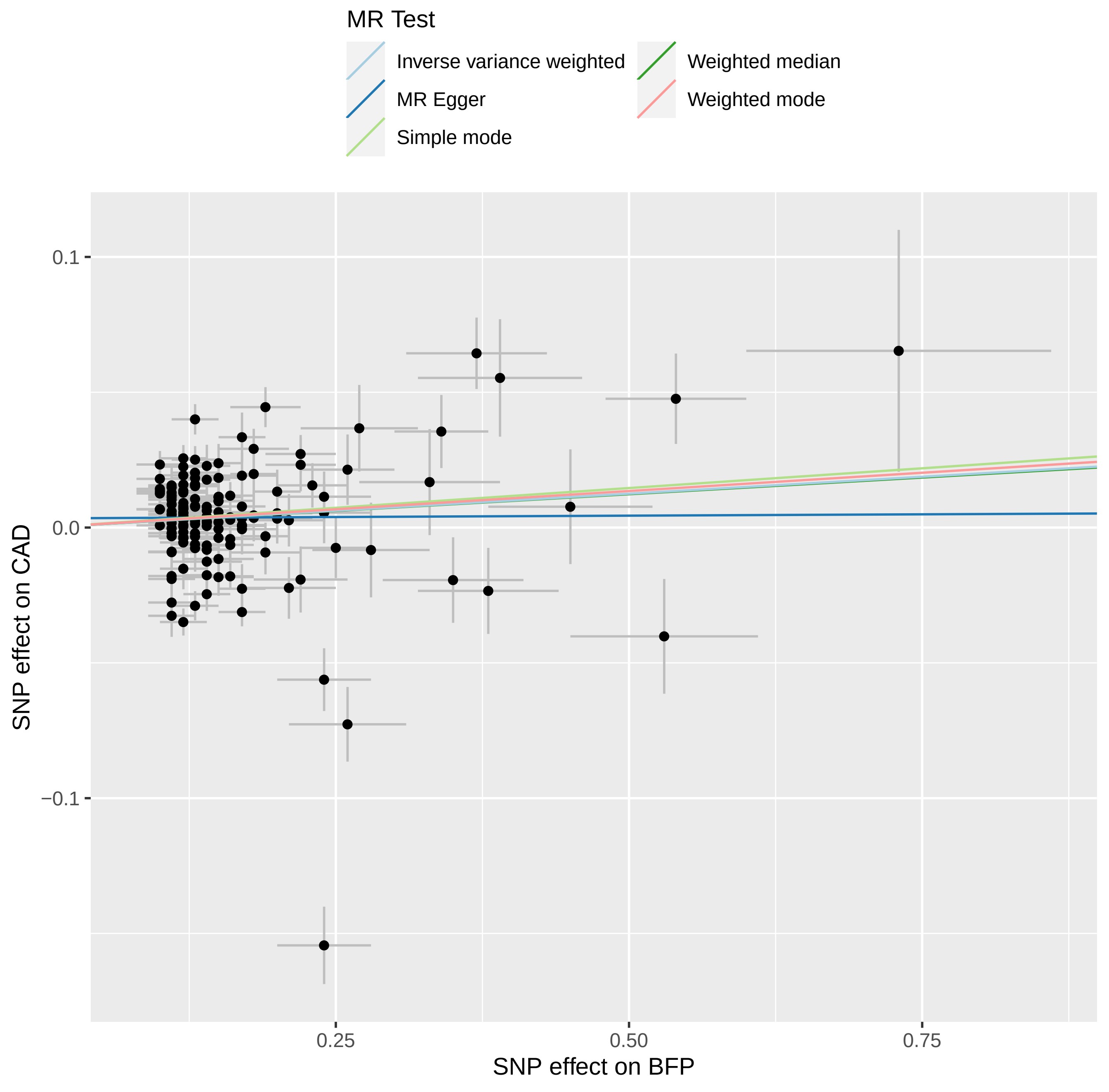

MR Test
Inverse variance weighted
MR Egger
Simple mode
Weighted median
Weighted mode
0.1
0.0
−0.1
0.25
0.50
0.75
SNP effect on BFP
SNP effect on CAD

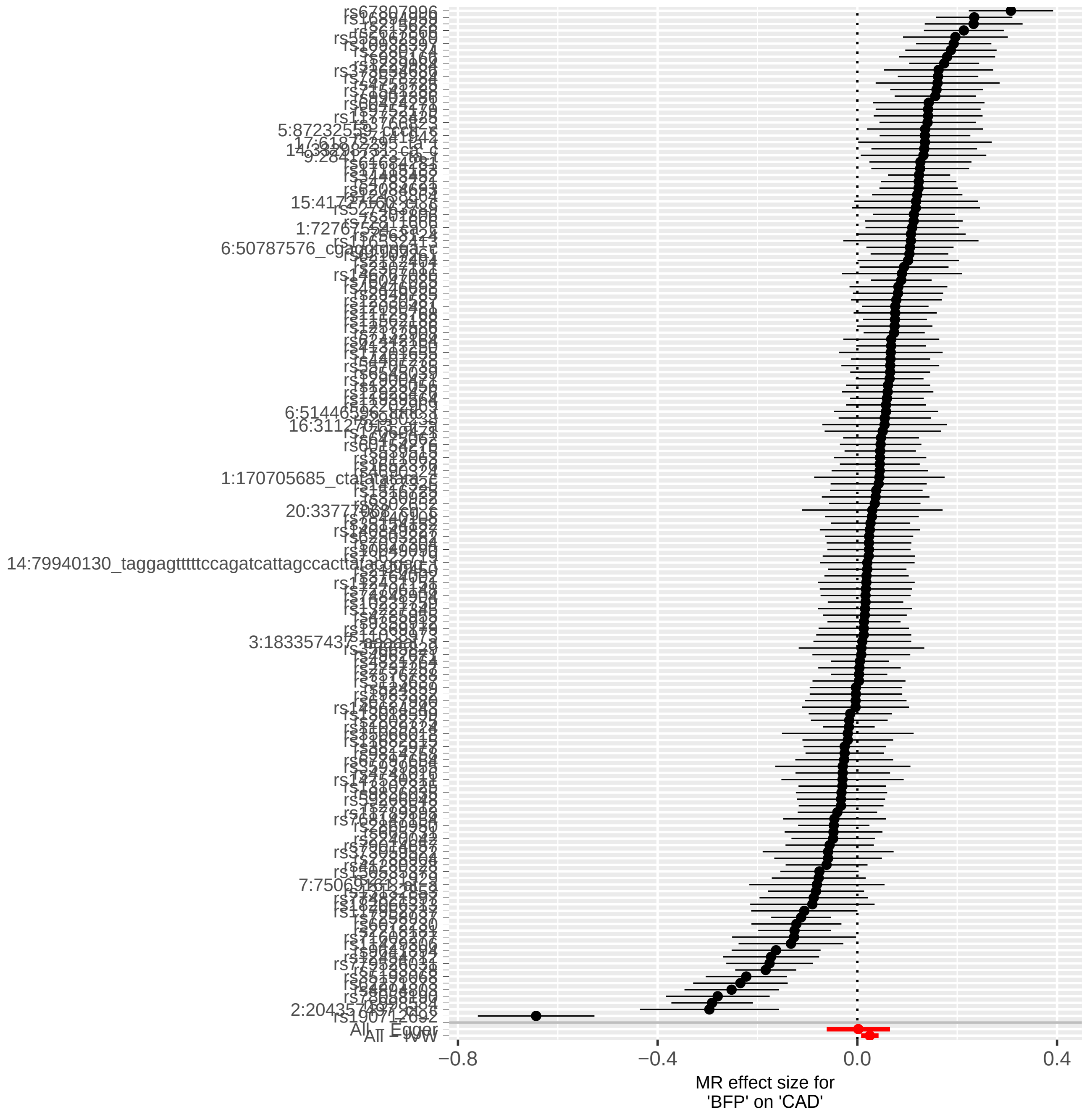
All − Egger
All − IVW
−0.8
−0.4
0.0
0.4
MR effect size for
'BFP' on 'CAD'

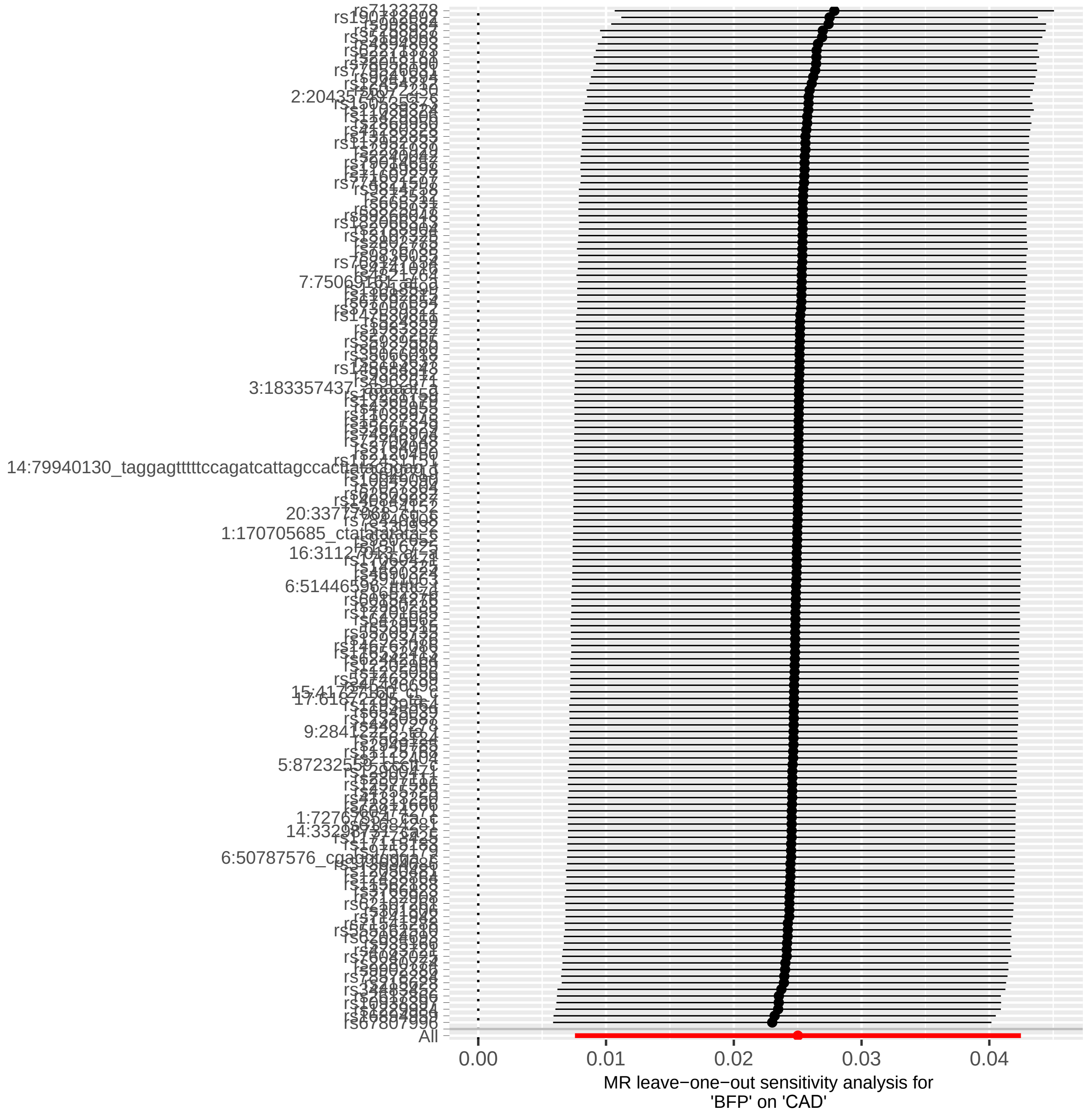
All
0.00
0.01
0.02
0.03
0.04
MR leave−one−out sensitivity analysis for
'BFP' on 'CAD'

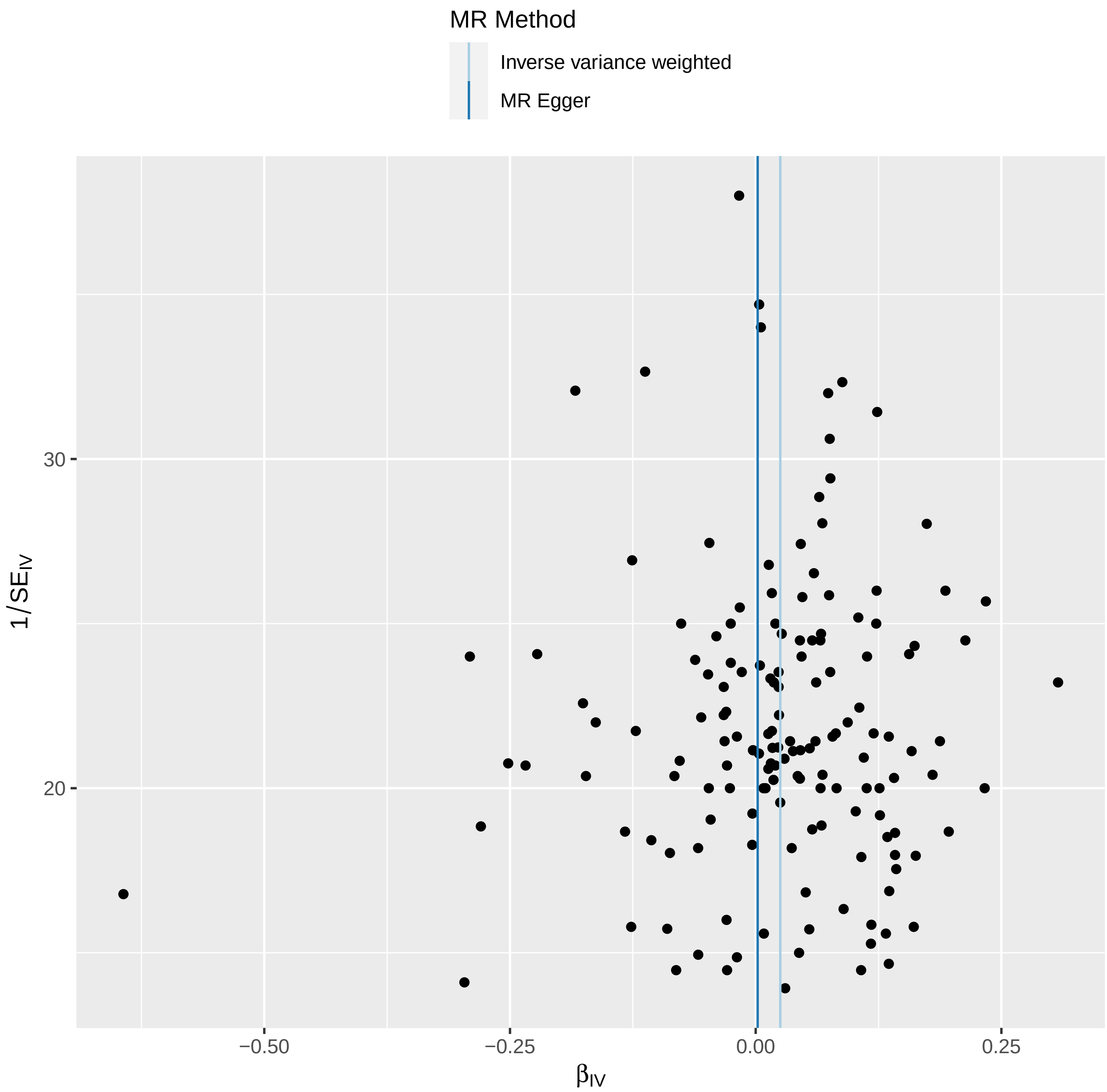

MR Method
Inverse variance weighted
MR Egger
30
20
1/SE_IV
−0.50
−0.25
0.00
0.25
β_IV

### 3- HDL

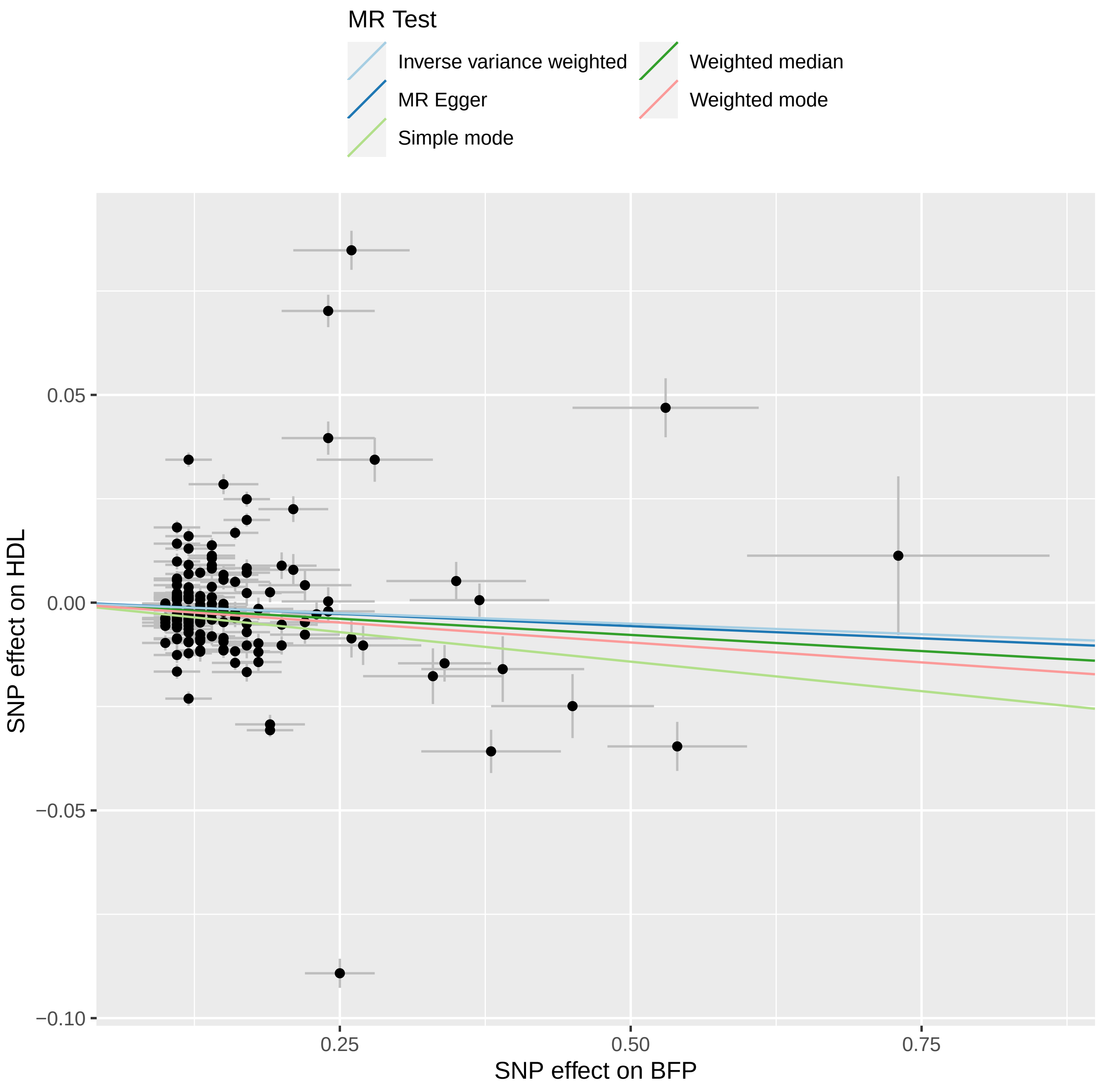
MR Test
Inverse variance weighted
MR Egger
Simple mode
Weighted median
Weighted mode
SNP effect on HDL
0.05
0.00
−0.05
−0.10
0.25
0.50
0.75
SNP effect on BFP

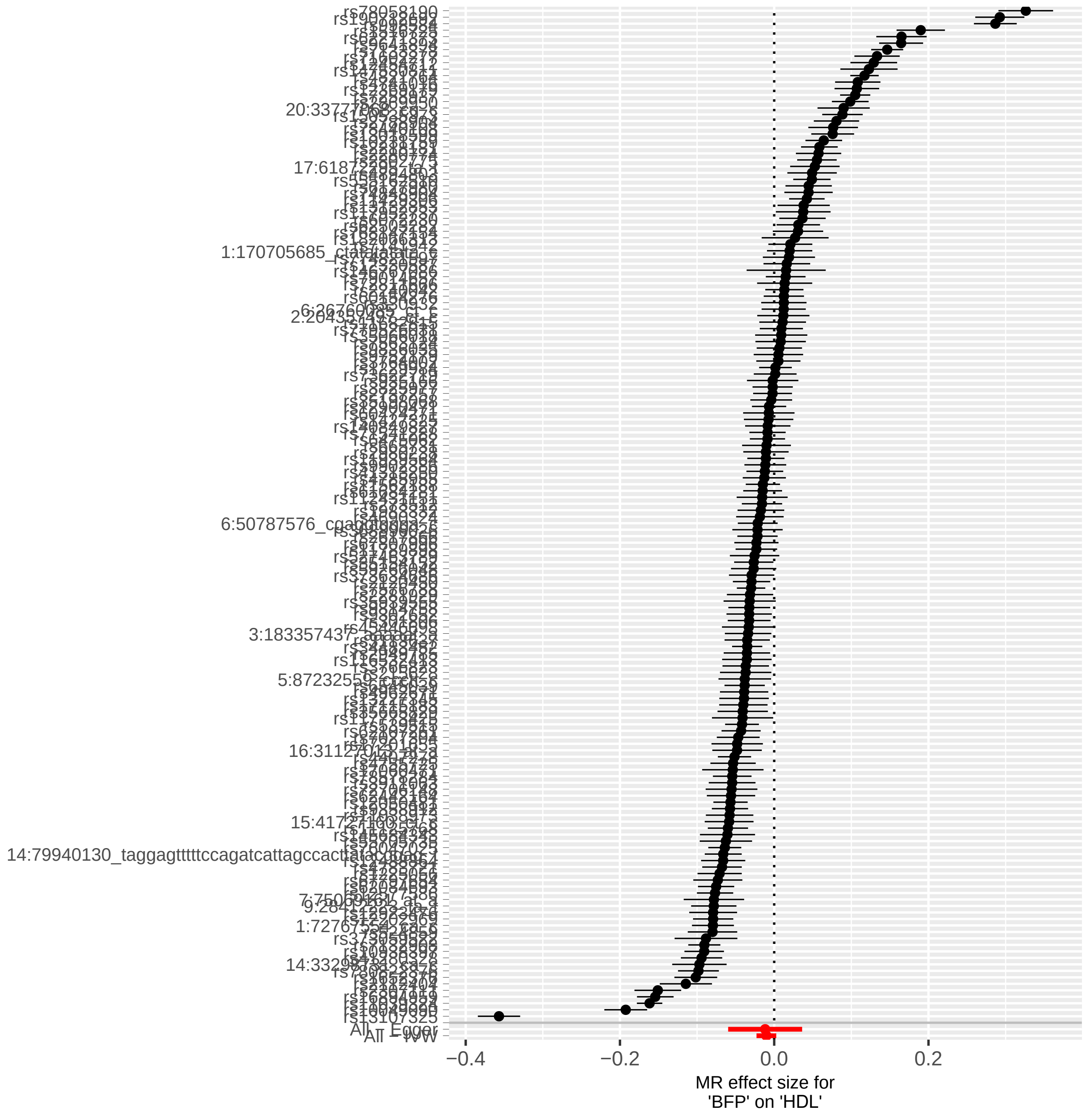

All – Egger
All – IVW
−0.4
−0.2
0.0
0.2
MR effect size for
'BFP' on 'HDL'

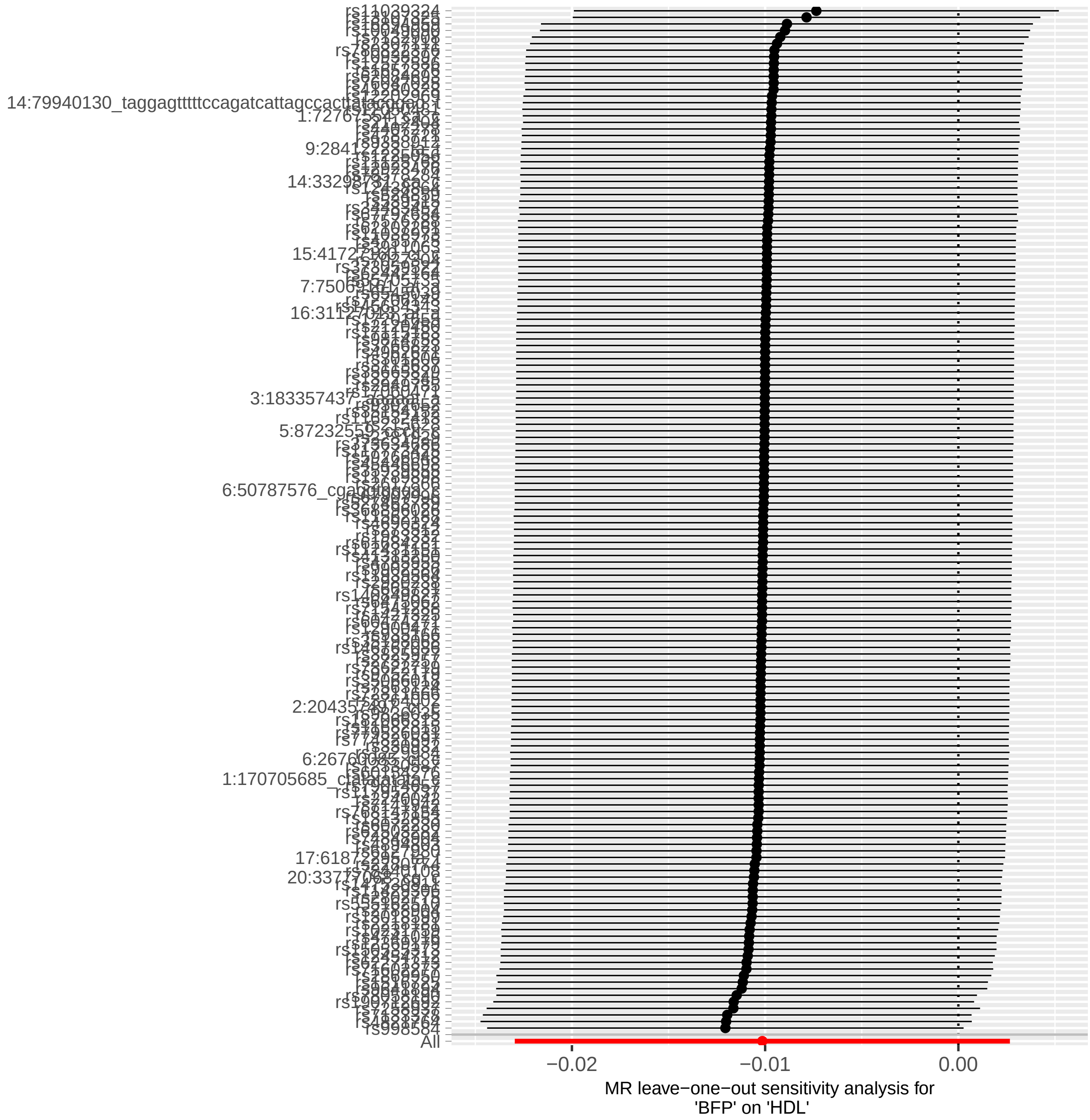

rs11039324
rs998584
All
−0.02
−0.01
0.00
MR leave−one−out sensitivity analysis for
'BFP' on 'HDL'

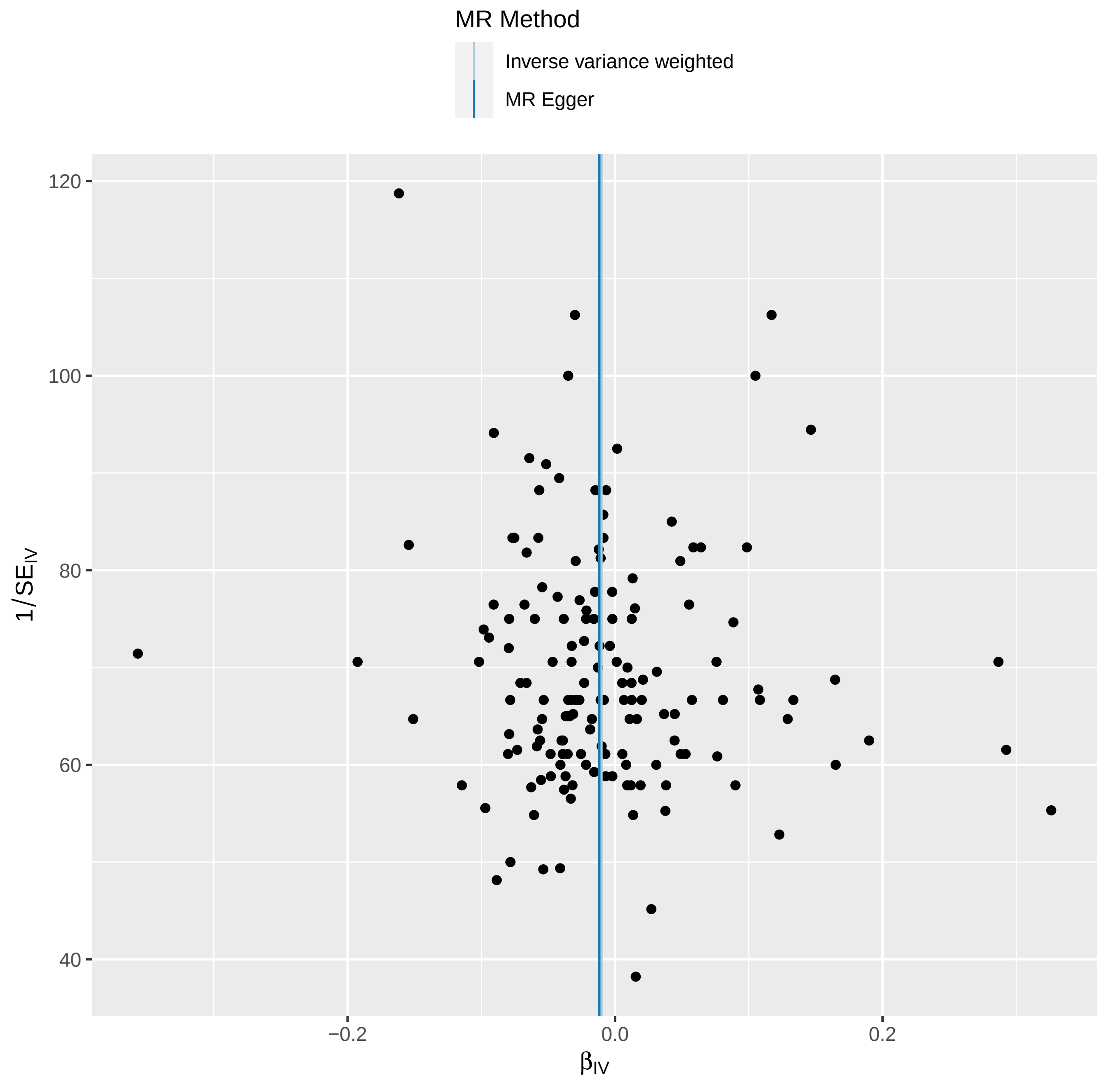

MR Method
Inverse variance weighted
MR Egger
120
100
80
60
40
1/SE$_{IV}$
−0.2
0.0
0.2
β$_{IV}$

# 4- TG

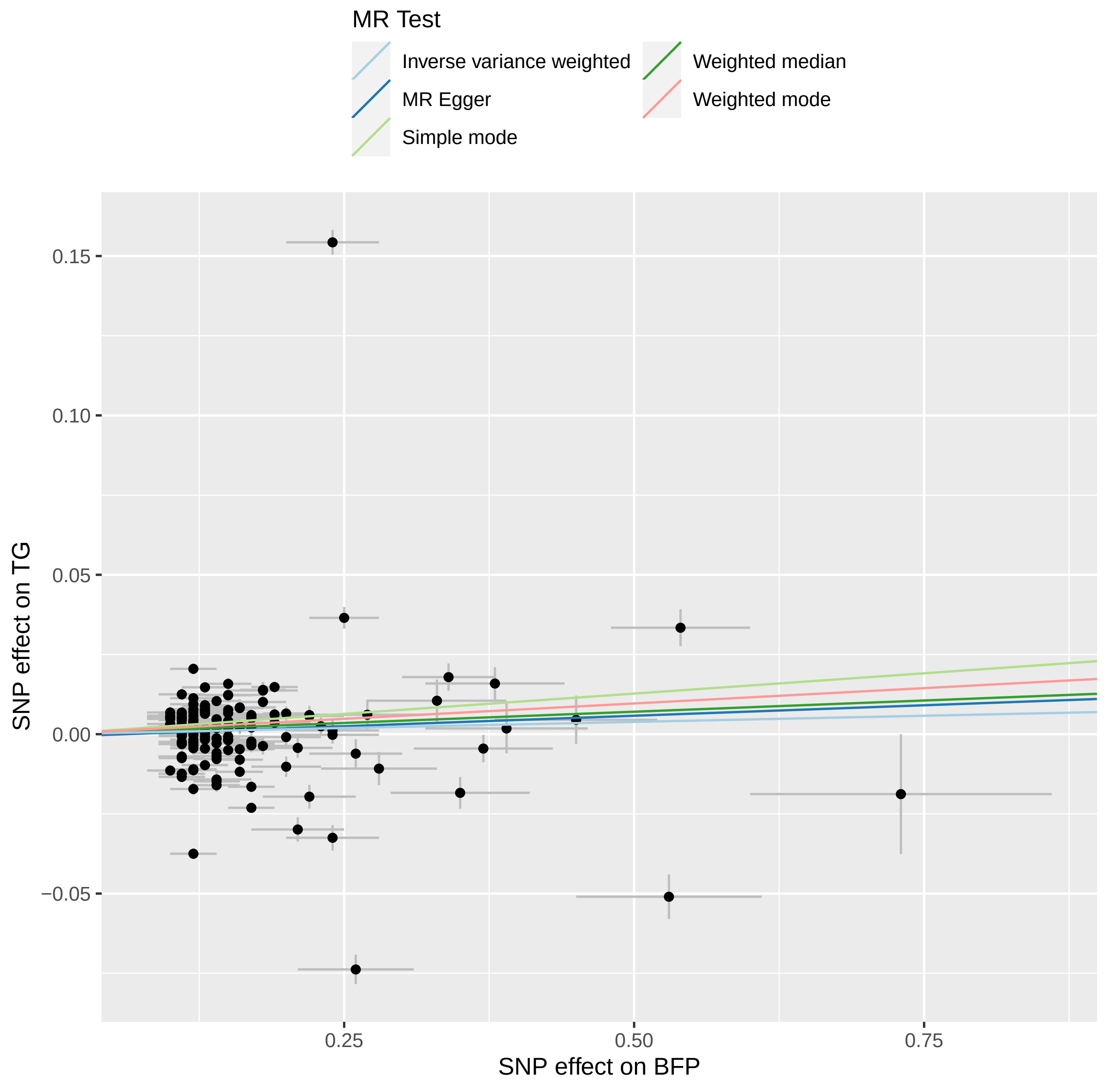
MR Test
Inverse variance weighted
MR Egger
Simple mode
Weighted median
Weighted mode
0.15
0.10
0.05
0.00
−0.05
SNP effect on TG
0.25
0.50
0.75
SNP effect on BFP

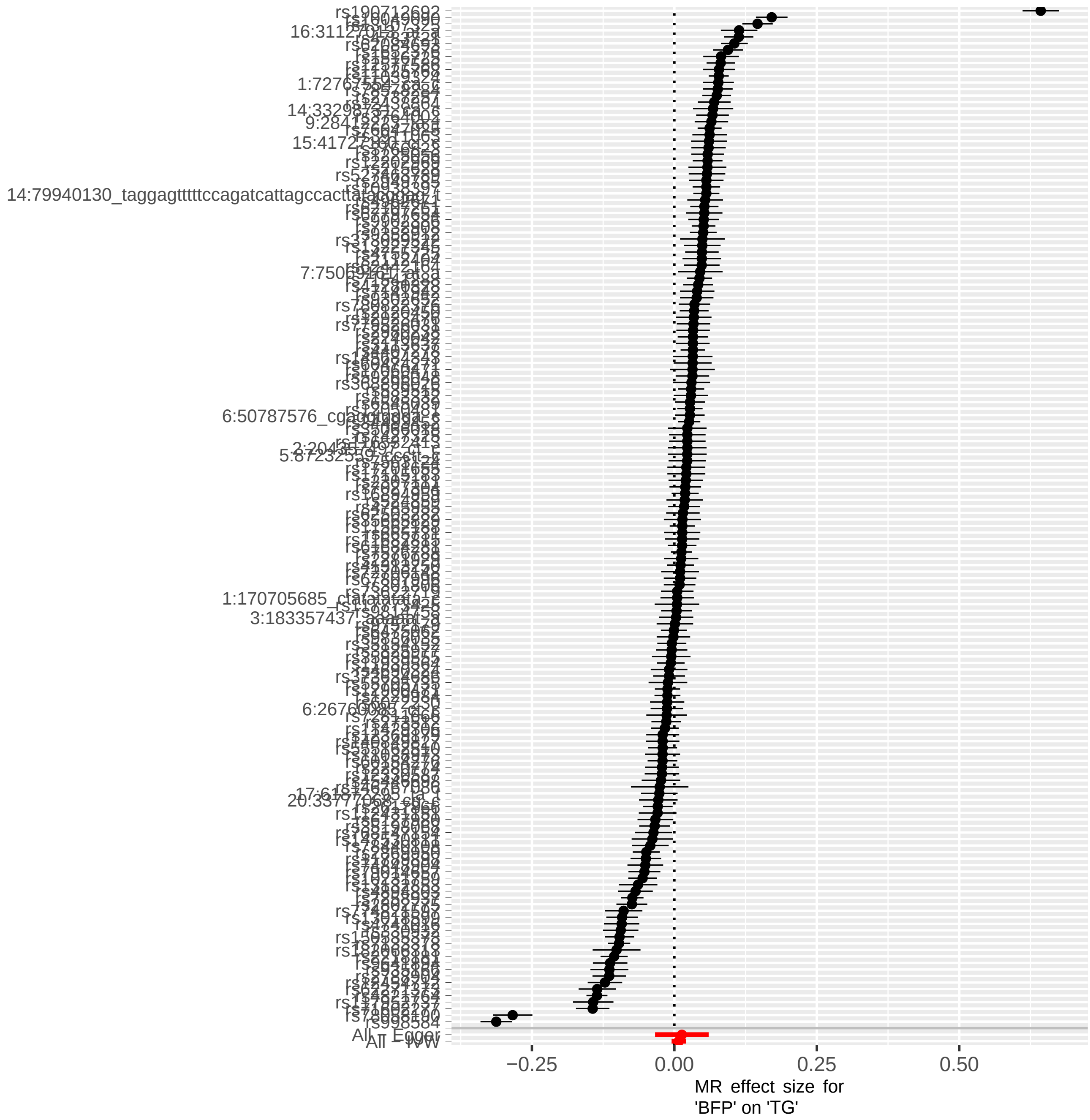
rs190712692
rs998584
All - Egger
All - IVW
−0.25
0.00
0.25
0.50
MR effect size for
'BFP' on 'TG'

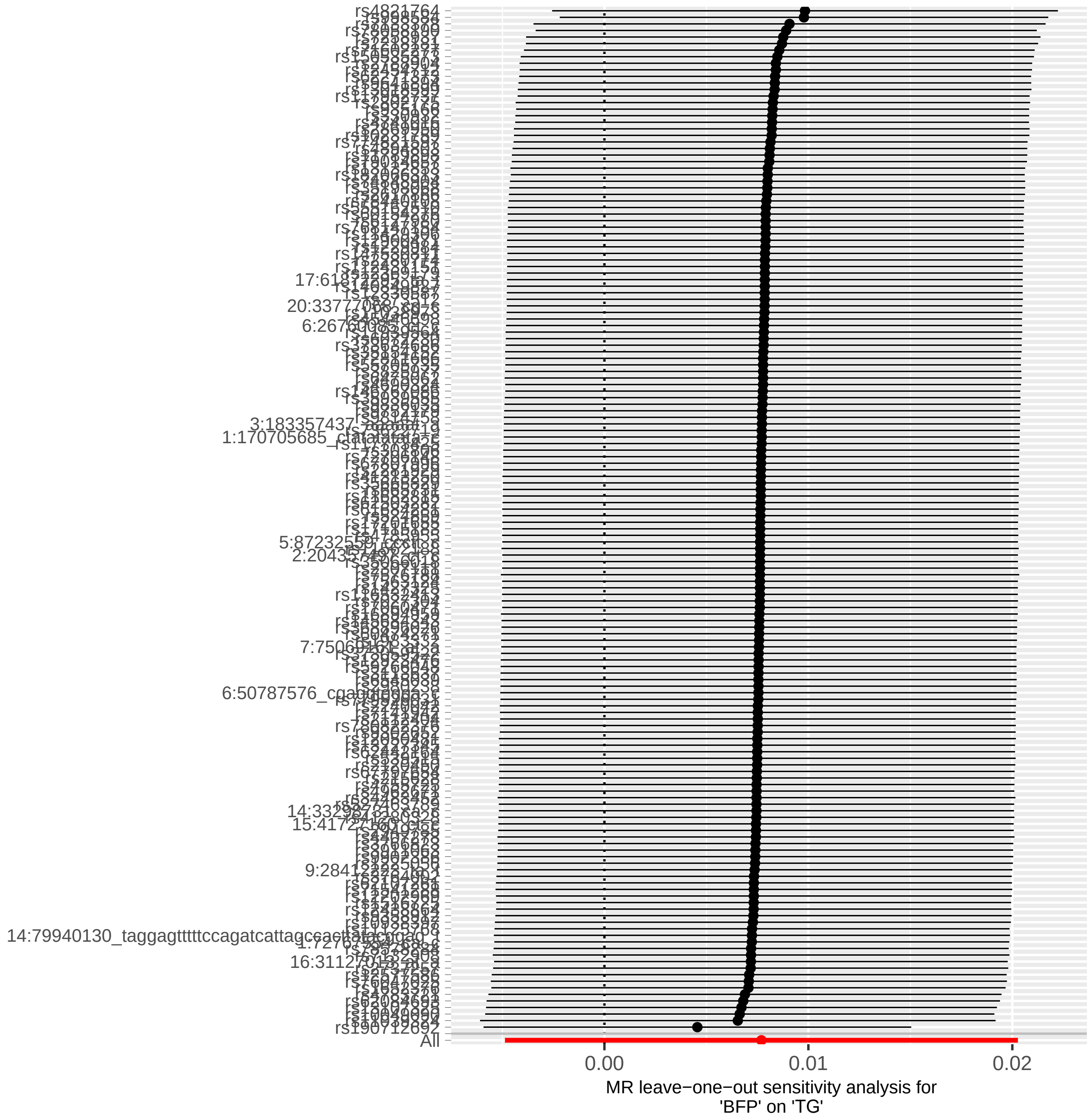
rs4821764
rs190712092
All
0.00
0.01
0.02
MR leave−one−out sensitivity analysis for
'BFP' on 'TG'

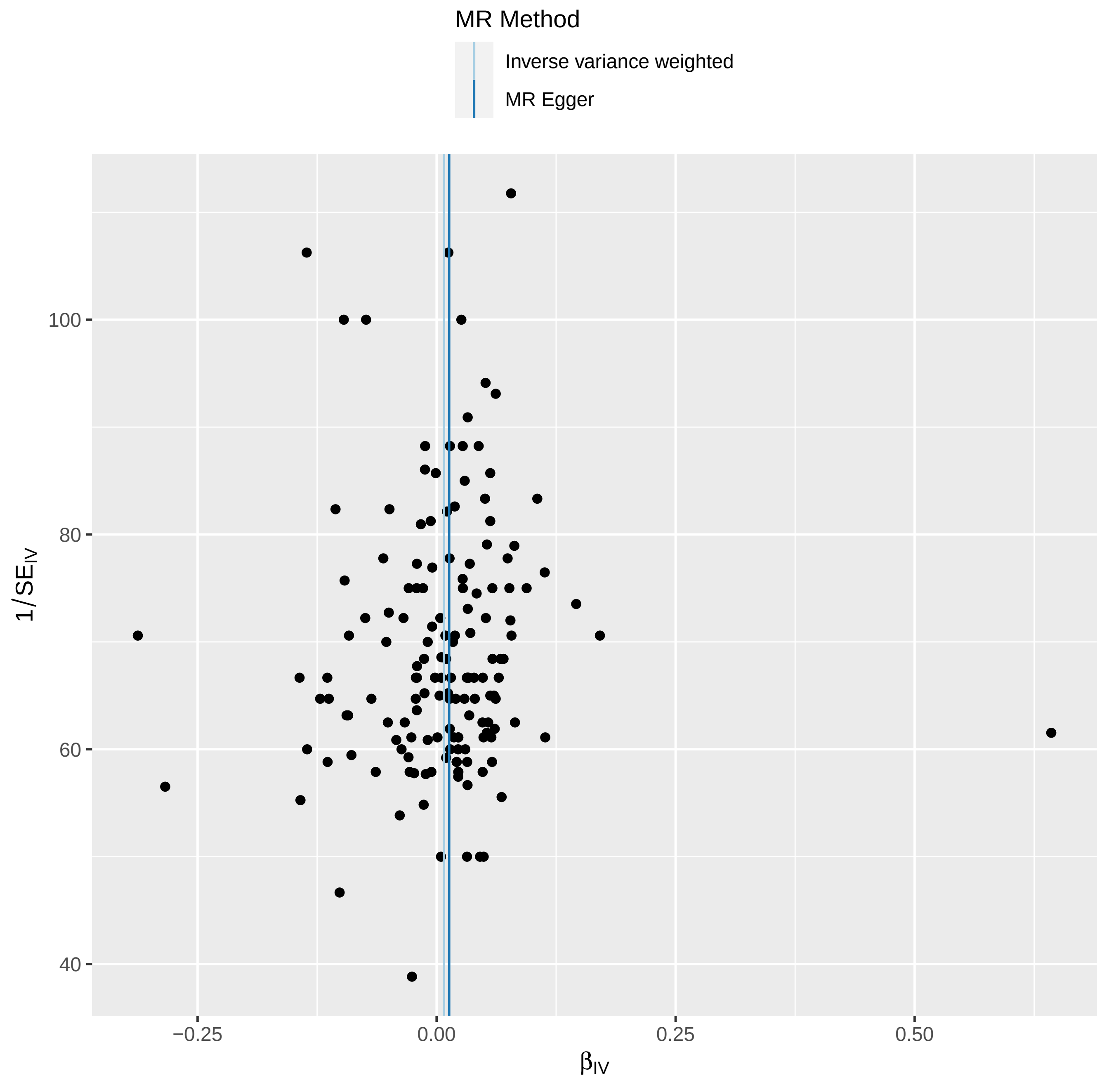

MR Method
Inverse variance weighted
MR Egger
100
80
60
40
1/SE$_{IV}$
−0.25
0.00
0.25
0.50
β$_{IV}$
