## Supplementary Tables and Figures for "Sex-stratified GWAS of Body Fat Percentage after Adjusting for Testosterone and SHBG in the UK Biobank"

### Supplemental Material

**Table S1:** The list of medical treatment (Unique Data Identifier (UDI) 20003-0.0) that subjects on them were excluded from the analysis

| Males | Females |
|---|---|
| 1. anastrozole<br>2. arimidex 1mg tablet<br>3. buserelin<br>4. buserelin product<br>5. clomid 50mg tablet<br>6. conjugated oestrogens 0.3mg / medroxyprogesterone 1.5mg tab<br>7. cyclogest 200mg suppository<br>8. cyprostat 50mg tablet<br>9. cyproterone<br>10. cyproterone acetate+ethinyloestradiol<br>11. de-capeptyl sr 4.2mg injection (pdr for recon)+diluent<br>12. deltahydrocortisone<br>13. deltastab 1mg tablet<br>14. dexamethasone<br>15. drogenil<br>16. dydrogesterone<br>17. ethinylnortestosterone<br>18. ethinyloestradiol product<br>19. ethinyloestradiol+norethisterone acetate 20mcg/1mg tablet<br>20. ethinyloestradiol+norgestimate 35mcg/250mcg tablet<br>21. ethynodiol diacetate<br>22. flutamide<br>23. gonadorelin<br>24. goserelin<br>25. goserelin product<br>26. hydrocortisone<br>27. hydrocortisone product<br>28. hydrocortistab 20mg tablet<br>29. hydrocortone 10mg tablet<br>30. letrozole<br>31. leuprorelin<br>32. levonorgestrel<br>33. levonorgestrel product<br>34. loestrin<br>35. medrone 2mg tablet<br>36. medroxyprogesterone<br>37. mestranol+norethisterone 50micrograms/1mg tablet<br>38. methylprednisolone<br>39. methyltestosterone product<br>40. micronor<br>41. microval tablet<br>42. nafarelin | 1. methyltestosterone product<br>2. testotop tts 15mg transdermal patch<br>3. yohimbine/pemoline/methyltestosterone<br>4. testosterone product<br>5. primoteston depot 250mg/1ml oily injection<br>6. testoderm 6mg/24hours transdermal patch<br>7. testogel 50mg gel 5g sachet<br>8. cyprostat 50mg tablet<br>9. flutamide<br>10. drogenil<br>11. anastrozole<br>12. arimidex 1mg tablet<br>13. letrozole<br>14. cyproterone<br>15. cyproterone acetate+ethinyloestradiol<br>16. cyproterone acetate+ethinylestradiol<br>17. dydrogesterone<br>18. proscar 5mg tablet<br>19. clomid 50mg tablet<br>20. medrone 2mg tablet<br>21. prednisone<br>22. prednisolone<br>23. prednesol 5mg tablet<br>24. hydrocortistab 20mg tablet<br>25. hydrocortone 10mg tablet<br>26. methylprednisolone<br>27. prednisolone product<br>28. hydrocortisone<br>29. deltastab 1mg tablet<br>30. precortisyl 1mg tablet<br>31. deltahydrocortisone<br>32. hydrocortisone product<br>33. dexamethasone |

| |
|---|
| 43. norethisterone<br>44. norethisterone product<br>45. norgeston tablet<br>46. noriday<br>47. norinyl-1 tablet<br>48. noristerat 200mg/1ml oily injection<br>49. ortho-novin 1/50 tablet<br>50. precortisyl 1mg tablet<br>51. prednesol 5mg tablet<br>52. prednisolone<br>53. prednisolone product<br>54. prednisone<br>55. primoteston depot 250mg/1ml oily injection<br>56. progesterone product<br>57. proscar 5mg tablet<br>58. prostap sr 3.75mg injection (pdr for recon)+diluent+kit<br>59. suprefact 100micrograms nasal spray<br>60. testoderm 6mg/24hours transdermal patch<br>61. testogel 50mg gel 5g sachet<br>62. testosterone product<br>63. testotop tts 15mg transdermal patch<br>64. triptorelin<br>65. yohimbine/pemoline/methyltestosterone |

**Table S2:** The GWAS used to extract association of SNPs with SHBG, testosterone, TG, HDL, T2D, CAD, VAT, ASAT, and GFAT

| **Trait** | **First Author** | **Ethnicity** | **Includes UKBB** | **Sex Stratified** | **Subjects** | **Link to Download** |
|---|---|---|---|---|---|---|
| SHBG | Ruth et al [1] | European | Yes | Yes | 180,726 Males<br>189,473 Females | Males: https://www.ebi.ac.uk/gwas/studies/GCST90012109<br>Females: https://www.ebi.ac.uk/gwas/studies/GCST90012107 |
| Testosterone | Ruth et al [1] | European | Yes | Yes | 194,453 Males<br>230,454 Females | Males: https://www.ebi.ac.uk/gwas/studies/GCST90012113<br>Females: https://www.ebi.ac.uk/gwas/studies/GCST90012112 |
| HDL & TG | Graham et al [2] | European 80%<br>East Asian 9%<br>African 6%<br>Hispanic 3%<br>South Asian 2% | Yes | Yes | 1,654,960<br>51% Females | http://csg.sph.umich.edu/willer/public/glgc-lipids2021/results/sex_specific_summary_stats/ |
| T2D | Mahajan et al [3] | European | Yes | No | 74,124 Cases<br>824,006 Controls | http://diagram-consortium.org/downloads.html<br>Mahajan.NatGen2022.DIAMANTE-EUR.sumstat.zip |
| CAD | Aragam et al [4] | European | Yes | No | 181,522 Cases<br>984,168 Controls | http://ftp.ebi.ac.uk/pub/databases/gwas/summary_statistics/GCST90132001-GCST90133000/GCST90132314/ |
| GFAT, VAT & ASAT | Agrawal et al [5] | European 87% | Yes | Yes | 38,965<br>51% Females | https://cvd.hugeamp.org/downloads.html#summary |
| WHR adjusted for BMI | Pulit et al [6] | European | Yes | Yes | 315,284 Males<br>379,501 Females | https://zenodo.org/record/1251813#.Y-u7InbMKUl |

**Table S3:** Subjects characteristics

| | Males (N = 157,937) | | Females (N = 154,337) | |
|---|---|---|---|---|
| | **Mean** | **SD** | **Mean** | **SD** |
| **Age (years)** | 57.1 | 8.1 | 56.3 | 8.0 |
| **Albumin (g/L)** | 45.5 | 2.6 | 45.0 | 2.6 |
| **SHBG (nmol/L)** | 39.9 | 16.7 | 61.8 | 30.1 |
| **Testosterone (nmol/L)** | 12.0 | 3.7 | 1.1 | 0.6 |
| **Body fat percentage (%)** | 25.3 | 5.8 | 36.7 | 6.9 |

**Table S4:** Association of covariates with body fat percentage in males and females

| | **Males** | | | | **Females** | | | |
|---|---|---|---|---|---|---|---|---|
| | **Beta** | **SE** | **P** | **$R^2$** | **Beta** | **SE** | **P** | **$R^2$** |
| **Age** | 0.1559 | 0.0020 | <2E-16 | 0.034 | 0.1148 | 0.0002 | <2E-16 | 0.029 |
| **Centered Age$^2$** | -0.0010 | 0.0002 | 1.04E-5 | | -0.0022 | 0.0003 | <2E-16 | |
| **Albumin** | -0.0562 | 0.0054 | <2E-16 | 0.004 | -0.4242 | 0.0060 | <2E-16 | 0.026 |
| **Centered Albumin$^2$** | -0.0012 | 0.0012 | 0.33 | | 0.0135 | 0.0015 | <2E-16 | |
| **SHBG** | -0.0768 | 0.0013 | <2E-16 | 0.049 | -0.1142 | 0.0006 | <2E-16 | 0.183 |
| **Centered SHBG$^2$** | 0.0005 | 0.00002 | <2E-16 | | 0.0007 | 0.00001 | <2E-16 | |
| **Testosterone** | -0.2755 | 0.005 | <2E-16 | 0.084 | 1.3180 | 0.0328 | <2E-16 | 0.006 |
| **Centered Testosterone$^2$** | 0.0121 | 0.0006 | <2E-16 | | -0.2535 | 0.0120 | <2E-16 | |
| **$R^2$** | **0.13** | | | | **0.23** | | | |

Multi-variable linear regression was used to test association of covariates with body fat percentage.

**Table S5:** Correlation between covariates and body fat percentage

**A.**

| | **BFP** | **Albumin** | **SHBG** | **Testosterone** |
|---|---|---|---|---|
| **BFP** | | -0.06 | -0.21 | -0.28 |
| **Albumin** | <1E-4 | | -0.11 | 0.01 |
| **SHBG** | <1E-4 | <1E-4 | | 0.58 |
| **Testosterone** | <1E-4 | 3.15E-02 | <1E-4 | |

**B.**

| | **BFP** | **Albumin** | **SHBG** | **Testosterone** |
|---|---|---|---|---|
| **BFP** | | -0.16 | -0.43 | 0.07 |
| **Albumin** | <1E-5 | | 0.00 | 0.02 |
| **SHBG** | <1E-5 | 0.46 | | -0.02 |
| **Testosterone** | <1E-5 | <1E-5 | <1E-5 | |

A: Males, B: Females
Above diagonal: Spearman correlation coefficient; Below diagonal: p-value

**Table S6:** Association of newly identified BFP loci not been associated with adiposity related traits before with MRI-derived ASAT, VAT and GFAT adjusted for BMI and height [5]

| | | | | | | | | | Agrawal et al | | | | | |
|---|---|---|---|---|---|---|---|---|---|---|---|---|---|---|
| | | | | | Males | | Females | | ASAT | | VAT | | GFAT | |
| CHR | BP (HG19) | SNP | A0 | A1 | BETA | LOG10P | BETA | LOG10P | Beta | p | Beta | p | Beta | p |
| 1 | 106802195 | rs11184828 | A | G | -0.07 | 3.09 | -0.14 | 8.65 | -0.01 | 0.21 | 0.005 | 0.43 | -0.003 | 0.64 |
| 1 | 150055361 | rs116819476 | C | T | 0.17 | 2.79 | 0.33 | 7.39 | 0.006 | 0.71 | 0.008 | 0.78 | -0.007 | 0.83 |
| 1 | 222061973 | rs11577023 | T | C | -0.06 | 2.66 | -0.12 | 7.32 | -0.01 | 0.17 | -0.007 | 0.28 | 0.013 | 0.17 |
| 2 | 54881621 | rs2941584 | T | C | -0.04 | 1.26 | -0.12 | 7.68 | -0.014 | 0.059 | -0.006 | 0.58 | -0.003 | 0.62 |
| 2 | 101414496 | rs2309885 | C | T | 0.1 | 7.44 | 0.03 | 0.65 | -0.014 | 0.07 | -0.002 | 0.59 | 0.023 | 0.0011 |
| 3 | 30071380 | rs7426945 | A | G | 0.05 | 2.35 | 0.12 | 8.06 | 0.008 | 0.21 | -0.003 | 0.54 | -0.006 | 0.44 |
| 4 | 2405062 | rs35802140 | C | G | 0.07 | 1.79 | 0.19 | 8.73 | 0.005 | 0.71 | 0.016 | 0.083 | -0.005 | 0.66 |
| 10 | 126305434 | rs4962671 | T | C | -0.1 | 7.91 | -0.03 | 0.98 | 0.001 | 0.96 | 0.008 | 0.38 | -0.003 | 0.56 |
| 10 | 131430686 | rs524804 | G | A | -0.07 | 3.46 | -0.13 | 8.75 | -0.017 | 0.016 | 0.001 | 0.86 | 0.001 | 0.82 |
| 11 | 66820856 | rs117773425 | C | A | 0.39 | 7.68 | 0 | 0.01 | 0.031 | 0.25 | 0.006 | 0.75 | 0.38 | 0.22 |
| 13 | 76086882 | rs531470369 | C | CTTTTTTT | 0.03 | 1.11 | 0.12 | 7.39 | - | - | - | - | - | - |
| 16 | 25247974 | rs151118254 | C | T | 0.17 | 0.85 | 0.73 | 7.42 | -0.001 | 0.93 | -0.006 | 0.9 | 0.009 | 0.78 |
| 19 | 47282245 | rs56385874 | C | T | 0.05 | 1.3 | 0.17 | 9.32 | 0.011 | 0.22 | -0.009 | 0.31 | 0.019 | 0.038 |
| 20 | 38490795 | rs768147154 | T | TAGAG | -0.12 | 8.1 | -0.06 | 1.77 | -0.009 | 0.34 | 0.008 | 0.25 | -0.016 | 0.062 |
| 20 | 55823762 | rs6127980 | G | A | 0.15 | 7.31 | 0.03 | 0.48 | -0.007 | 0.6 | -0.001 | 0.97 | 0.018 | 0.13 |
| 23 | 43,017,461 | rs5950969 | C | T | 0.10 | 7.46 | -0.02 | 0.25 | - | - | - | - | - | - |
| 23 | 83,562,659 | rs73505165 | G | A | 0.07 | 7.66 | -0.00 | 0.01 | - | - | - | - | - | - |

A0: reference allele, A1: Effect Allele, ASAT: Abdominal subcutaneous adipose tissue, VAT: Visceral adipose tissue, GFAT: Gluteofemoral adipose tissue

**Table S7:** SNPs interacting sex affecting BFP

| CHR | BP (HG19) | ID | A0 | A1 | A1FREQ | INFO | BETA | SE | LOG10P |
|---|---|---|---|---|---|---|---|---|---|
| 4 | 62,634,106 | rs754823863 | T | C | 0.999 | 0.730 | 3.600 | 0.650 | 7.52 |
| 8 | 36,794,458 | rs16885587 | C | T | 0.872 | 0.977 | 0.229 | 0.041 | 7.54 |
| 8 | 36,829,456 | rs61146997 | CTAAA | C | 0.814 | 0.948 | 0.197 | 0.036 | 7.38 |
| 8 | 36,846,435 | rs75772194 | G | A | 0.836 | 0.999 | 0.202 | 0.037 | 7.36 |
| 8 | 36,847,115 | rs10110651 | C | T | 0.836 | 1.000 | 0.203 | 0.037 | 7.43 |
| 8 | 36,847,709 | rs10086016 | C | T | 0.836 | 1.000 | 0.204 | 0.037 | 7.49 |
| 8 | 36,848,038 | rs28735403 | C | T | 0.836 | 0.999 | 0.203 | 0.037 | 7.43 |
| 8 | 36,848,357 | rs16885613 | C | T | 0.836 | 0.999 | 0.203 | 0.037 | 7.42 |
| 8 | 36,849,946 | rs4286946 | G | C | 0.836 | 0.998 | 0.202 | 0.037 | 7.33 |
| 8 | 36,851,901 | rs10955009 | A | C | 0.836 | 0.998 | 0.203 | 0.037 | 7.44 |
| 8 | 36,853,213 | rs10096213 | T | C | 0.836 | 0.995 | 0.204 | 0.037 | 7.48 |
| 8 | 36,853,217 | rs10095380 | C | G | 0.836 | 0.995 | 0.204 | 0.037 | 7.48 |
| 10 | 70,975,897 | rs5030937 | T | C | 0.308 | 0.994 | -0.168 | 0.030 | 7.82 |
| 10 | 70,975,916 | rs5030938 | T | C | 0.308 | 0.994 | -0.167 | 0.030 | 7.80 |
| 10 | 70,976,833 | rs10762264 | A | G | 0.309 | 0.995 | -0.166 | 0.030 | 7.68 |
| 10 | 70,977,308 | rs10998647 | G | T | 0.308 | 0.995 | -0.166 | 0.030 | 7.66 |
| 10 | 70,977,395 | rs10998648 | C | A | 0.308 | 0.996 | -0.167 | 0.030 | 7.73 |
| 10 | 70,979,924 | rs10823318 | T | A | 0.309 | 0.996 | -0.166 | 0.030 | 7.67 |
| 10 | 70,982,136 | rs35696875 | TCA | T | 0.301 | 0.997 | -0.163 | 0.030 | 7.37 |
| 10 | 70,982,941 | rs4746822 | T | C | 0.301 | 0.999 | -0.163 | 0.030 | 7.38 |
| 10 | 70,983,629 | rs9663238 | G | A | 0.301 | 0.999 | -0.164 | 0.030 | 7.44 |
| 10 | 70,983,936 | rs35199395 | C | G | 0.301 | 0.998 | -0.164 | 0.030 | 7.45 |
| 10 | 70,985,267 | rs2394529 | C | G | 0.301 | 0.999 | -0.163 | 0.030 | 7.41 |
| 10 | 70,986,723 | rs9645500 | G | T | 0.301 | 1.000 | -0.164 | 0.030 | 7.47 |
| 17 | 7,568,925 | rs55745760 | T | C | 0.597 | 0.960 | -0.165 | 0.029 | 8.15 |

**Table S8:** The SNPs associated with BFP in Lu et al 2016 paper [7]

| | | | | | Males | | Females | |
|---|---|---|---|---|---|---|---|---|
| **SNP** | **CHR** | **BP (HG18)** | **BP (HG19)** | **Gene** | **Top SNP** | **BP (HG19)** | **Top SNP** | **BP (HG19)** |
| rs543874 | 1 | 176,156,103 | 177,889,480 | SEC16B | rs539515 | 177,889,025 | rs543874 | 177,889,480 |
| rs6755502 | 2 | 625,721 | 635,721 | TMEM18 | rs4407278 | 648,607 | rs6751993 | 635,864 |
| rs6738627 | 2 | 165,252,696 | 165,544,450 | COBLL1 | rs1128249 | 165,528,624 | rs200472737* | 165,544,573 |
| rs2943652 | 2 | 226,816,690 | 227,108,446 | IRS1 | rs2943650 | 227,105,921 | **No Signal** | - |
| rs693839 | 13 | 79,856,289 | 80,958,288 | SPRY2 | **No Signal** | - | **No Signal** | - |
| rs4788099 | 16 | 28,763,228 | 28,855,727 | TUFM | 16:28579915_GT_G | 28,579,915 | rs437564 | 28,818,037 |
| rs1558902 | 16 | 52,361,075 | 53,803,574 | FTO | rs56094641* | 53,806,453 | rs1421085 | 53,800,954 |
| rs9906944 | 17 | 44,446,419 | 47,091,420 | IGF2BP1 | **No Signal** | - | **No Signal** | - |
| rs6567160 | 18 | 55,980,115 | 57,829,135 | MC4R | rs73455668 | 57,985,366 | rs11451426 | 60,189,009 |
| rs757318 | 19 | 18,681,308 | 18,820,308 | CRTC1 | **No Signal** | - | **No Signal** | - |
| rs6857 | 19 | 50,084,094 | 45,392,254 | TOMM40 | rs190712692 | 45,425,178 | rs7412 | 45,412,079 |
| rs3761445 | 22 | 36,925,357 | 38,595,411 | PLA2G6 | rs4821764 | 38,599,364 | rs4820325 | 38,599,978 |

* There are other independent signals in the locus.

#### **Table S9:** Novel loci that are eQTLs based on GTEx v8

| Gencode Id | Gene Symbol | Variant Id | SNP Id | P-Value | NES | Tissue |
|---|---|---|---|---|---|---|
| ENSG00000266472.5 | MRPS21 | chr1_150083277_C_T_b38 | rs116819476 | 0.0000048 | 0.36 | Adipose - Visceral (Omentum) |
| ENSG00000143369.14 | ECM1 | chr1_150083277_C_T_b38 | rs116819476 | 0.000085 | -0.26 | Artery - Tibial |
| ENSG00000266472.5 | MRPS21 | chr1_150083277_C_T_b38 | rs116819476 | 0.00029 | 0.22 | Artery - Tibial |
| ENSG00000143369.14 | ECM1 | chr1_150083277_C_T_b38 | rs116819476 | 0.000056 | 0.63 | Brain - Cerebellar Hemisphere |
| ENSG00000143369.14 | ECM1 | chr1_150083277_C_T_b38 | rs116819476 | 6.40E-08 | 0.92 | Brain - Cerebellum |
| ENSG00000143369.14 | ECM1 | chr1_150083277_C_T_b38 | rs116819476 | 5.60E-09 | 0.49 | Cells - Cultured fibroblasts |
| ENSG00000143363.15 | PRUNE1 | chr1_150083277_C_T_b38 | rs116819476 | 0.00011 | 0.33 | Cells - Cultured fibroblasts |
| ENSG00000143382.14 | ADAMTSL4 | chr1_150083277_C_T_b38 | rs116819476 | 0.00024 | 0.21 | Cells - Cultured fibroblasts |
| ENSG00000266472.5 | MRPS21 | chr1_150083277_C_T_b38 | rs116819476 | 0.00057 | 0.28 | Cells - Cultured fibroblasts |
| ENSG00000143369.14 | ECM1 | chr1_150083277_C_T_b38 | rs116819476 | 3.50E-12 | 1.6 | Cells - EBV-transformed lymphocytes |
| ENSG00000266472.5 | MRPS21 | chr1_150083277_C_T_b38 | rs116819476 | 1.80E-07 | 0.48 | Colon - Sigmoid |
| ENSG00000266472.5 | MRPS21 | chr1_150083277_C_T_b38 | rs116819476 | 0.00021 | 0.31 | Esophagus - Mucosa |
| ENSG00000266472.5 | MRPS21 | chr1_150083277_C_T_b38 | rs116819476 | 1.50E-07 | 0.41 | Esophagus - Muscularis |
| ENSG00000266472.5 | MRPS21 | chr1_150083277_C_T_b38 | rs116819476 | 0.000015 | 0.36 | Heart - Atrial Appendage |
| ENSG00000143374.16 | TARS2 | chr1_150083277_C_T_b38 | rs116819476 | 0.000037 | -0.26 | Heart - Left Ventricle |
| ENSG00000266472.5 | MRPS21 | chr1_150083277_C_T_b38 | rs116819476 | 0.00001 | 0.31 | Nerve - Tibial |
| ENSG00000228126.1 | FALEC | chr1_150083277_C_T_b38 | rs116819476 | 0.00024 | 0.49 | Nerve - Tibial |
| ENSG00000266472.5 | MRPS21 | chr1_150083277_C_T_b38 | rs116819476 | 0.000032 | 0.76 | Pituitary |
| ENSG00000143369.14 | ECM1 | chr1_150083277_C_T_b38 | rs116819476 | 0.0000049 | 0.34 | Thyroid |
| ENSG00000163141.18 | BNIPL | chr1_150083277_C_T_b38 | rs116819476 | 0.000063 | 0.46 | Thyroid |
| ENSG00000143369.14 | ECM1 | chr1_150083277_C_T_b38 | rs116819476 | 0.0000021 | 0.51 | Whole Blood |
| ENSG00000238042.5 | RP11-815M8.1 | chr1_221888631_T_C_b38 | rs11577023 | 2.10E-16 | 0.46 | Lung |
| ENSG00000238042.5 | RP11-815M8.1 | chr1_221888631_T_C_b38 | rs11577023 | 0.00012 | 0.21 | Nerve - Tibial |
| ENSG00000214595.11 | EML6 | chr2_54654484_T_C_b38 | rs2941584 | 1.10E-17 | 0.4 | Artery - Tibial |
| ENSG00000214595.11 | EML6 | chr2_54654484_T_C_b38 | rs2941584 | 5.80E-16 | 0.46 | Esophagus - Muscularis |
| ENSG00000115306.15 | SPTBN1 | chr2_54654484_T_C_b38 | rs2941584 | 1.80E-14 | -0.2 | Whole Blood |
| ENSG00000214595.11 | EML6 | chr2_54654484_T_C_b38 | rs2941584 | 2.60E-11 | 0.39 | Artery - Aorta |
| ENSG00000214595.11 | EML6 | chr2_54654484_T_C_b38 | rs2941584 | 2.10E-08 | 0.48 | Artery - Coronary |
| ENSG00000237887.1 | RPL23AP32 | chr2_54654484_T_C_b38 | rs2941584 | 8.60E-08 | -0.25 | Lung |
| ENSG00000214595.11 | EML6 | chr2_54654484_T_C_b38 | rs2941584 | 1.60E-07 | 0.35 | Esophagus - Gastroesophageal Junction |
| ENSG00000238018.2 | AC093110.3 | chr2_54654484_T_C_b38 | rs2941584 | 2.60E-07 | 0.13 | Artery - Tibial |
| ENSG00000238018.2 | AC093110.3 | chr2_54654484_T_C_b38 | rs2941584 | 3.10E-07 | 0.18 | Muscle - Skeletal |
| ENSG00000237887.1 | RPL23AP32 | chr2_54654484_T_C_b38 | rs2941584 | 4.80E-07 | -0.22 | Thyroid |
| ENSG00000238018.2 | AC093110.3 | chr2_54654484_T_C_b38 | rs2941584 | 8.00E-07 | 0.15 | Skin - Sun Exposed (Lower leg) |
| ENSG00000237887.1 | RPL23AP32 | chr2_54654484_T_C_b38 | rs2941584 | 0.0000013 | -0.45 | Brain - Anterior cingulate cortex (BA24) |
| ENSG00000115306.15 | SPTBN1 | chr2_54654484_T_C_b38 | rs2941584 | 0.0000017 | 0.11 | Skin - Sun Exposed (Lower leg) |
| ENSG00000214595.11 | EML6 | chr2_54654484_T_C_b38 | rs2941584 | 0.0000018 | 0.28 | Colon - Sigmoid |
| ENSG00000214595.11 | EML6 | chr2_54654484_T_C_b38 | rs2941584 | 0.000003 | 0.27 | Adipose - Visceral (Omentum) |
| ENSG00000237887.1 | RPL23AP32 | chr2_54654484_T_C_b38 | rs2941584 | 0.0000034 | -0.34 | Brain - Cerebellar Hemisphere |
| ENSG00000115306.15 | SPTBN1 | chr2_54654484_T_C_b38 | rs2941584 | 0.000005 | 0.12 | Skin - Not Sun Exposed (Suprapubic) |
| ENSG00000237887.1 | RPL23AP32 | chr2_54654484_T_C_b38 | rs2941584 | 0.0000065 | -0.34 | Testis |
| ENSG00000237887.1 | RPL23AP32 | chr2_54654484_T_C_b38 | rs2941584 | 0.0000071 | -0.22 | Adipose - Visceral (Omentum) |
| ENSG00000238018.2 | AC093110.3 | chr2_54654484_T_C_b38 | rs2941584 | 0.0000098 | 0.11 | Nerve - Tibial |
| ENSG00000238018.2 | AC093110.3 | chr2_54654484_T_C_b38 | rs2941584 | 0.000016 | 0.11 | Adipose - Subcutaneous |
| ENSG00000115306.15 | SPTBN1 | chr2_54654484_T_C_b38 | rs2941584 | 0.000018 | -0.14 | Cells - Cultured fibroblasts |
| ENSG00000237887.1 | RPL23AP32 | chr2_54654484_T_C_b38 | rs2941584 | 0.000027 | -0.22 | Adipose - Subcutaneous |
| ENSG00000214595.11 | EML6 | chr2_54654484_T_C_b38 | rs2941584 | 0.000032 | -0.19 | Thyroid |
| ENSG00000238018.2 | AC093110.3 | chr2_54654484_T_C_b38 | rs2941584 | 0.000038 | 0.11 | Esophagus - Gastroesophageal Junction |
| ENSG00000238018.2 | AC093110.3 | chr2_54654484_T_C_b38 | rs2941584 | 0.000072 | 0.11 | Thyroid |
| ENSG00000115306.15 | SPTBN1 | chr2_54654484_T_C_b38 | rs2941584 | 0.0001 | 0.14 | Muscle - Skeletal |
| ENSG00000214595.11 | EML6 | chr2_54654484_T_C_b38 | rs2941584 | 0.00016 | -0.14 | Skin - Sun Exposed (Lower leg) |
| ENSG00000238018.2 | AC093110.3 | chr2_54654484_T_C_b38 | rs2941584 | 0.00024 | 0.099 | Lung |
| ENSG00000159733.13 | ZFYVE28 | chr4_2403335_C_G_b38 | rs35802140 | 5.40E-14 | -0.55 | Colon - Sigmoid |
| ENSG00000159733.13 | ZFYVE28 | chr4_2403335_C_G_b38 | rs35802140 | 1.10E-11 | -0.43 | Esophagus - Muscularis |

| Gencode Id | Gene Symbol | Variant Id | SNP Id | P-Value | NES | Tissue |
|---|---|---|---|---|---|---|
| ENSG00000159733.13 | ZFYVE28 | chr4_2403335_C_G_b38 | rs35802140 | 5.50E-11 | -0.56 | Heart - Atrial Appendage |
| ENSG00000159733.13 | ZFYVE28 | chr4_2403335_C_G_b38 | rs35802140 | 8.20E-09 | -0.33 | Thyroid |
| ENSG00000159733.13 | ZFYVE28 | chr4_2403335_C_G_b38 | rs35802140 | 2.20E-08 | -0.55 | Prostate |
| ENSG00000159733.13 | ZFYVE28 | chr4_2403335_C_G_b38 | rs35802140 | 3.30E-07 | -0.4 | Heart - Left Ventricle |
| ENSG00000159733.13 | ZFYVE28 | chr4_2403335_C_G_b38 | rs35802140 | 4.60E-07 | -0.36 | Esophagus - Gastroesophageal Junction |
| ENSG00000159733.13 | ZFYVE28 | chr4_2403335_C_G_b38 | rs35802140 | 0.0000011 | -0.3 | Adipose - Visceral (Omentum) |
| ENSG00000206113.10 | CFAP99 | chr4_2403335_C_G_b38 | rs35802140 | 0.0000047 | -0.26 | Thyroid |
| ENSG00000159733.13 | ZFYVE28 | chr4_2403335_C_G_b38 | rs35802140 | 0.0000059 | -0.25 | Artery - Tibial |
| ENSG00000159733.13 | ZFYVE28 | chr4_2403335_C_G_b38 | rs35802140 | 0.000023 | -0.27 | Lung |
| ENSG00000063978.15 | RNF4 | chr4_2403335_C_G_b38 | rs35802140 | 0.000024 | 0.11 | Cells - Cultured fibroblasts |
| ENSG00000063978.15 | RNF4 | chr4_2403335_C_G_b38 | rs35802140 | 0.00003 | 0.25 | Colon - Sigmoid |
| ENSG00000159733.13 | ZFYVE28 | chr4_2403335_C_G_b38 | rs35802140 | 0.00015 | -0.2 | Adipose - Subcutaneous |
| ENSG00000203791.14 | METTL10 | chr10_124616865_T_C_b38 | rs4962671 | 3.80E-29 | 0.3 | Cells - Cultured fibroblasts |
| ENSG00000203791.14 | METTL10 | chr10_124616865_T_C_b38 | rs4962671 | 5.40E-28 | 0.32 | Esophagus - Mucosa |
| ENSG00000203791.14 | METTL10 | chr10_124616865_T_C_b38 | rs4962671 | 1.50E-27 | 0.31 | Artery - Tibial |
| ENSG00000203791.14 | METTL10 | chr10_124616865_T_C_b38 | rs4962671 | 2.30E-27 | 0.32 | Adipose - Subcutaneous |
| ENSG00000203791.14 | METTL10 | chr10_124616865_T_C_b38 | rs4962671 | 3.50E-26 | 0.3 | Thyroid |
| ENSG00000203791.14 | METTL10 | chr10_124616865_T_C_b38 | rs4962671 | 2.00E-24 | 0.38 | Breast - Mammary Tissue |
| ENSG00000203791.14 | METTL10 | chr10_124616865_T_C_b38 | rs4962671 | 6.20E-22 | 0.33 | Lung |
| ENSG00000203791.14 | METTL10 | chr10_124616865_T_C_b38 | rs4962671 | 1.60E-21 | 0.31 | Adipose - Visceral (Omentum) |
| ENSG00000203791.14 | METTL10 | chr10_124616865_T_C_b38 | rs4962671 | 1.30E-20 | 0.36 | Artery - Aorta |
| ENSG00000203791.14 | METTL10 | chr10_124616865_T_C_b38 | rs4962671 | 6.80E-20 | 0.26 | Skin - Sun Exposed (Lower leg) |
| ENSG00000203791.14 | METTL10 | chr10_124616865_T_C_b38 | rs4962671 | 4.30E-19 | 0.22 | Muscle - Skeletal |
| ENSG00000203791.14 | METTL10 | chr10_124616865_T_C_b38 | rs4962671 | 1.30E-17 | 0.26 | Skin - Not Sun Exposed (Suprapubic) |
| ENSG00000203791.14 | METTL10 | chr10_124616865_T_C_b38 | rs4962671 | 1.10E-16 | 0.3 | Colon - Transverse |
| ENSG00000203791.14 | METTL10 | chr10_124616865_T_C_b38 | rs4962671 | 3.00E-15 | 0.27 | Testis |
| ENSG00000107902.13 | LHPP | chr10_124616865_T_C_b38 | rs4962671 | 5.60E-15 | -0.15 | Nerve - Tibial |
| ENSG00000203791.14 | METTL10 | chr10_124616865_T_C_b38 | rs4962671 | 7.80E-14 | 0.33 | Stomach |
| ENSG00000203791.14 | METTL10 | chr10_124616865_T_C_b38 | rs4962671 | 8.60E-14 | 0.24 | Nerve - Tibial |
| ENSG00000203791.14 | METTL10 | chr10_124616865_T_C_b38 | rs4962671 | 5.10E-13 | 0.21 | Esophagus - Muscularis |
| ENSG00000203791.14 | METTL10 | chr10_124616865_T_C_b38 | rs4962671 | 3.10E-12 | 0.33 | Heart - Left Ventricle |
| ENSG00000203791.14 | METTL10 | chr10_124616865_T_C_b38 | rs4962671 | 3.20E-12 | 0.33 | Prostate |
| ENSG00000203791.14 | METTL10 | chr10_124616865_T_C_b38 | rs4962671 | 3.30E-12 | 0.36 | Liver |
| ENSG00000203791.14 | METTL10 | chr10_124616865_T_C_b38 | rs4962671 | 1.10E-10 | 0.27 | Heart - Atrial Appendage |
| ENSG00000203791.14 | METTL10 | chr10_124616865_T_C_b38 | rs4962671 | 1.10E-10 | 0.41 | Small Intestine - Terminal Ileum |
| ENSG00000203791.14 | METTL10 | chr10_124616865_T_C_b38 | rs4962671 | 1.50E-10 | 0.45 | Cells - EBV-transformed lymphocytes |
| ENSG00000203791.14 | METTL10 | chr10_124616865_T_C_b38 | rs4962671 | 1.60E-10 | 0.35 | Artery - Coronary |
| ENSG00000203791.14 | METTL10 | chr10_124616865_T_C_b38 | rs4962671 | 2.80E-10 | 0.14 | Whole Blood |
| ENSG00000203791.14 | METTL10 | chr10_124616865_T_C_b38 | rs4962671 | 3.00E-10 | 0.34 | Adrenal Gland |
| ENSG00000203791.14 | METTL10 | chr10_124616865_T_C_b38 | rs4962671 | 5.30E-10 | 0.32 | Brain - Caudate (basal ganglia) |
| ENSG00000203791.14 | METTL10 | chr10_124616865_T_C_b38 | rs4962671 | 8.20E-10 | 0.35 | Pituitary |
| ENSG00000203791.14 | METTL10 | chr10_124616865_T_C_b38 | rs4962671 | 3.00E-09 | 0.21 | Esophagus - Gastroesophageal Junction |
| ENSG00000203791.14 | METTL10 | chr10_124616865_T_C_b38 | rs4962671 | 5.30E-09 | 0.33 | Brain - Nucleus accumbens (basal ganglia) |
| ENSG00000203791.14 | METTL10 | chr10_124616865_T_C_b38 | rs4962671 | 6.70E-09 | 0.43 | Brain - Amygdala |
| ENSG00000203791.14 | METTL10 | chr10_124616865_T_C_b38 | rs4962671 | 7.90E-09 | 0.35 | Brain - Cortex |
| ENSG00000203791.14 | METTL10 | chr10_124616865_T_C_b38 | rs4962671 | 8.30E-09 | 0.35 | Spleen |
| ENSG00000203791.14 | METTL10 | chr10_124616865_T_C_b38 | rs4962671 | 2.80E-08 | 0.2 | Colon - Sigmoid |
| ENSG00000203791.14 | METTL10 | chr10_124616865_T_C_b38 | rs4962671 | 3.20E-08 | 0.41 | Vagina |
| ENSG00000203791.14 | METTL10 | chr10_124616865_T_C_b38 | rs4962671 | 2.00E-07 | 0.32 | Brain - Putamen (basal ganglia) |
| ENSG00000203791.14 | METTL10 | chr10_124616865_T_C_b38 | rs4962671 | 2.70E-07 | 0.27 | Pancreas |
| ENSG00000203791.14 | METTL10 | chr10_124616865_T_C_b38 | rs4962671 | 5.80E-07 | 0.29 | Minor Salivary Gland |
| ENSG00000203791.14 | METTL10 | chr10_124616865_T_C_b38 | rs4962671 | 0.0000065 | 0.24 | Brain - Hypothalamus |
| ENSG00000203791.14 | METTL10 | chr10_124616865_T_C_b38 | rs4962671 | 0.000012 | 0.32 | Uterus |
| ENSG00000107902.13 | LHPP | chr10_124616865_T_C_b38 | rs4962671 | 0.000024 | -0.21 | Brain - Putamen (basal ganglia) |
| ENSG00000107902.13 | LHPP | chr10_124616865_T_C_b38 | rs4962671 | 0.00007 | -0.11 | Artery - Tibial |
| ENSG00000189319.13 | FAM53B | chr10_124616865_T_C_b38 | rs4962671 | 0.000088 | -0.12 | Cells - Cultured fibroblasts |

| Gencode Id | Gene Symbol | Variant Id | SNP Id | P-Value | NES | Tissue |
|---|---|---|---|---|---|---|
| ENSG00000170430.9 | MGMT | chr10_129632422_G_A_b38 | rs524804 | 2.10E-70 | -0.64 | Thyroid |
| ENSG00000170430.9 | MGMT | chr10_129632422_G_A_b38 | rs524804 | 8.80E-64 | -0.63 | Skin - Sun Exposed (Lower leg) |
| ENSG00000170430.9 | MGMT | chr10_129632422_G_A_b38 | rs524804 | 5.10E-62 | -0.73 | Esophagus - Mucosa |
| ENSG00000170430.9 | MGMT | chr10_129632422_G_A_b38 | rs524804 | 7.40E-61 | -0.64 | Adipose - Visceral (Omentum) |
| ENSG00000170430.9 | MGMT | chr10_129632422_G_A_b38 | rs524804 | 2.50E-60 | -0.63 | Adipose - Subcutaneous |
| ENSG00000170430.9 | MGMT | chr10_129632422_G_A_b38 | rs524804 | 7.30E-60 | -0.62 | Skin - Not Sun Exposed (Suprapubic) |
| ENSG00000170430.9 | MGMT | chr10_129632422_G_A_b38 | rs524804 | 1.00E-55 | -0.52 | Artery - Tibial |
| ENSG00000170430.9 | MGMT | chr10_129632422_G_A_b38 | rs524804 | 2.10E-54 | -0.51 | Cells - Cultured fibroblasts |
| ENSG00000170430.9 | MGMT | chr10_129632422_G_A_b38 | rs524804 | 2.50E-50 | -0.62 | Lung |
| ENSG00000170430.9 | MGMT | chr10_129632422_G_A_b38 | rs524804 | 4.90E-48 | -0.37 | Whole Blood |
| ENSG00000170430.9 | MGMT | chr10_129632422_G_A_b38 | rs524804 | 8.10E-47 | -0.71 | Artery - Aorta |
| ENSG00000170430.9 | MGMT | chr10_129632422_G_A_b38 | rs524804 | 1.80E-46 | -0.41 | Muscle - Skeletal |
| ENSG00000170430.9 | MGMT | chr10_129632422_G_A_b38 | rs524804 | 1.80E-43 | -0.54 | Nerve - Tibial |
| ENSG00000170430.9 | MGMT | chr10_129632422_G_A_b38 | rs524804 | 2.70E-41 | -0.56 | Breast - Mammary Tissue |
| ENSG00000170430.9 | MGMT | chr10_129632422_G_A_b38 | rs524804 | 4.30E-41 | -0.56 | Esophagus - Muscularis |
| ENSG00000170430.9 | MGMT | chr10_129632422_G_A_b38 | rs524804 | 1.20E-38 | -0.53 | Heart - Atrial Appendage |
| ENSG00000170430.9 | MGMT | chr10_129632422_G_A_b38 | rs524804 | 6.10E-38 | -0.68 | Colon - Transverse |
| ENSG00000170430.9 | MGMT | chr10_129632422_G_A_b38 | rs524804 | 8.10E-35 | -0.65 | Stomach |
| ENSG00000170430.9 | MGMT | chr10_129632422_G_A_b38 | rs524804 | 3.10E-34 | -0.63 | Esophagus - Gastroesophageal Junction |
| ENSG00000170430.9 | MGMT | chr10_129632422_G_A_b38 | rs524804 | 2.50E-33 | -0.57 | Heart - Left Ventricle |
| ENSG00000170430.9 | MGMT | chr10_129632422_G_A_b38 | rs524804 | 1.80E-30 | -0.54 | Colon - Sigmoid |
| ENSG00000170430.9 | MGMT | chr10_129632422_G_A_b38 | rs524804 | 2.00E-29 | -0.73 | Adrenal Gland |
| ENSG00000170430.9 | MGMT | chr10_129632422_G_A_b38 | rs524804 | 2.50E-28 | -0.54 | Testis |
| ENSG00000170430.9 | MGMT | chr10_129632422_G_A_b38 | rs524804 | 2.60E-27 | -0.48 | Liver |
| ENSG00000170430.9 | MGMT | chr10_129632422_G_A_b38 | rs524804 | 1.30E-26 | -0.65 | Prostate |
| ENSG00000170430.9 | MGMT | chr10_129632422_G_A_b38 | rs524804 | 2.60E-23 | -0.67 | Artery - Coronary |
| ENSG00000170430.9 | MGMT | chr10_129632422_G_A_b38 | rs524804 | 3.00E-23 | -0.63 | Pituitary |
| ENSG00000170430.9 | MGMT | chr10_129632422_G_A_b38 | rs524804 | 6.90E-22 | -0.59 | Pancreas |
| ENSG00000170430.9 | MGMT | chr10_129632422_G_A_b38 | rs524804 | 5.50E-19 | -0.49 | Brain - Caudate (basal ganglia) |
| ENSG00000170430.9 | MGMT | chr10_129632422_G_A_b38 | rs524804 | 3.60E-18 | -0.59 | Brain - Cortex |
| ENSG00000170430.9 | MGMT | chr10_129632422_G_A_b38 | rs524804 | 1.60E-17 | -0.59 | Brain - Cerebellar Hemisphere |
| ENSG00000170430.9 | MGMT | chr10_129632422_G_A_b38 | rs524804 | 4.50E-17 | -0.58 | Brain - Cerebellum |
| ENSG00000170430.9 | MGMT | chr10_129632422_G_A_b38 | rs524804 | 2.20E-16 | -0.46 | Brain - Nucleus accumbens (basal ganglia) |
| ENSG00000170430.9 | MGMT | chr10_129632422_G_A_b38 | rs524804 | 2.50E-16 | -0.53 | Brain - Spinal cord (cervical c-1) |
| ENSG00000170430.9 | MGMT | chr10_129632422_G_A_b38 | rs524804 | 3.40E-16 | -0.6 | Spleen |
| ENSG00000170430.9 | MGMT | chr10_129632422_G_A_b38 | rs524804 | 2.00E-14 | -0.64 | Small Intestine - Terminal Ileum |
| ENSG00000170430.9 | MGMT | chr10_129632422_G_A_b38 | rs524804 | 3.10E-14 | -0.82 | Cells - EBV-transformed lymphocytes |
| ENSG00000170430.9 | MGMT | chr10_129632422_G_A_b38 | rs524804 | 4.70E-14 | -0.49 | Brain - Hypothalamus |
| ENSG00000170430.9 | MGMT | chr10_129632422_G_A_b38 | rs524804 | 1.70E-13 | -0.52 | Brain - Anterior cingulate cortex (BA24) |
| ENSG00000170430.9 | MGMT | chr10_129632422_G_A_b38 | rs524804 | 9.90E-13 | -0.57 | Brain - Hippocampus |
| ENSG00000170430.9 | MGMT | chr10_129632422_G_A_b38 | rs524804 | 1.30E-12 | -0.47 | Brain - Frontal Cortex (BA9) |
| ENSG00000170430.9 | MGMT | chr10_129632422_G_A_b38 | rs524804 | 1.90E-11 | -0.44 | Brain - Putamen (basal ganglia) |
| ENSG00000170430.9 | MGMT | chr10_129632422_G_A_b38 | rs524804 | 3.70E-11 | -0.6 | Minor Salivary Gland |
| ENSG00000170430.9 | MGMT | chr10_129632422_G_A_b38 | rs524804 | 5.70E-11 | -0.57 | Brain - Substantia nigra |
| ENSG00000170430.9 | MGMT | chr10_129632422_G_A_b38 | rs524804 | 3.90E-09 | -0.63 | Uterus |
| ENSG00000170430.9 | MGMT | chr10_129632422_G_A_b38 | rs524804 | 4.60E-09 | -0.45 | Brain - Amygdala |
| ENSG00000170430.9 | MGMT | chr10_129632422_G_A_b38 | rs524804 | 5.30E-08 | -0.58 | Vagina |
| ENSG00000170430.9 | MGMT | chr10_129632422_G_A_b38 | rs524804 | 6.90E-08 | -0.47 | Ovary |
| ENSG00000108001.13 | EBF3 | chr10_129632422_G_A_b38 | rs524804 | 0.000015 | -0.14 | Esophagus - Mucosa |
| ENSG00000172725.13 | CORO1B | chr11_67053385_C_A_b38 | rs117773425 | 0.000049 | 0.42 | Spleen |
| ENSG00000174165.7 | ZDHHC24 | chr11_67053385_C_A_b38 | rs117773425 | 0.000052 | 0.51 | Artery - Tibial |
| ENSG00000255468.6 | RP11-867G23.8 | chr11_67053385_C_A_b38 | rs117773425 | 0.00033 | -0.66 | Thyroid |
| ENSG00000105287.12 | PRKD2 | chr19_46778988_C_T_b38 | rs56385874 | 1.10E-19 | 0.27 | Artery - Tibial |
| ENSG00000181027.10 | FKRP | chr19_46778988_C_T_b38 | rs56385874 | 2.50E-19 | 0.38 | Skin - Sun Exposed (Lower leg) |
| ENSG00000181027.10 | FKRP | chr19_46778988_C_T_b38 | rs56385874 | 4.20E-17 | 0.3 | Muscle - Skeletal |
| ENSG00000181027.10 | FKRP | chr19_46778988_C_T_b38 | rs56385874 | 1.40E-16 | 0.39 | Skin - Not Sun Exposed (Suprapubic) |

| Gencode Id | Gene Symbol | Variant Id | SNP Id | P-Value | NES | Tissue |
|---|---|---|---|---|---|---|
| ENSG00000181027.10 | FKRP | chr19_46778988_C_T_b38 | rs56385874 | 2.10E-16 | 0.54 | Esophagus - Muscularis |
| ENSG00000181027.10 | FKRP | chr19_46778988_C_T_b38 | rs56385874 | 7.70E-16 | 0.4 | Nerve - Tibial |
| ENSG00000105281.12 | SLC1A5 | chr19_46778988_C_T_b38 | rs56385874 | 1.10E-13 | 0.25 | Cells - Cultured fibroblasts |
| ENSG00000105287.12 | PRKD2 | chr19_46778988_C_T_b38 | rs56385874 | 3.30E-13 | 0.16 | Whole Blood |
| ENSG00000105287.12 | PRKD2 | chr19_46778988_C_T_b38 | rs56385874 | 4.60E-13 | 0.32 | Cells - Cultured fibroblasts |
| ENSG00000181027.10 | FKRP | chr19_46778988_C_T_b38 | rs56385874 | 6.40E-13 | 0.34 | Artery - Tibial |
| ENSG00000181027.10 | FKRP | chr19_46778988_C_T_b38 | rs56385874 | 1.90E-12 | 0.35 | Thyroid |
| ENSG00000105287.12 | PRKD2 | chr19_46778988_C_T_b38 | rs56385874 | 1.10E-11 | 0.31 | Nerve - Tibial |
| ENSG00000181027.10 | FKRP | chr19_46778988_C_T_b38 | rs56385874 | 1.30E-11 | 0.35 | Adipose - Subcutaneous |
| ENSG00000181027.10 | FKRP | chr19_46778988_C_T_b38 | rs56385874 | 2.90E-10 | 0.29 | Adipose - Visceral (Omentum) |
| ENSG00000181027.10 | FKRP | chr19_46778988_C_T_b38 | rs56385874 | 3.20E-10 | 0.48 | Colon - Sigmoid |
| ENSG00000181027.10 | FKRP | chr19_46778988_C_T_b38 | rs56385874 | 4.90E-10 | 0.28 | Cells - Cultured fibroblasts |
| ENSG00000181027.10 | FKRP | chr19_46778988_C_T_b38 | rs56385874 | 5.20E-10 | 0.31 | Esophagus - Mucosa |
| ENSG00000105287.12 | PRKD2 | chr19_46778988_C_T_b38 | rs56385874 | 6.30E-10 | 0.23 | Thyroid |
| ENSG00000090372.14 | STRN4 | chr19_46778988_C_T_b38 | rs56385874 | 2.40E-09 | 0.14 | Skin - Sun Exposed (Lower leg) |
| ENSG00000181027.10 | FKRP | chr19_46778988_C_T_b38 | rs56385874 | 7.20E-09 | 0.29 | Colon - Transverse |
| ENSG00000181027.10 | FKRP | chr19_46778988_C_T_b38 | rs56385874 | 3.70E-08 | 0.36 | Breast - Mammary Tissue |
| ENSG00000105287.12 | PRKD2 | chr19_46778988_C_T_b38 | rs56385874 | 6.30E-08 | 0.22 | Esophagus - Muscularis |
| ENSG00000181027.10 | FKRP | chr19_46778988_C_T_b38 | rs56385874 | 9.20E-08 | 0.31 | Artery - Aorta |
| ENSG00000181027.10 | FKRP | chr19_46778988_C_T_b38 | rs56385874 | 1.30E-07 | 0.23 | Lung |
| ENSG00000181027.10 | FKRP | chr19_46778988_C_T_b38 | rs56385874 | 3.20E-07 | 0.33 | Brain - Hippocampus |
| ENSG00000090372.14 | STRN4 | chr19_46778988_C_T_b38 | rs56385874 | 3.70E-07 | 0.13 | Esophagus - Mucosa |
| ENSG00000181027.10 | FKRP | chr19_46778988_C_T_b38 | rs56385874 | 4.80E-07 | 0.3 | Heart - Left Ventricle |
| ENSG00000090372.14 | STRN4 | chr19_46778988_C_T_b38 | rs56385874 | 5.20E-07 | 0.15 | Skin - Not Sun Exposed (Suprapubic) |
| ENSG00000181027.10 | FKRP | chr19_46778988_C_T_b38 | rs56385874 | 6.00E-07 | 0.46 | Prostate |
| ENSG00000105287.12 | PRKD2 | chr19_46778988_C_T_b38 | rs56385874 | 0.0000033 | 0.27 | Pancreas |
| ENSG00000105287.12 | PRKD2 | chr19_46778988_C_T_b38 | rs56385874 | 0.0000048 | 0.25 | Colon - Sigmoid |
| ENSG00000105287.12 | PRKD2 | chr19_46778988_C_T_b38 | rs56385874 | 0.0000049 | 0.17 | Adipose - Visceral (Omentum) |
| ENSG00000105287.12 | PRKD2 | chr19_46778988_C_T_b38 | rs56385874 | 0.0000086 | 0.28 | Pituitary |
| ENSG00000181027.10 | FKRP | chr19_46778988_C_T_b38 | rs56385874 | 0.0000095 | 0.15 | Whole Blood |
| ENSG00000105287.12 | PRKD2 | chr19_46778988_C_T_b38 | rs56385874 | 0.000014 | 0.34 | Liver |
| ENSG00000105287.12 | PRKD2 | chr19_46778988_C_T_b38 | rs56385874 | 0.000021 | 0.18 | Breast - Mammary Tissue |
| ENSG00000181027.10 | FKRP | chr19_46778988_C_T_b38 | rs56385874 | 0.000029 | 0.38 | Esophagus - Gastroesophageal Junction |
| ENSG00000269487.1 | CTB-174O21.2 | chr19_46778988_C_T_b38 | rs56385874 | 0.000031 | -0.31 | Nerve - Tibial |
| ENSG00000105281.12 | SLC1A5 | chr19_46778988_C_T_b38 | rs56385874 | 0.000036 | 0.19 | Artery - Aorta |
| ENSG00000105281.12 | SLC1A5 | chr19_46778988_C_T_b38 | rs56385874 | 0.000039 | 0.14 | Artery - Tibial |
| ENSG00000090372.14 | STRN4 | chr19_46778988_C_T_b38 | rs56385874 | 0.000045 | 0.091 | Whole Blood |
| ENSG00000181027.10 | FKRP | chr19_46778988_C_T_b38 | rs56385874 | 0.000057 | 0.33 | Brain - Cerebellar Hemisphere |
| ENSG00000105287.12 | PRKD2 | chr19_46778988_C_T_b38 | rs56385874 | 0.000066 | 0.15 | Muscle - Skeletal |
| ENSG00000090372.14 | STRN4 | chr19_46778988_C_T_b38 | rs56385874 | 0.000089 | 0.11 | Cells - Cultured fibroblasts |
| ENSG00000101144.12 | BMP7 | chr20_57248706_G_A_b38 | rs6127980 | 4.10E-10 | 0.17 | Skin - Sun Exposed (Lower leg) |
| ENSG00000101146.12 | RAE1 | chr20_57248706_G_A_b38 | rs6127980 | 0.0000036 | 0.15 | Testis |
| ENSG00000101144.12 | BMP7 | chr20_57248706_G_A_b38 | rs6127980 | 0.000015 | 0.18 | Skin - Not Sun Exposed (Suprapubic) |
| ENSG00000132819.16 | RBM38 | chr20_57248706_G_A_b38 | rs6127980 | 0.00002 | 0.22 | Esophagus - Mucosa |

**Table S10:** Association of rs7133378 & rs1716407 and rs56361048 & rs7258937 with BFP in males when both SNPs were in the model

| SNP | CHR | BP (HG19) | A0 | A1 | β | SE | P | Interaction P |
|---|---|---|---|---|---|---|---|---|
| **rs7133378 & rs1716407 both in the model** | | | | | | | | |
| rs7133378 | 12 | 124,409,502 | A | G | -0.15 | 0.02 | 2.53E-12 | 0.72 |
| rs1716407 | 12 | 124,515,218 | A | G | 0.11 | 0.02 | 4.20E-8 | |
| **rs56361048 & rs7258937 both in the model** | | | | | | | | |
| rs56361048 | 19 | 33,885,318 | C | T | -0.10 | 0.03 | 2.66E-4 | 0.95 |
| rs7258937 | 19 | 33,938,800 | T | C | -0.14 | 0.02 | 5.09E-12 | |

Linear regression was used to test association of the two SNPs with BFP in males when Age, $Age^2$, Albumin, $Albumin^2$, SHBG, $SHBG^2$, Testosterone, $Testosterone^2$ and first 10 PCs were in the model.

**Table S11:** Association of rs74841570 and rs11057402 with BFP in females when both SNPs were in the model

| SNP | CHR | BP (HG19) | A0 | A1 | β | SE | P | Int β | Int SE | Int P |
|---|---|---|---|---|---|---|---|---|---|---|
| rs74841570 | 12 | 124,407,903 | C | A | -0.25 | 0.05 | 3.33E-7 | 0.39 | 0.15 | 1.20E-2 |
| rs11057402 | 12 | 124,430,767 | A | T | 0.22 | 0.03 | 5.42E-11 | | | |

Linear regression was used to test association of the two SNPs with BFP in females when Age, $Age^2$, Albumin, $Albumin^2$, SHBG, $SHBG^2$, Testosterone, $Testosterone^2$ and first 10 PCs were in the model.

**Figure S1:** Distribution of body fat percentage in males and females

**A.**

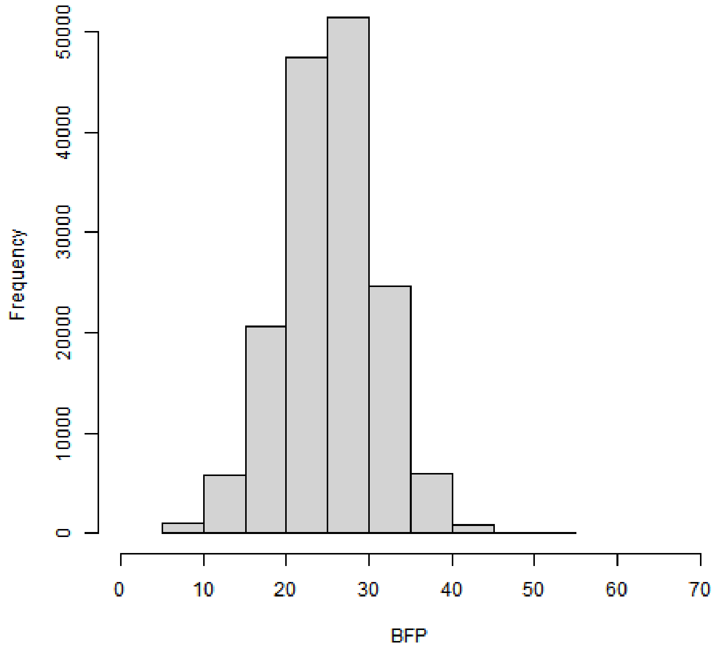

**B.**

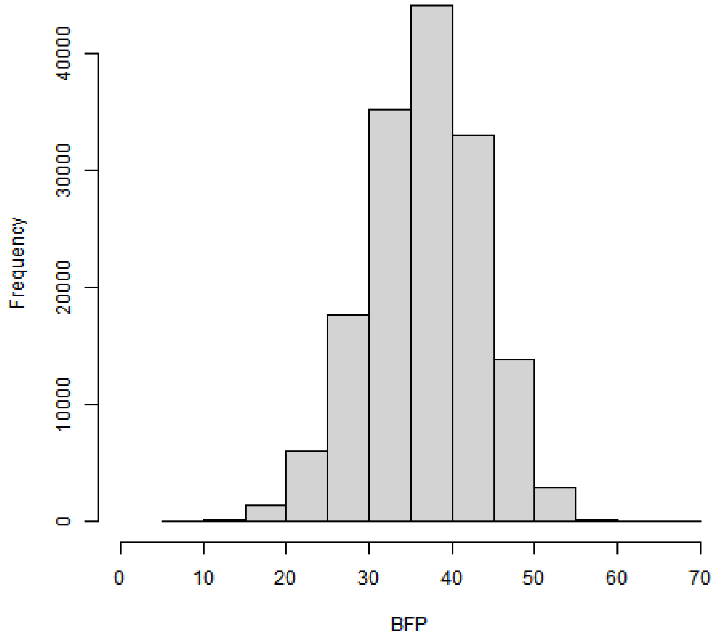

A: Males, B: Females

**Figure S2:** Association of SNPs on Chr X with BFP in males and females

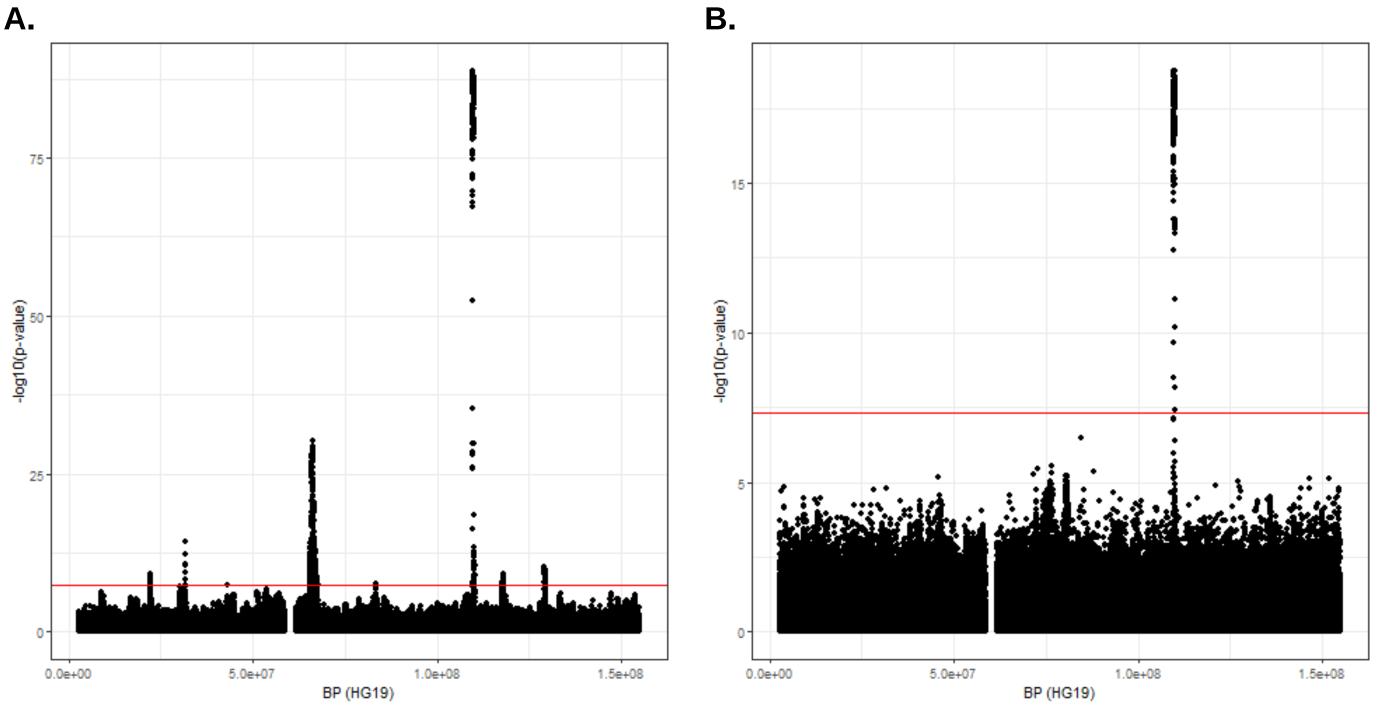

A: Males, B: Females
